# Effects of Social Prescribing on Mental, Physical, and Social Health Outcomes: A Systematic Review and Meta-Analysis of Randomised Trials

**DOI:** 10.64898/2026.08.02.26359484

**Authors:** Xiaoqi Feng, Raju Kanukula, Nicole Evangelidis, Bruce Neal, Patricia M. Davidson, Bogda Koczwara, Kheng Hock Lee, Thomas Astell-Burt

## Abstract

**Importance:** Social prescribing is implemented to address unmet social needs and improve health, but expansion has outpaced evidence from randomized trials.

**Objective:** To quantify the effects of social prescribing on mental, physical, and social health outcomes in adults.

**Data Sources:** Medline, Embase, Cochrane Central, AMED, CINAHL, PsycInfo, Web of Science, NHS EED, CEA registry, clinicaltrials.gov, OpenGrey, and WHO ICTRP (English language). Trials published from 1992 through 2022 were identified from 23 reviews, supplemented by searches from January 2023 through September 2025.

**Study Selection:** Randomized trials comparing usual care or waitlist with interventions facilitating connection to community-based resources delivered by non-health care professionals.

**Data Extraction and Synthesis:** Pairs of reviewers screened studies, extracted data, and assessed risk of bias using Cochrane Risk of Bias 2. Random-effects meta-analyses pooled mean differences or standardized mean differences as Hedges *g* with 95% CIs.

**Main Outcomes and Measures:** Mental health, blood pressure, metabolic and anthropometric outcomes, physical activity, loneliness and social isolation, quality of life, health care use, and adverse events.

**Results:** Thirty-three randomized trials involving 13 714 participants were included. High risk of bias was identified in 40% of trials. Social prescribing was associated with reduced depressive symptoms (7 trials; 1087 participants; standardized mean difference [SMD], −0.23; 95% CI, −0.38 to −0.08), lower systolic blood pressure (11 trials; 2817 participants; mean difference, −2.69 mm Hg; 95% CI, −5.36 to −0.02), increased physical activity (7 trials; 3409 participants; SMD, 0.16; 95% CI, 0.06-0.25), and improved quality of life (10 trials; 3134 participants; SMD, 0.15; 95% CI, 0.01-0.29). No clear benefit was found for anxiety, loneliness and social isolation, glycemic control, blood lipids, anthropometric outcomes, or health care use. One trial reported a process evaluation, 7 included economic evaluations, and adverse events were infrequently reported.

**Conclusions and Relevance:** Social prescribing was associated with modest improvements in depressive symptoms, systolic blood pressure, physical activity, and quality of life. Evidence was lacking for other claimed benefits, and process and economic evaluations were uncommon, identifying priorities for future trials.

**KEY POINTS:** *Question:* What are the effects of social prescribing on mental, physical, and social health outcomes in adults?

*Findings:* In this systematic review and meta-analysis of 33 randomized clinical trials involving 13 714 participants, social prescribing was associated with modest improvements in depressive symptoms, systolic blood pressure, physical activity, and quality of life. There was no clear evidence of benefit for anxiety, loneliness and social isolation, glycemic control, blood lipids, anthropometric outcomes, or health care use; process evaluations, economic evaluations, and analyses of mechanisms were largely absent.

*Meaning:* Social prescribing may improve selected health outcomes, but randomized trial evidence does not support many of the broader benefits commonly attributed to these interventions. More rigorous evaluation of effectiveness, safety, costs, and mechanisms is needed to support robust, equitable, and sustainable implementation.

## INTRODUCTION

Health systems focused mainly on pharmacotherapy and biomedical technologies are ill-equipped for addressing social determinants of health. Social prescribing is used increasingly to address unmet social needs with the goal of keeping people healthy and out of hospital.^1^ Typically, social prescribing is a complex intervention involving a link worker, community health worker or wellbeing coordinator, who provides referrals or facilitated connection to non-clinical community activities and local services informed by personalised assessment, emotional support, and motivational coaching.^2^ These might include referrals to men’s sheds, allotments or walking groups, to choirs, crisis shelters and counselling. Social prescribing is an explicit acknowledgement that the causes of poor health are social and economic, and so too must be the solutions.^3^

Reviews of social prescribing suggest possible benefits for chronic disease prevention^4^ and management,^5^ mental health,^6^ loneliness,^7^ and quality of life^8^ but the quality of the evidence is limited as most of the included studies were non-randomised. Randomised trials are crucial for giving the best possible evidence of effectiveness to inform policy. Since the last comprehensive quantitative summary of the evidence from randomised trials was done in 2023 multiple new randomised trials have completed providing data about a broader range of health outcomes.^9–12^ A recent editorial noted that implementation of social prescribing is ahead of the evidence base, highlighting the need for a high-quality, up-to-date, quantitative synthesis of all the available evidence.^13^ The goal of this systematic review and meta-analysis was to address that shortfall.

## METHODS

### Search strategy

We adopted a two-stage search strategy to identify these trials. We began by reviewing 23 prior systematic reviews^4–8,14–31^ on social prescribing published up to October 2024. All randomised trials included in those reviews were identified and included in our review. We then conducted a systematic search for randomised trials published from January 2023 to November 2024 and updated those searches in September 2025. We systematically searched Medline in Process (Ovid), EMBASE, Cochrane Central, AMED, CINAHL, PsycInfo, Web of Science, NHS EED, CEA registry, clinicaltrials.gov, Open Grey, and WHO ICTRP for English language studies. A hand search of citations was also conducted. Detailed search strategies and the PRISMA checklist are provided in Supplementary Tables 1-8. The review protocol was registered with PROSPERO (CRD42024610742) and reported per PRISMA 2020 guidelines.^32^ The protocol included all populations whereas the current review focusses on adults.

### Trial selection criteria

Interventions were eligible when their core mechanism was facilitated connection, navigation, or referral to community-based resources delivered by a non-healthcare professional, rather than conventional clinical care, education, or motivational interviewing alone. We used a PICO framework: adults with non-communicable diseases; social prescribing interventions delivered by a non-healthcare professional tested via randomised trials with usual care, waitlist, or placebo controls; and primary outcomes including depression, anxiety, blood pressure, blood lipids, haemoglobin A1c, weight, diet, sleep quality, drug and alcohol misuse, loneliness, physical activity, general health, or quality of life. Studies were included if they were (1) randomised, (2) done in adults (≥18 years), (3) addressed non-communicable diseases, and (4) assessed a social prescribing intervention delivered by a non-healthcare professional. Studies done in home, community or primary care settings, with any underpinning theory, delivered face-to-face or online, of any duration, and using any resources were eligible. Interventions delivered by healthcare professionals, or in hospitals, or those providing only advice, education, or motivational interviewing without referral to a community resource were ineligible. Eligible control conditions were usual care, waitlist, or placebo. Quasi-experimental and observational studies were excluded.

### Data extraction

Two reviewers (RK and NE) independently screened titles, abstracts, and full texts in Covidence software. Disagreements were resolved by discussion or a third reviewer (TAB). The same reviewers independently extracted data on study and intervention characteristics and outcomes using a standard sheet. We extracted data from published trials and verified outcomes that meta-analysed by previously published systematic reviews. They assessed risk of bias with the Cochrane Risk of Bias 2 tool.^33^ We standardized units where needed (e.g., cholesterol: 1 mmol/L = 38.67 mg/dL; triglycerides: 1 mmol/L = 88.57 mg/dL) to the International System of Units.

### Outcomes

Outcomes of interest were informed by a recent outcome mapping review of social prescribing interventions.^34^ These included blood pressure (SBP, DBP), blood pressure control, quality of life, anxiety, depression, lipids (LDL, TC, HDL, TG), physical activity, glycated haemoglobin A1c, anthropometrics (BMI, weight, waist to hip ratio, waist circumference), loneliness and social isolation. Device-measured outcomes (n=12) included blood pressure (n=8),^35–42^ weight (n=3),^35,36,43^ and physical activity (n=1).^10^ Self-reported outcomes were used for physical activity (n=2),^40,44^ mental health (n=6),^11,12,36,45,46^ and dietary behaviours (n=2).^36,41^

### Data synthesis

We pooled outcomes reported by two or more studies using RevMan^47^ to compare post-intervention data between randomised arms. Medians, interquartile ranges, 95%CIs, and SEs were converted to means and SD per Cochrane guidelines.^48^ For multi-arm trials, only data from the social prescribing arm versus standard care were included. Single study results were described narratively. Standardized mean differences (SMD) using Hedges’s g with 95% confidence intervals (CIs) were estimated for outcomes measured on different scales, such as quality of life and mental health. Mean differences (MD) with 95% CIs were estimated for body mass index, weight, and blood pressure. For dichotomous outcome such as blood pressure control, odds ratio with 95% CI were calculated. We considered both clinical and methodological heterogeneity, however these trials are bound to have heterogeneity due to various population types, interventions and follow-up times. Hence, we quantified the statistical heterogeneity and assessed by χ² and I² tests^48^ and conducted subgroup analyses wherever possible. Random-effects models with restricted maximum likelihood estimators were used if I² > 50%, else fixed effects were used.^49^ Funnel plots were used to assess publication bias when ≥10 studies contributed to a meta-analysis.^50^ Statistical significance was set at 5%.

### Assessing certainty in the findings

GRADE was used to grade evidence certainty, with a Summary of Findings generated via GRADEpro (McMaster University, Canada).^51^ This included all outcomes, absolute risks, relative risk estimates, and quality ratings based on risk of bias, directness, heterogeneity, precision, and publication bias.

### Patient and public involvement

Patients and members of the public were not involved in the design and execution of the study.

## RESULTS

### Search results

We identified 5,788 citations from the database searches with 4,288 remaining after duplicates were removed (Figure 1). Screening titles, abstracts, and 142 full text articles led to 13 eligible randomised trials being identified in the data spanning January 2023–October 2025. This provided for a total of 33 eligible trials (35 reports) after including those identified (n=20 trials) from earlier reviews published to December 2022.^9–12,35–46,52–70^ This represents a 62% increase in randomised trials since January 2023 (Supplementary Figures). Inclusion and exclusion criteria and details of 130 excluded studies are listed in Supplementary Tables 10 and 11. The inter-rater reliability of search results yielded proportionate agreement of 0.859 at the full-text scale. We also found 19 ongoing trials, mostly still recruiting, with expected completion dates between December 2025 and December 2028.

**Figure 1:**
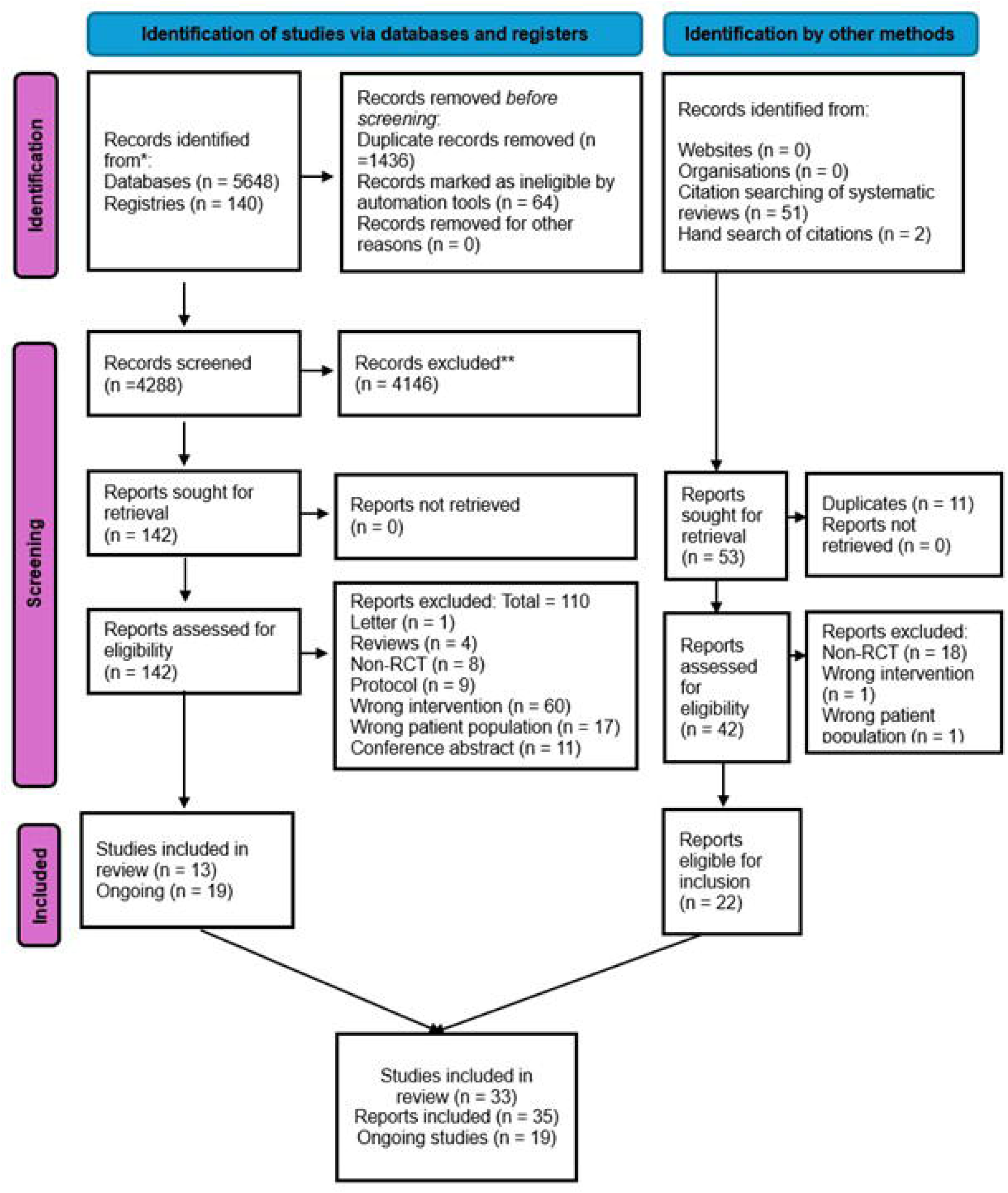
PRISMA chart.

### Characteristics of included trials

There were 28 trials with a two-arm parallel design,^9,10,12,36,37,39–41,43–46,52–68,70^ four with three-arm parallel designs^35,38,42,69^ and one feasibility randomised trial (Supplementary Tables).^11^ One reported process evaluation,^9^ seven included health economic evaluations^11,35,45,58,65,66,70^ and twelve trials were theory-based using a framework such as social cognitive theory.^12,35,46,53,55–57,62–64,69^

The 33 trials involved 13,714 participants, with 19 conducted in the USA,^9,12,38,41–43,53–59,62–65,68,69^ 8 in the UK,^35–37,44,45,67,70^ and one each in Ireland, Belgium, Japan, Spain, Kenya, and China. Average participant age ranged from 50 to 61 years, with 52.5% to 76.5% female participants in the trials. Most included people with diabetes (n=8),^9,36,41,46,55,56,63,68^ hypertension (n=3),^37,39,61^ overweight (n=2),^37,66^ or mental health conditions (n=2).^64,66^ Overall, there were 14% of participants that withdrew prior to trial completion citing lack of interest, home relocation, personal issues, or due to death.

The trials reported 148 different outcomes that we grouped into 30 broad categories based on a recent outcome mapping review^34^ (supplementary material). There were 17 outcomes for which there were two or more trials from which data could be quantitatively summarised (Figure 2). Follow-up periods ranged from 4 weeks^45^ to 36 months,^58^ with longer follow-ups focusing on quality of life, mental health, and blood pressure.^42,57,67^

**Figure 2:**
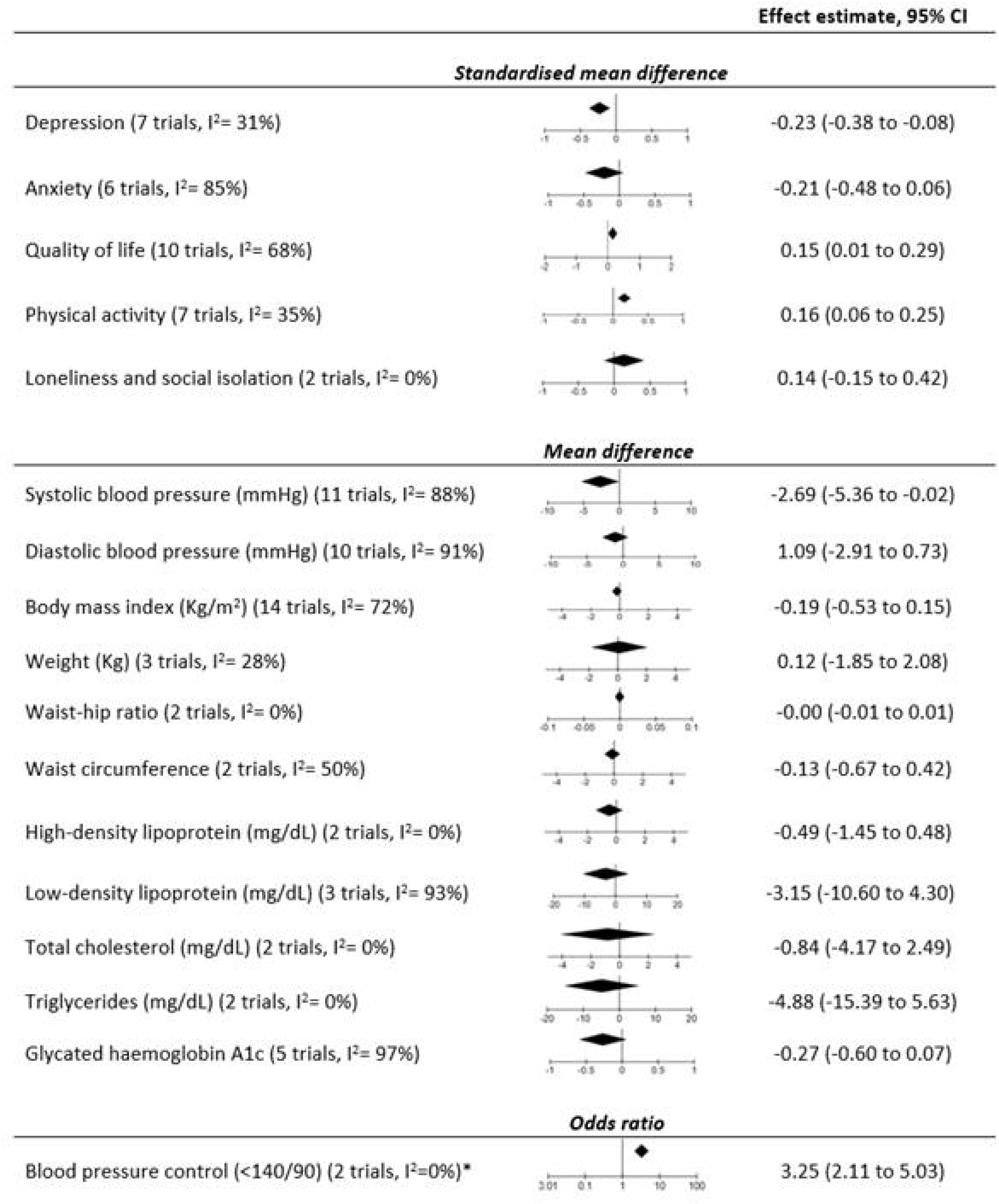
Effects of social prescribing on health outcomes.

Risk of bias was deemed high in 40% of the trials (4011 participants) (Figure S20), with key reasons being poor outcome measurement, missing data, underpowered analyses,^59^ and poor reporting of randomization processes.^37,55,64,69^ In addition nine lacked trial protocols^10,11,45,53,59,61,66,67,69^ and in the nine trials^9,36,56,58,59,61–64^ that reported on the fidelity of intervention delivery, high fidelity was reported by two.^36,56^

### Features of the social prescribing interventions

Most trials had a 6-month intervention period (n=8) but the range was from 5 weeks^61^ to 24 months.^58^ Social prescribing interventions were heterogenous as expected to attend to specific unmet health and social needs and reflecting contemporary international practice including community health worker programs (n=5), walking programs (n=6), disease management programs (n=5), as well as supervised physical activity, psychological support, and online peer leadership programs (n=18). Most were delivered in person (n=28), with 6 conducted remotely.^9,11,12,52,58,64^ Seven trials provided financial incentives (total $10 to $168) as part of the intervention,^12,35,39,42,53,57,59^ one offered free program access,^53^ and another discounted leisure centre entry costs.^35^ Details of interventions characteristics are reported in Supplementary Table 13.

### Effects of social prescribing on mental health outcomes

There was a clear benefit for depression (SMD, −0.23, 95% CI −0.38 to −0.08, 7 trials, I^2^ =31%) but no clear effect for anxiety, loneliness or social isolation^52,64^ (Figure 2 and Supplementary Figures). One trial assessed chronic stress with no clinical differences between groups,^71^ though one other trial showed a reduction of cortisol concentrations.^42^

### Effects of social prescribing on clinical outcomes

There was a significant reduction in systolic blood pressure (SBP) (MD, −2.69, 95% CI −5.36 to −0.02, I^2^ =88%) but not diastolic blood pressure (DBP) (Figure 2 and Supplementary Figures). There was also a greater likelihood of achieving a blood pressure control target of <140/90 mm Hg (OR 3.25, 95% CI 2.11 to 5.03, p<0.001, I^2^= 0.0%) (Supplementary Figures 7), though funnel plots indicated potential publication bias for blood pressure outcomes. Subgroup analysis based on studies included people with diabetes found no significant reduction in SBP and DBP. There was no effect on HbA1c in the five trials that contributed to the meta-analysis (Figure 2 and Supplementary Figure 8) and no effect on insulin resistance in the one trial that reported that outcome.^42^ Subgroup analysis of studies included participants with diabetes also found no reduction in HbA1C levels. Six trials reported on lipids, with no observed effects on total cholesterol, high-density lipoprotein cholesterol, low-density lipoprotein cholesterol, or triglycerides (Figure 2 and Supplementary Figures). Fifteen trials reported anthropometric outcomes without clear benefits for any of BMI, weight, waist to hip ratio or waist circumference (Figure 2 and Supplementary Figures 13-16). One trial also reported null effects on total lean mass and total fat mass.^53^

### Effects of social prescribing on physical activity and quality of life

Seven studies with 3409 participants reported quantitative data on physical activity though a further 6 appear to have recorded data on physical activity. In the 7 studies with data, physical activity was improved by social prescribing compared to control (Sup. Figure 18; Hedges g, 0.16, 95% CI 0.06 to 0.25, I^2^ =35%). Among the 10 trials that reported quantitative data on quality of life a significant improvement was observed (SMD 0.15, 95% CI 0.01 to 0.29, I^2^ =68%) (Figure 2 Sup. Figure 19). There were another 3 trials that collected but did not report quality of life data.

### Effects of social prescribing on serious adverse events and death

Two studies reported adverse events with one reporting 4 falls compared to none in the intervention versus control groups,^35^ and another reporting more non-serious short term injuries related to physical activity (e.g., shoulder injury) in the intervention group.^36^ Seven studies reported deaths^9,38,54,55,58,61,67^ with similar numbers of deaths across randomised groups (intervention n=109, control n=98) and no increase in risk.

### Effects on other outcomes

There were data about multiple other outcomes reported by individual studies or in formats that could not be quantitively summarised. These outcomes included sleep quality,^9,62^ diet,^36,41,69^ social support,^45,59,61^ drug and alcohol misuse,^59^ diabetic distress,^38^ shelter use,^59^ financial needs,^9^ healthcare usage,^9,11,35,46,58,63–65^ healthcare costs,^11,35,45,58,65,66,70^ health literacy,^9,46,54–56^ disease self-management,^46^ happiness,^10^ civic participation,^10^ satisfaction with healthcare decision-making,^58^ life satisfaction,^58^ reciprocal behaviour,^10^ patient activation,^46,58,63^ social cohesion^10^ and general health.^45^ For none was there compelling evidence of clear benefits or harms from social prescribing (Supplementary Tables). The main findings were summarised by mapping the results into the Pathway Domains Framework adapted for social prescribing^72^ (Figure 3).

**Figure 3:**
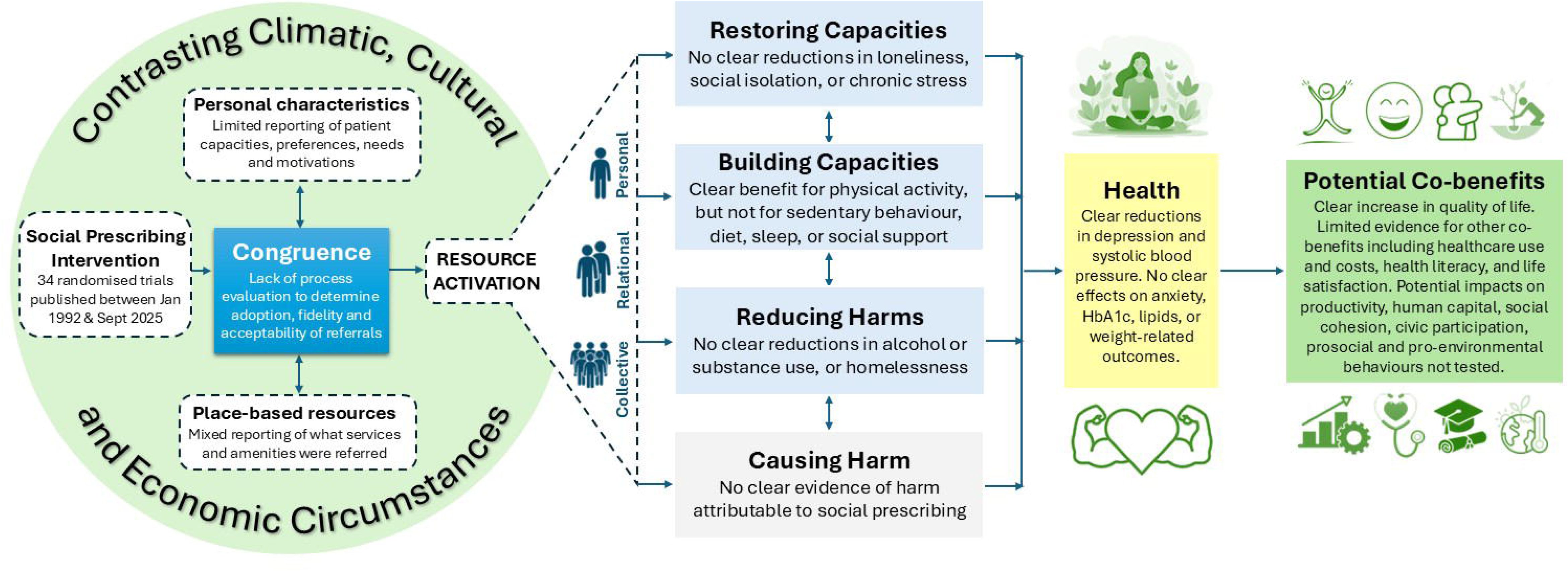
Review findings within the Pathway Domains Framework for Social Prescribing.

## DISCUSSION

New randomised trials of social prescribing reported over the last 2-3 years have added significantly to the evidence base. There is now sufficient evidence to conclude that social prescribing interventions can likely reduce depressive symptoms, lower blood pressure, improve blood pressure control, promote physical activity, and enhance quality of life. Longer trial follow-up with maintenance periods, economic evaluation, and commitment to device-based data ascertainment for some outcomes (e.g., physical activity) will strengthen the quality of evidence. Consideration might also be given to different forms of control, for example, a waitlist option to augment care as usual, and where possible, alternative interventions to establish comparative effectiveness (e.g., with respect to pharmacological options for certain health conditions).

Nevertheless, implementation of social prescribing in many countries has outpaced trial-based evidence for many commonly claimed outcomes, such as loneliness and social isolation.^27^ While there are compelling reasons why social prescribing may have wider benefits,^72^ there also remains insufficient evidence to conclude these interventions can favourably influence anxiety, insulin resistance, glucose, lipids, BMI, sleep quality, and alcohol consumption. There was also no clear evidence of social prescribing reducing healthcare use or costs. Adverse events were rare and reported in two trials.

Synthesis of the trials within the Pathway Domains Framework adapted for social prescribing^72^ indicates that the weight of the research effort thus far has been on assessing specific health outcomes like blood pressure and depression alongside quantifying impact on physical activity in the building capacities domain pathway. Assessment of co-benefits was restricted to quality of life, healthcare use and cost, with limited attention to potential impacts on sustainability, human capital and labour market outcomes. The restoring capacities and reducing harms domain pathways were under-researched, with little data for key outcomes like loneliness.^52^ More trials are needed to assess variables in these domain pathways both as endpoints in their own right, but also to understand how they might act as mediating pathways for physical and mental health outcomes. Investment is needed specifically in randomised trials, from pilot to effectiveness and implementation, and the mechanisms through which they have effects.

Process evaluations were largely absent, with just a few studies reporting measures of intervention acceptability, and the fidelity with which program delivery was achieved. As a consequence, the extent to which the unmet health and social needs, preferences and capacities (i.e., ‘congruence’ in the Pathway Domains Framework) were met by the social prescribing intervention is not clear. Studies were heterogenous in the design of their interventions, which often included multiple elements, without explicit assessment of which intervention components were effective and which were not. The absence of such data makes it difficult to make solid recommendations with respect to implementation and scaling-up programs. Thus, more randomised trials are needed not only from the countries where investment has already been made, but also from a diverse range of contexts so that social prescribing is adaptable to the diverse cultural, climatic and economic contexts where it is emerging.^72,73^ It is also crucial to appreciate that practices aligned with what is commonly known as social prescribing today long pre-date use of that term.^74–77^ In countries where social prescribing has been implemented, challenges reported include variations in awareness among health professionals,^78^ stigma attached to mental health and loneliness presenting barriers,^79,80^ inequities in access to link workers,^81,82^ high workloads and a felt lack of training and integration in health services,^83–85^ and a lack of consensus on educational standards for link workers. Co-design and co-production of randomised trials with communities as co-researchers is critical to their acceptance, implementation and impact.

Our review provides the first quantitative meta-analyses of randomised trials describing the effects of social prescribing on depression, anxiety, loneliness, and quality of life, while extending and enhancing prior results for disease biomarkers. One limitation is the considerable heterogeneity within and between the trials. This heterogeneity reflects the inherently personalised and context-specific nature of social prescribing interventions, although it reduces certainty of the pooled estimates for some outcomes. Other limitations of the evidence include measurement, for example, physical activity was self-reported which is prone to bias; trials using device-measured movement behaviours are needed. There was some evidence of reporting bias in the quantitative analyses of the data, although this may under-represent the reporting bias problem because many studies mentioned collecting outcomes that they did not provide outcome data for. We restricted our searches to English language studies and we acknowledge that some studies may have been missed as a consequence. There were also multiple studies identified as at high risk of bias. The majority of the included studies were from developed countries which limits generalisability of the overview conclusions. The adaptation of implementation strategies to the healthcare system in different countries may not be feasible due to accessibility, availability of community services, numbers of link workers and equivalent professionals, technical support systems and cultural differences.

## Recommendations for future research

Overall, trial evidence supports likely benefits of social prescribing for reducing high blood pressure and depression,^50^ with a co-benefit of improved quality of life. Reports also suggest that physical activity can be improved though device-based outcome measurement is needed. There needs to be investment in well-powered randomised trials with longer follow-up and maintenance periods, focussed on a wide range of under-researched health outcomes (e.g., loneliness) as well as social, economic and environmental co-benefits. Finally, it should be a requirement that publications routinely report effectiveness, cost-effectiveness, and process outcomes of social prescribing interventions to ensure robust and transparent evaluation that will provide best evidence for impacts on population health and health equity.

## Supporting information

PRISMA check list

## Data Availability

All data produced in the present work are contained in the manuscript

## Data sharing

Data in this systematic review and meta-analysis are extracted from published studies available elsewhere. All processed data are presented in this Article and the appendix.

## Declaration of interests

We declare no competing interests.

## Funding

TAB secured funding to support RK and NE from the University of Sydney. TAB is funded by the Australian Research Council.

## Author contributions

XF and TAB co-conceptualised and designed the study, contributed to the review design, led the writing of the initial draft and revisions. TAB secured funding and supervised research staff. RK contributed to review design, led the systematic review method and results, and contributed to the writing of the initial draft and revisions. NE contributed to the systematic review method and results, and contributed to the writing of the initial draft and revisions. PMD, BK, MT, BN and KHL reviewed and provided critical revisions. All authors have reviewed the submitted manuscript.

The corresponding author attests that all listed authors meet authorship criteria and that no others meeting the criteria have been omitted

## Acknowledgement

We used the Gemini 2.5 Flash model to concisely generate the intervention descriptions and we verified the text.

## Competing interests

All authors have completed the ICMJE uniform disclosure form at http://www.icmje.org/disclosure-of-interest/ and declare: no support from any organisation for the submitted work; no financial relationships with any organisations that might have an interest in the submitted work in the previous three years; no other relationships or activities that could appear to have influenced the submitted work.

## Notes

### Competing Interest Statement

The authors have declared no competing interest.

## REFERENCES

1. Morse DF, Sandhu S, Mulligan K, et al. Global developments in social prescribing. BMJ Global Health. 2022;7(5)

2. Lee KH, Low LL, Lu SY, Lee CE. Implementation of social prescribing: lessons learnt from contextualising an intervention in a community hospital in Singapore. The Lancet Regional Health–Western Pacific. 2023;35

3. Rose G. Sick individuals and sick populations. International Journal of Epidemiology. 1985;14:32–38.

4. Htun HL, Teshale AB, Cumpston MS, et al. Effectiveness of social prescribing for chronic disease prevention in adults: a systematic review and meta-analysis of randomised controlled trials. J Epidemiol Community Health. 2023;77(4):265–276.

5. O’Sullivan DJ, Bearne LM, Harrington JM, Cardoso JR, McVeigh JG. The effectiveness of social prescribing in the management of long-term conditions in community-based adults: a systematic review and meta-analysis. Clinical Rehabilitation. 2024;38(10):1306–1320.

6. Cooper M, Avery L, Scott J, et al. Effectiveness and active ingredients of social prescribing interventions targeting mental health: a systematic review. BMJ open. 2022;12(7):e060214.

7. Reinhardt GY, Vidovic D, Hammerton C. Understanding loneliness: a systematic review of the impact of social prescribing initiatives on loneliness. Perspectives in public health. 2021;141(4):204–213.

8. Kiely B, Croke A, O’Shea M, et al. Effect of social prescribing link workers on health outcomes and costs for adults in primary care and community settings: a systematic review. BMJ open. 2022;12(10):e062951.

9. McQueen A, von Nordheim D, Caburnay C, et al. A Randomized Controlled Trial Testing the Effects of a Social Needs Navigation Intervention on Health Outcomes and Healthcare Utilization among Medicaid Members with Type 2 Diabetes. International journal of environmental research and public health. 2024;21(7):936.

10. Yamashita R, Sato S, Sakai Y, et al. Effects of small community walking intervention on physical activity, well-being, and social capital among older patients with cardiovascular disease in the maintenance phase: A randomized controlled trial. Journal of Physical Therapy Science. 2024;36(3):128–135.

11. Kiely B, Hobbins A, Boland F, et al. An exploratory randomised trial investigating feasibility, potential impact and cost effectiveness of link workers for people living with multimorbidity attending general practices in deprived urban communities. BMC Primary Care. 2024;25(1):233.

12. Ugarte DA, Cumberland WG, Singh P, Saadat S, Garett R, Young SD. A HOPE Online Community Peer Support Intervention for Help Seeking: A Randomized Controlled Trial. Psychiatric Services. 2023;74(6):648–651.

13. Lancet T. Social prescribing: bringing community back to health? 2025. p. 103.

14. Percival A, Newton C, Mulligan K, Petrella RJ, Ashe MC. Systematic review of social prescribing and older adults: where to from here? Family medicine and community health. 2022;10(Suppl 1):e001829.

15. Vidovic D, Reinhardt GY, Hammerton C. Can social prescribing foster individual and community well-being? A systematic review of the evidence. International journal of environmental research and public health. 2021;18(10):5276.

16. Napierala H, Krüger K, Kuschick D, Heintze C, Herrmann WJ, Holzinger F. Social prescribing: systematic review of the effectiveness of psychosocial community referral interventions in primary care. International journal of integrated care. 2022;22(3):11.

17. Anderst J, Hunter K, Andersen M, et al. Screening and social prescribing in healthcare and social services to address housing issues among children and families: a systematic review. Bmj Open. 2022;12(4):e054338.

18. Dash S, McNamara S, De Courten M, Calder R. Social prescribing for suicide prevention: a rapid review. Frontiers in public health. 2024;12:1396614.

19. Pescheny JV, Pappas Y, Randhawa G. Facilitators and barriers of implementing and delivering social prescribing services: a systematic review. BMC health services research. 2018;18(1):86.

20. Pescheny JV, Randhawa G, Pappas Y. The impact of social prescribing services on service users: a systematic review of the evidence. European journal of public health. 2020;30(4):664–673.

21. Zhang CX, Wurie F, Browne A, et al. Social prescribing for migrants in the United Kingdom: a systematic review and call for evidence. Journal of Migration and Health. 2021;4:100067.

22. Dubbeldeman EM, Kiefte-de Jong JC, Ardesch FH, et al. Intervention Characteristics and Mechanisms and Their Relationship with the Influence of Social Prescribing: A Systematic Review. Health & Social Care in the Community. 2024;2024(1):5597259.

23. Smith TO, Jimoh OF, Cross J, et al. Social prescribing programmes to prevent or delay frailty in community-dwelling older adults. Geriatrics. 2019;4(4):65.

24. Rapo E, Johansson E, Jonsson F, Hörnsten Å, Lundgren AS, Nilsson I. Critical components of social prescribing programmes with a focus on older adults-a systematic review. Scandinavian Journal of Primary Health Care. 2023;41(3):326–342.

25. Grover S, Sandhu P, Nijjar GS, et al. Older adults and social prescribing experience, outcomes, and processes: a meta-aggregation systematic review. Public Health. 2023;218:197–207.

26. Cooper M, Flynn D, Avery L, et al. Service user perspectives on social prescribing services for mental health in the UK: a systematic review. Perspectives in public health. 2023;143(3):135–144.

27. Liebmann M, Pitman A, Hsueh Y-C, Bertotti M, Pearce E. Do people perceive benefits in the use of social prescribing to address loneliness and/or social isolation? A qualitative meta-synthesis of the literature. BMC health services research. 2022;22(1):1264.

28. Gordon K, Gordon L, Basu AP. Social prescribing for children and young people with neurodisability and their families initiated in a hospital setting: a systematic review. BMJ open. 2023;13(12):e078097.

29. Thomas G, Lynch M, Spencer LH. A systematic review to examine the evidence in developing social prescribing interventions that apply a co-productive, co-designed approach to improve well-being outcomes in a community setting. International Journal of Environmental Research and Public Health. 2021;18(8):3896.

30. Iverson T, Alfares H, Nijjar GS, et al. A rapid systematic review of the effect of health or peer volunteers for diabetes self-management: Synthesizing evidence to guide social prescribing. PLOS Global Public Health. 2024;4(12):e0004071.

31. Al-Khudairy L, Ayorinde A, Ghosh I, et al. Evidence and methods required to evaluate the impact for patients who use social prescribing: a rapid systematic review and qualitative interviews. Health and Social Care Delivery Research. 2022;10(29):1–88.

32. Page MJ, McKenzie JE, Bossuyt PM, et al. The PRISMA 2020 statement: an updated guideline for reporting systematic reviews. bmj. 2021;372

33. Sterne JA, Savović J, Page MJ, et al. RoB 2: a revised tool for assessing risk of bias in randomised trials. bmj. 2019;366

34. Sonke J, Manhas N, Belden C, et al. Social prescribing outcomes: a mapping review of the evidence from 13 countries to identify key common outcomes. Frontiers in Medicine. 2023;10:1266429.

35. Isaacs AJ, Critchley JA, Tai SS, et al. Exercise Evaluation Randomised Trial (EXERT): a randomised trial comparing GP referral for leisure centre-based exercise, community-based walking and advice only. Health Technol Assess. Mar 2007;11(10):1–165, iii-iv. doi:10.3310/hta11100

36. Smith JR, Greaves CJ, Thompson JL, et al. The community-based prevention of diabetes (ComPoD) study: a randomised, waiting list controlled trial of a voluntary sector-led diabetes prevention programme. International Journal of Behavioral Nutrition and Physical Activity. 2019;16:1–14.

37. Taylor AH, Doust J, Webborn N. Randomised controlled trial to examine the effects of a GP exercise referral programme in Hailsham, East Sussex, on modifiable coronary heart disease risk factors. Journal of Epidemiology & Community Health. 1998;52(9):595–601.

38. Spencer MS, Kieffer EC, Sinco B, et al. Outcomes at 18 Months From a Community Health Worker and Peer Leader Diabetes Self-Management Program for Latino Adults. Diabetes Care. Jul 2018;41(7):1414–1422. doi:10.2337/dc17-0978

39. Mbuthia GW, Mwangi J, Magutah K, Oguta JO, Ngure K, McGarvey ST. Preliminary efficacy of a community health worker homebased intervention for the control and management of hypertension in Kiambu County, Kenya-a randomized control trial. Plos one. 2024;19(8):e0293791.

40. Lamb SE, Bartlett H, Ashley A, Bird W. Can lay-led walking programmes increase physical activity in middle aged adults? A randomised controlled trial. Journal of Epidemiology & Community Health. 2002;56(4):246–252.

41. Gray KE, Hoerster KD, Taylor L, Krieger J, Nelson KM. Improvements in physical activity and some dietary behaviors in a community health worker-led diabetes self-management intervention for adults with low incomes: results from a randomized controlled trial. Transl Behav Med. Dec 14 2021;11(12):2144–2154. doi:10.1093/tbm/ibab113

42. Wagner JA, Bermúdez-Millán A, Buckley TE, et al. Community-based diabetes prevention randomized controlled trial in refugees with depression: effects on metabolic outcomes and depression. Scientific reports. 2023;13(1):8718.

43. Islam NS, Wyatt LC, Ali SH, et al. Integrating community health workers into community-based primary care practice settings to improve blood pressure control among South Asian immigrants in New York City: results from a randomized control trial. Circulation: Cardiovascular Quality and Outcomes. 2023;16(3):e009321.

44. Harrison RA, Roberts C, Elton PJ. Does primary care referral to an exercise programme increase physical activity one year later? A randomized controlled trial. Journal of Public Health. 2005;27(1):25–32.

45. Grant C, Goodenough T, Harvey I, Hine C. A randomised controlled trial and economic evaluation of a referrals facilitator between primary care and the voluntary sector. Bmj. 2000;320(7232):419–423.

46. Kangovi S, Mitra N, Grande D, Huo H, Smith RA, Long JA. Community Health Worker Support for Disadvantaged Patients With Multiple Chronic Diseases: A Randomized Clinical Trial. Am J Public Health. Oct 2017;107(10):1660–1667. doi:10.2105/ajph.2017.303985

47. Review Manager (RevMan). Version 7.2.0. The Cochrane Collaboration; 2024. revman.cochrane.org.

48. Higgins JP, Thompson SG, Deeks JJ, Altman DG. Measuring inconsistency in meta-analyses. bmj. 2003;327(7414):557–560.

49. Raudenbush SW. Analyzing effect sizes: Random-effects models. The handbook of research synthesis and meta-analysis. 2009;2:295–316.

50. Sterne JA, Sutton AJ, Ioannidis JP, et al. Recommendations for examining and interpreting funnel plot asymmetry in meta-analyses of randomised controlled trials. Bmj. 2011;343

51. Balshem H, Helfand M, Schünemann HJ, et al. GRADE guidelines: 3. Rating the quality of evidence. Journal of clinical epidemiology. 2011;64(4):401–406.

52. Vanden Bossche D, Lagaert S, Willems S, Decat P. Community Health Workers as a Strategy to Tackle Psychosocial Suffering Due to Physical Distancing: A Randomized Controlled Trial. Int J Environ Res Public Health. Mar 17 2021;18(6)doi:10.3390/ijerph18063097

53. Watts A, Szabo-Reed A, Baker J, et al. LEAP! Rx: A randomized trial of a pragmatic approach to lifestyle medicine. Alzheimers Dement. Dec 2024;20(12):8374–8386. doi:10.1002/alz.14265

54. Spencer MS, Rosland AM, Kieffer EC, et al. Effectiveness of a community health worker intervention among African American and Latino adults with type 2 diabetes: a randomized controlled trial. Am J Public Health. Dec 2011;101(12):2253–60. doi:10.2105/ajph.2010.300106

55. Spencer MS, Hawkins J, Espitia NR, et al. Influence of a Community Health Worker Intervention on Mental Health Outcomes among Low-Income Latino and African American Adults with Type 2 Diabetes. Race Soc Probl. Jun 1 2013;5(2):137–146. doi:10.1007/s12552-013-9098-6

56. Rovner BW, Casten R, Chang AM, et al. Interprofessional intervention to reduce emergency department visits in Black individuals with diabetes. Population Health Management. 2023;26(1):46–52.

57. Ramirez AG, Choi BY, Munoz E, et al. Assessing the effect of patient navigator assistance for psychosocial support services on health-related quality of life in a randomized clinical trial in Latino breast, prostate, and colorectal cancer survivors. Cancer. Mar 1 2020;126(5):1112–1123. doi:10.1002/cncr.32626

58. Patel MI, Kapphahn K, Wood E, et al. Effect of a Community Health Worker-Led Intervention Among Low-Income and Minoritized Patients With Cancer: A Randomized Clinical Trial. J Clin Oncol. Feb 10 2024;42(5):518–528. doi:10.1200/jco.23.00309

59. Nyamathi AM, Salem BE, Gelberg L, et al. Pilot randomized controlled trial of biofeedback on reducing psychological and physiological stress among persons experiencing homelessness. Stress and Health. 2024;40(4):e3366.

60. Nelson K, Taylor L, Silverman J, et al. Randomized Controlled Trial of a Community Health Worker Self-Management Support Intervention Among Low-Income Adults With Diabetes, Seattle, Washington, 2010-2014. Prev Chronic Dis. Feb 9 2017;14:E15. doi:10.5888/pcd14.160344

61. Ma C, Zhou W, Tang Q, Huang S. The impact of group-based Tai chi on health-status outcomes among community-dwelling older adults with hypertension. Heart & Lung. 2018;47(4):337–344.

62. Laroche HH, Andino J, O’Shea AMJ, et al. Family-Based Motivational Interviewing and Resource Mobilization to Prevent Obesity: Living Well Together Trial. J Nutr Educ Behav. Sep 2024;56(9):631–642. doi:10.1016/j.jneb.2024.05.227

63. Kangovi S, Mitra N, Norton L, et al. Effect of Community Health Worker Support on Clinical Outcomes of Low-Income Patients Across Primary Care Facilities: A Randomized Clinical Trial. JAMA Intern Med. Dec 1 2018;178(12):1635–1643. doi:10.1001/jamainternmed.2018.4630

64. Joo JH, Xie A, Choi N, et al. A Mixed Methods Effectiveness Study of a Peer Support Intervention for Older Adults During the COVID-19 Pandemic: Results of a Randomized Clinical Trial. Am J Geriatr Psychiatry. Apr 2025;33(4):389–401. doi:10.1016/j.jagp.2024.09.013

65. Heisler M, Lapidos A, Kieffer E, et al. Impact on health care utilization and costs of a Medicaid community health worker program in Detroit, 2018–2020: a randomized program evaluation. American Journal of Public Health. 2022;112(5):766–775.

66. Gusi N, Reyes MC, Gonzalez-Guerrero JL, Herrera E, Garcia JM. Cost-utility of a walking programme for moderately depressed, obese, or overweight elderly women in primary care: a randomised controlled trial. BMC public health. 2008;8:1–10.

67. Clarke M, Clarke SJ, Jagger C. Social intervention and the elderly: a randomized controlled trial. American Journal of Epidemiology. 1992;136(12):1517–1523.

68. Carrasquillo O, Lebron C, Alonzo Y, Li H, Chang A, Kenya S. Effect of a Community Health Worker Intervention Among Latinos With Poorly Controlled Type 2 Diabetes: The Miami Healthy Heart Initiative Randomized Clinical Trial. JAMA Internal Medicine. 2017;177(7):948–954. doi:10.1001/jamainternmed.2017.0926

69. Babamoto KS, Sey KA, Camilleri AJ, Karlan VJ, Catalasan J, Morisky DE. Improving diabetes care and health measures among hispanics using community health workers: results from a randomized controlled trial. Health Education & Behavior. 2009;36(1):113–126.

70. Murphy SM, Edwards RT, Williams N, et al. An evaluation of the effectiveness and cost effectiveness of the National Exercise Referral Scheme in Wales, UK: a randomised controlled trial of a public health policy initiative. J Epidemiol Community Health. 2012;66(8):745–753.

71. McQueen A, von Nordheim D, Caburnay C, et al. A Randomized Controlled Trial Testing the Effects of a Social Needs Navigation Intervention on Health Outcomes and Healthcare Utilization among Medicaid Members with Type 2 Diabetes. Int J Environ Res Public Health. Jul 18 2024;21(7)doi:10.3390/ijerph21070936

72. Feng X, Astell-Burt T. The Pathway Domains Framework for Social Prescribing. The Lancet Primary Care. 2026:100133.

73. Wong MSJ, Low LL, Gan WH, Lee KH. A stage-sensitive approach to evaluating social prescribing in complex and evolving health systems. The Lancet Regional Health– Western Pacific. 2026;

74. Yadav UN, Wyber R, Cornforth F, Lovett RW. “Social prescribing” another stolen Indigenous concept? Medical Journal of Australia. 2024;221(6):346–346.

75. Hartig T, van den Berg AE, Hagerhall CM, et al. Health benefits of nature experience. Forests, trees and human health. Springer; 2011:127–168.

76. Sangha K, Le Brocque A, Costanza R, Cadet-James Y. Ecosystems and indigenous well-being: An integrated framework. Global Ecology and Conservation. 2015;4:197–206.

77. Arteaga-Cruz E, Cuvi J. Thinking outside the modern capitalist logic: health-care systems based in other world views. The Lancet Global Health. 2021;9(10):e1355–e1356.

78. Evers S, Kenkre J, Kloppe T, et al. Survey of general practitioners’ awareness, practice and perception of social prescribing across Europe. European Journal of General Practice. 2024;30(1):2351806.

79. Pescheny J, Randhawa G, Pappas Y. Patient uptake and adherence to social prescribing: a qualitative study. BJGP open. 2018;2(3)

80. Degerstedt F, Rapo E, Viklund EW, Jonsson F, Lundgren AS, Nilsson I. Prerequisites for social prescribing in Swedish primary care–stakeholders’ perspectives. Scandinavian Journal of Primary Health Care. 2025;43(4):776–785.

81. Cartwright L, Burns L, Akinyemi O, et al. Who is and isn’t being referred to social prescribing? National Academy for Social Prescribing https://socialprescribingacademy.org.uk/media/jaibqf4q/evidence-review-who-is-accessing-social-prescribing.pdf

82. Wilding A, Sutton M, Agboraw E, Munford L, Wilson P. Geographic inequalities in need and provision of social prescribing link workers. British Journal of General Practice. 2024;

83. Griffiths C, Jiang H, Walker K. Social Prescribing: Link Workers’ Perspectives on Service Delivery. Open Journal of Social Sciences. 2023;11(5):63–80.

84. Rhodes J, Bell S. ’’It sounded a lot simpler on the job description’’: A qualitative study exploring the role of social prescribing link workers and their training and support needs. Health & Social Care in the Community. 2021;29(6):e338–e347.

85. Sharman LS, McNamara N, Hayes S, Dingle GA. Social prescribing link workers—A qualitative Australian perspective. Health & social care in the community. 2022;30(6):e6376–e6385.

