## supplementary figures for "Effects of Social Prescribing on Mental, Physical, and Social Health Outcomes: A Systematic Review and Meta-Analysis of Randomised Trials"

Supplementary Figure 1: Forest plot for depression

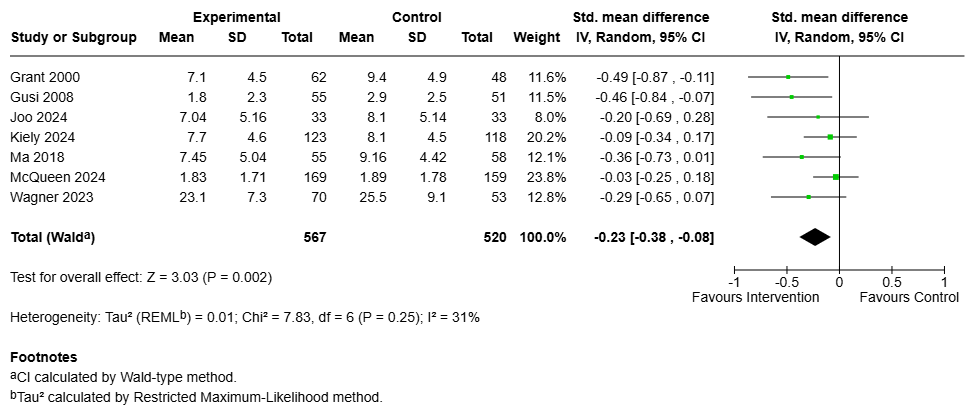

Supplementary Figure 2: Forest plot for anxiety

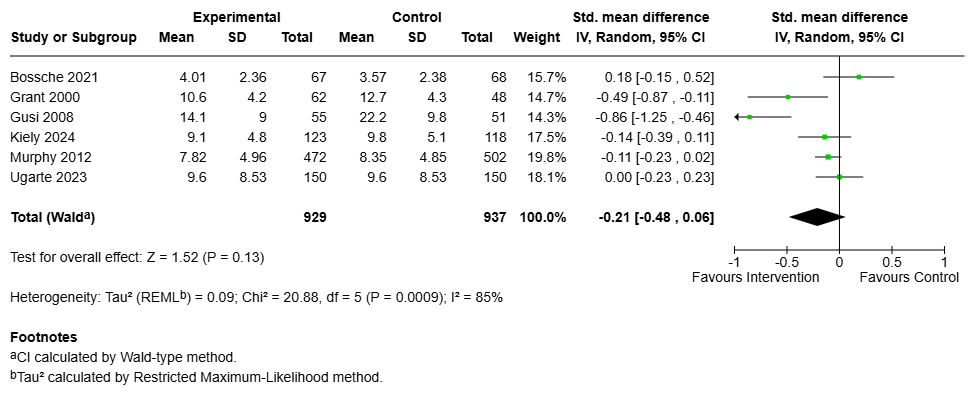

Supplementary Figure 3: Forest plot for systolic blood pressure

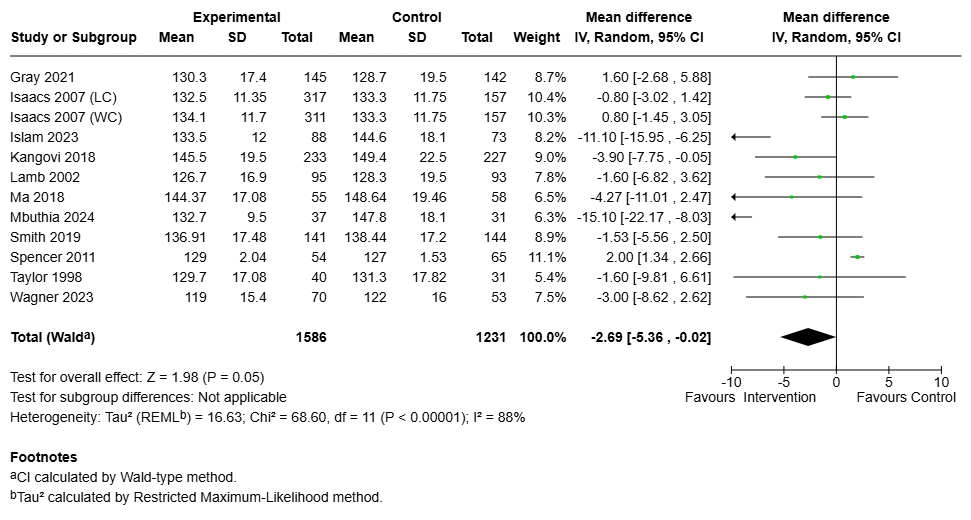

Supplementary Figure 4: Funnel plot for systolic blood pressure

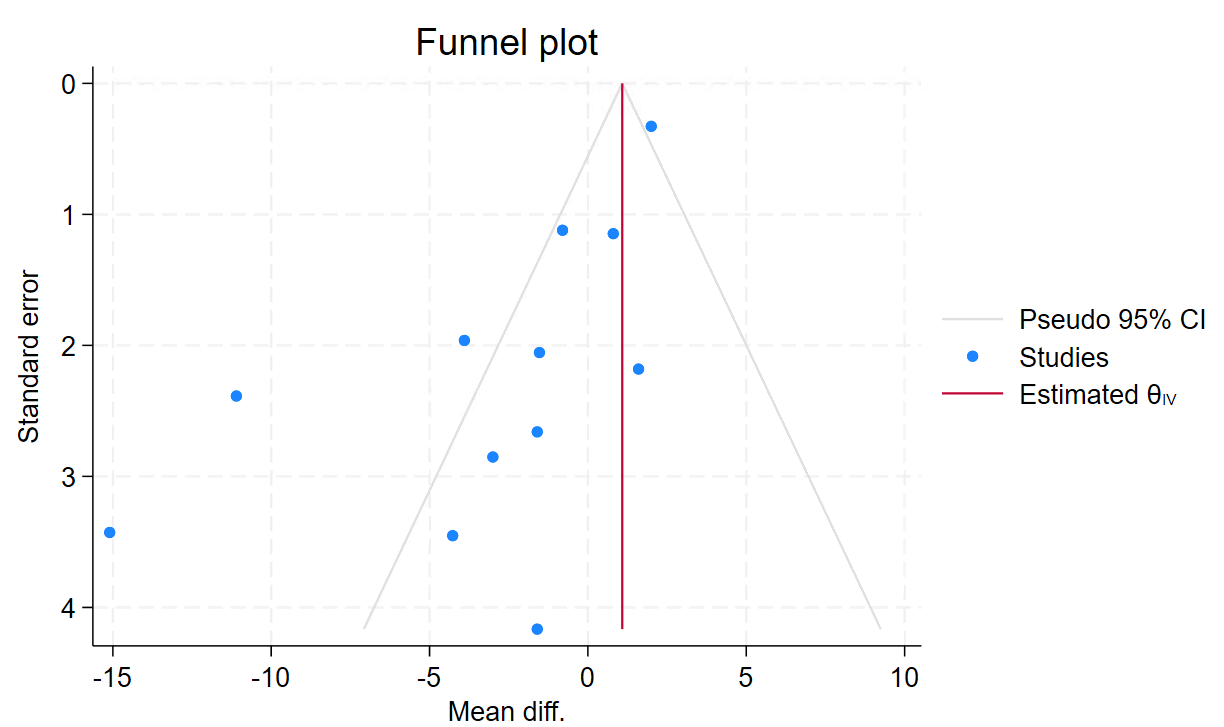

Supplementary Figure 5: Forest plot for diastolic blood pressure

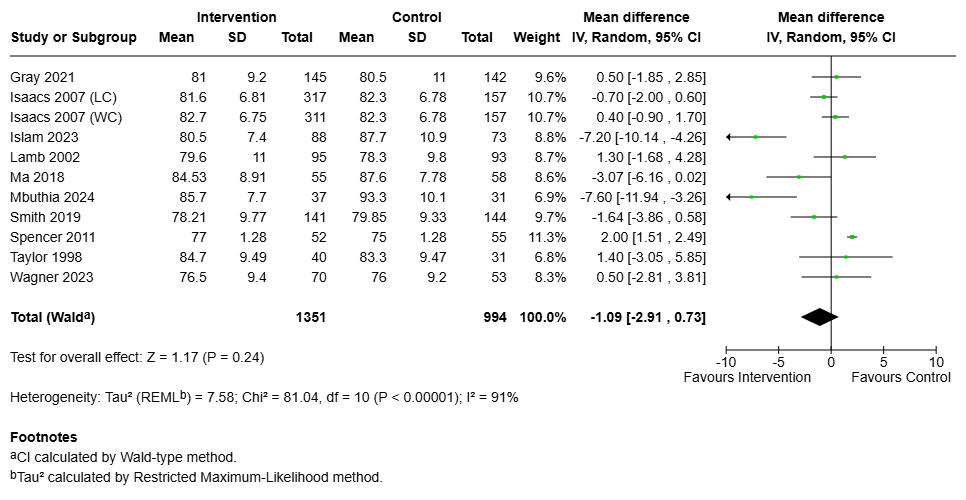

Supplementary Figure 6: Funnel plot for diastolic blood pressure

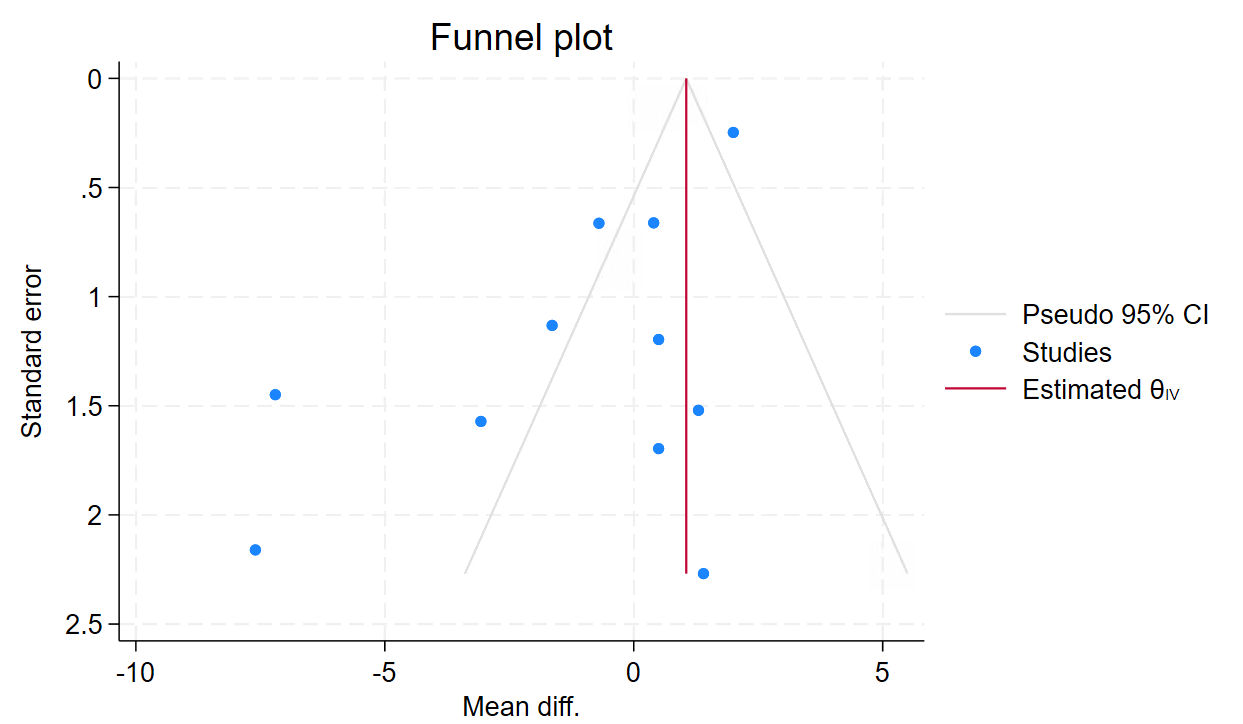

Supplementary Figure 7: Forest plot for blood pressure control

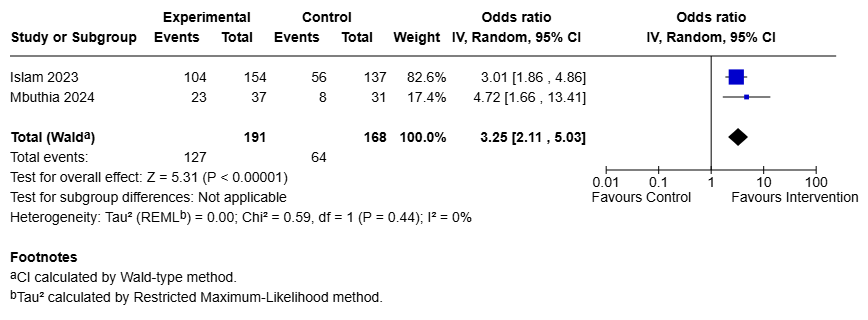

Supplementary Figure 8: Forest plot for glycated haemoglobin A1c (HbA1c)

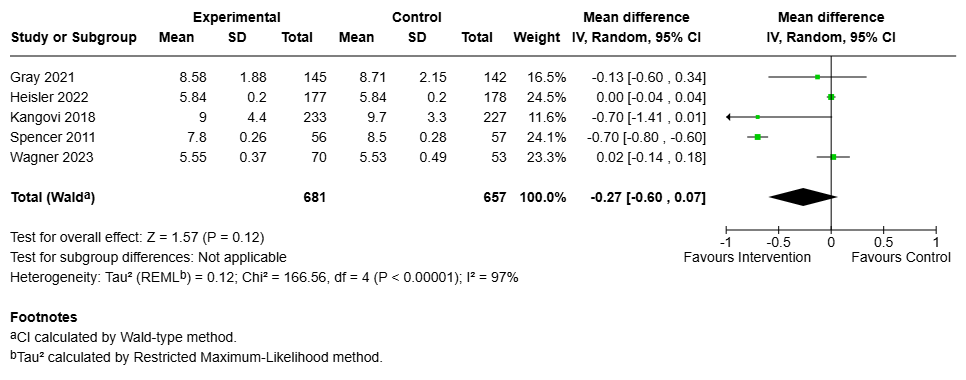

Supplementary Figure 9: Forest plot for high density lipoprotein (HDL),

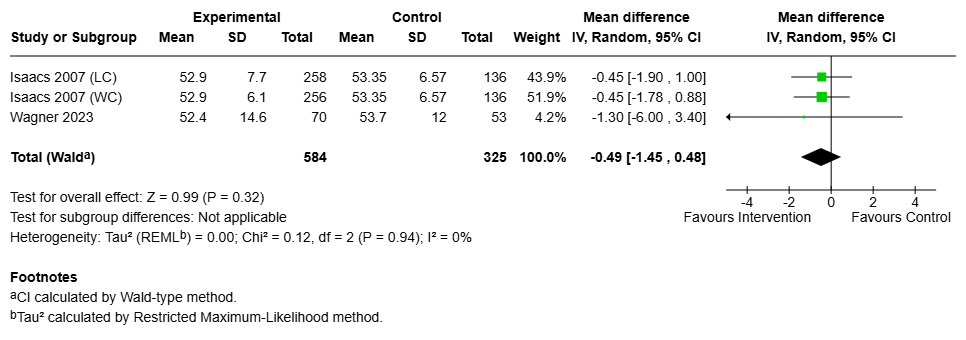

Supplementary Figure 10: Forest plot for low density lipoprotein (LDL)

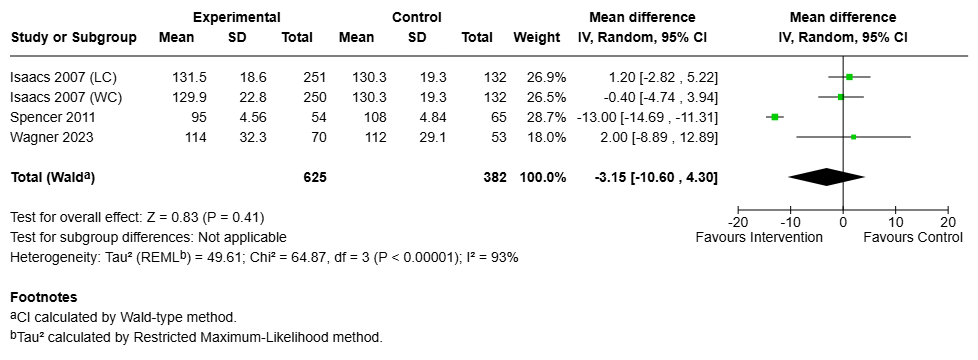

Supplementary Figure 11: Forest plot for total cholesterol (TC),

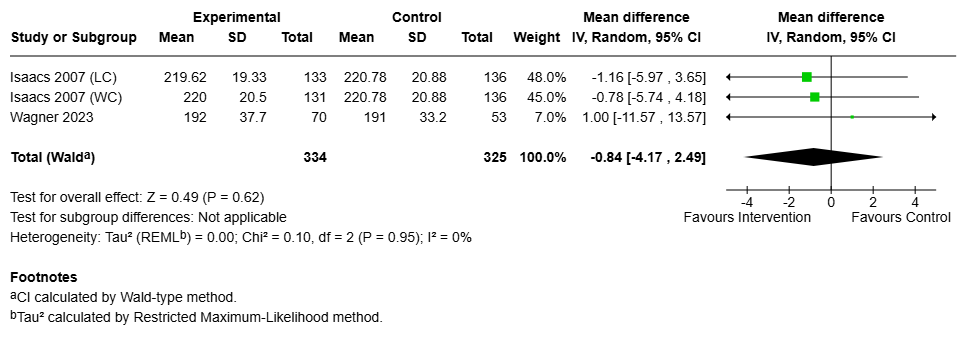

Supplementary Figure 12: Forest plot for triglycerides (TC)

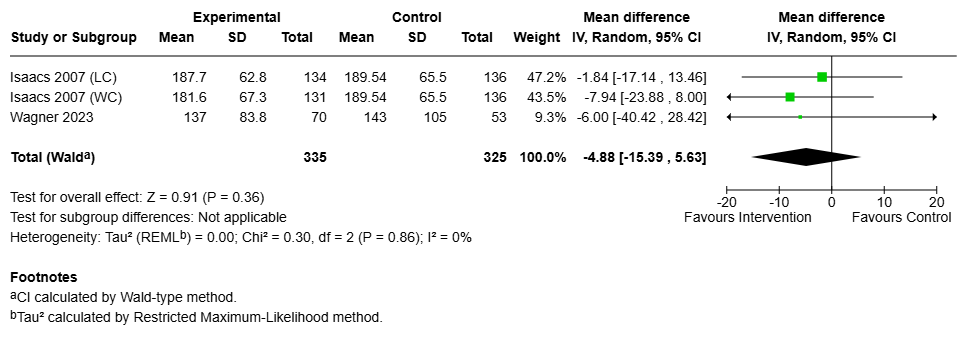

Supplementary Figure 13: Forest plot for body mass index (BMI)

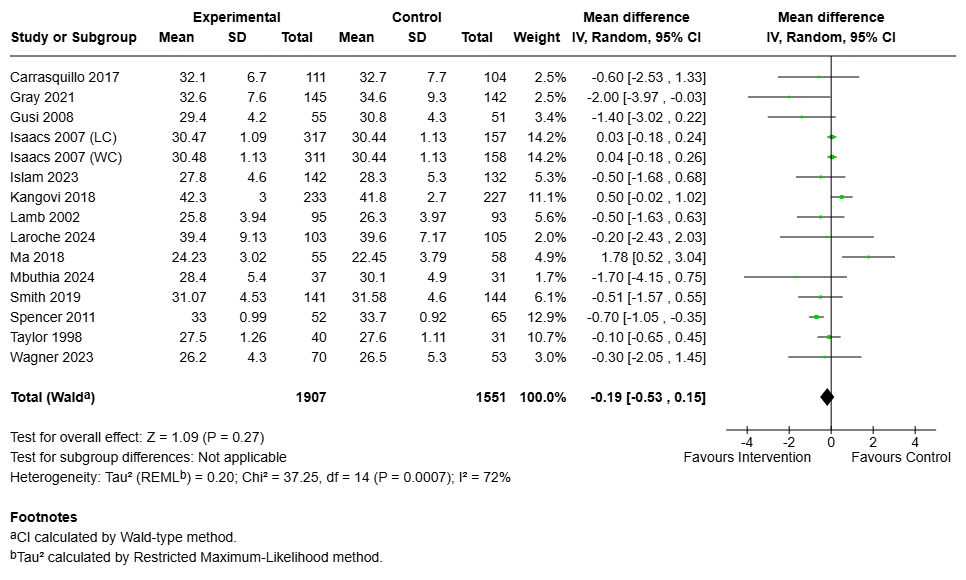

Supplementary Figure 14: Forest plot for waist circumference

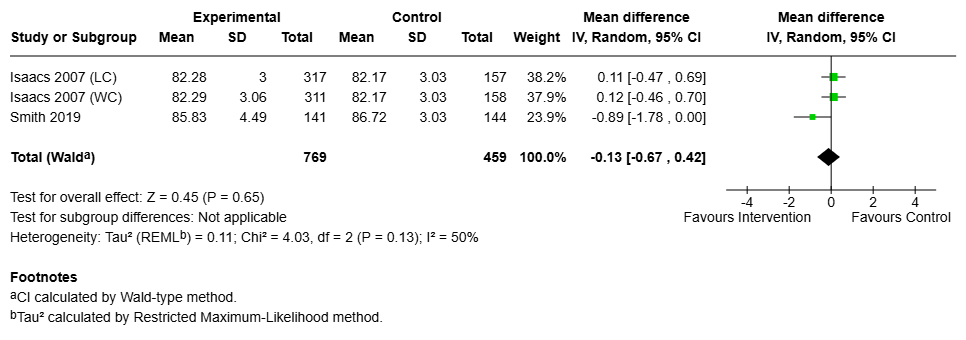

Supplementary Figure 15: Forest plot for waist-hip ratio

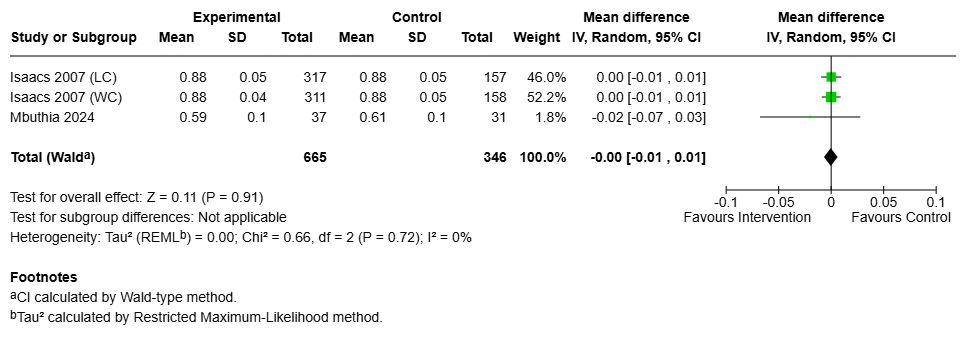

Supplementary Figure 16: Forest plot for weight

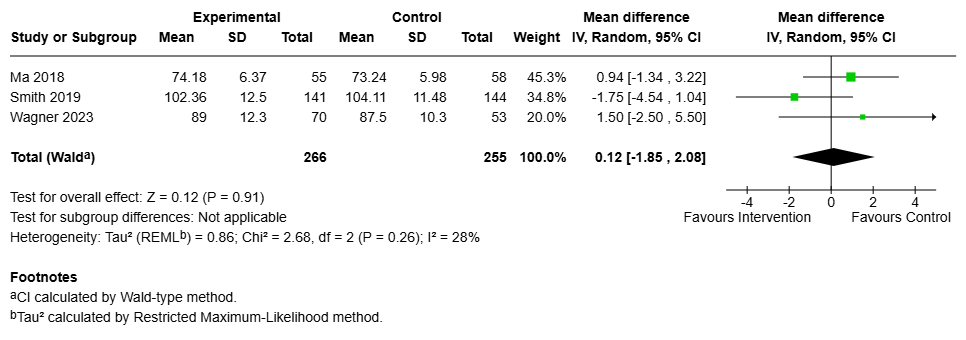

Supplementary Figure 17: Forest plot for loneliness and social isolation

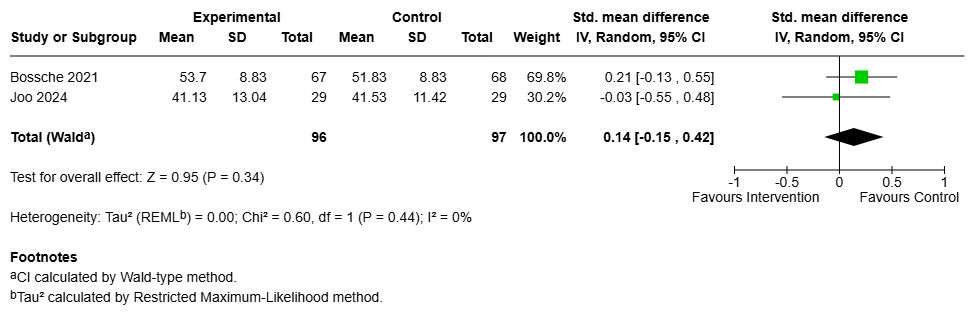

Supplementary Figure 18: Forest plot for physical activity

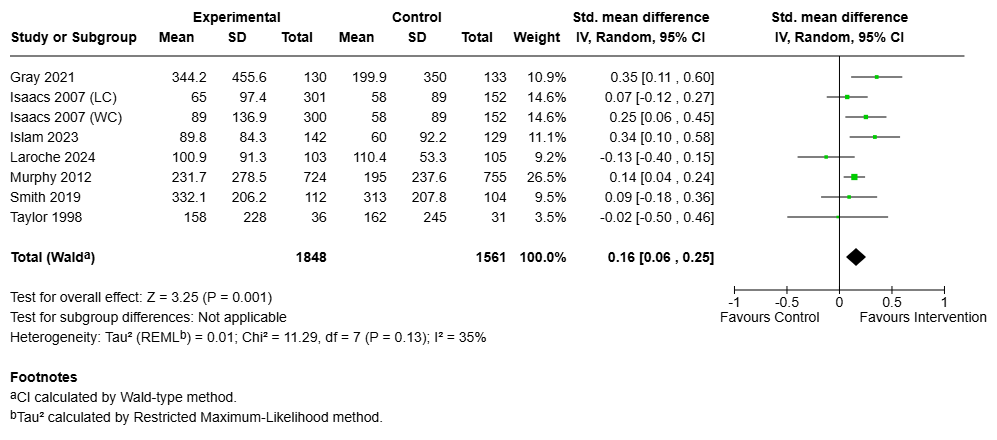

Supplementary Figure 19: Forest plot for quality of life

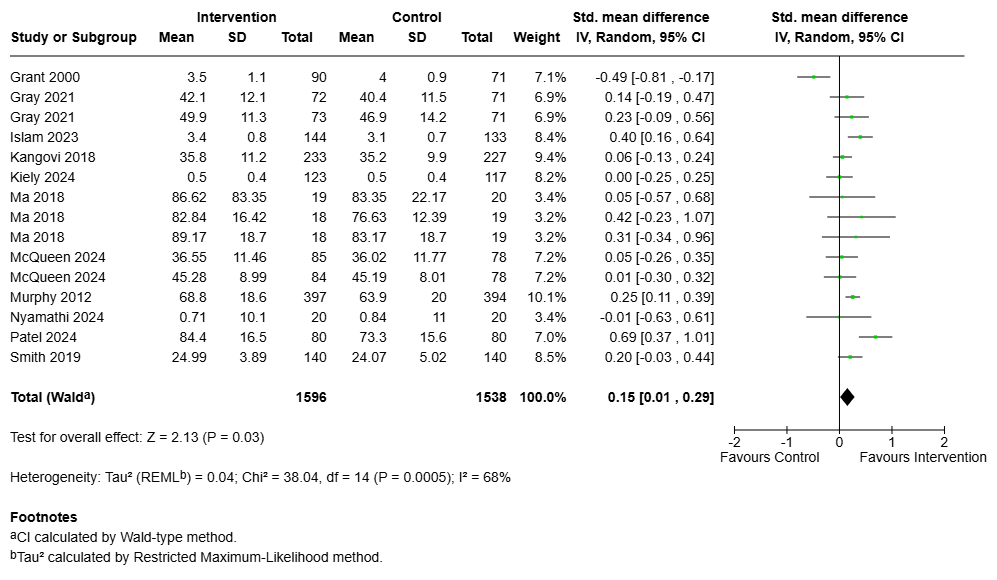

Supplementary Figure 20: Summaries of publication frequency and risk of bias

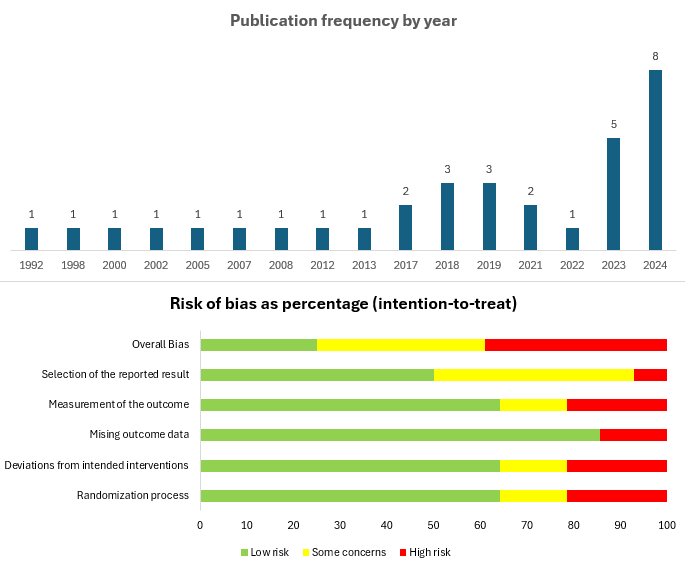

Supplementary Figure 21: Risk of bias assessment for included studies

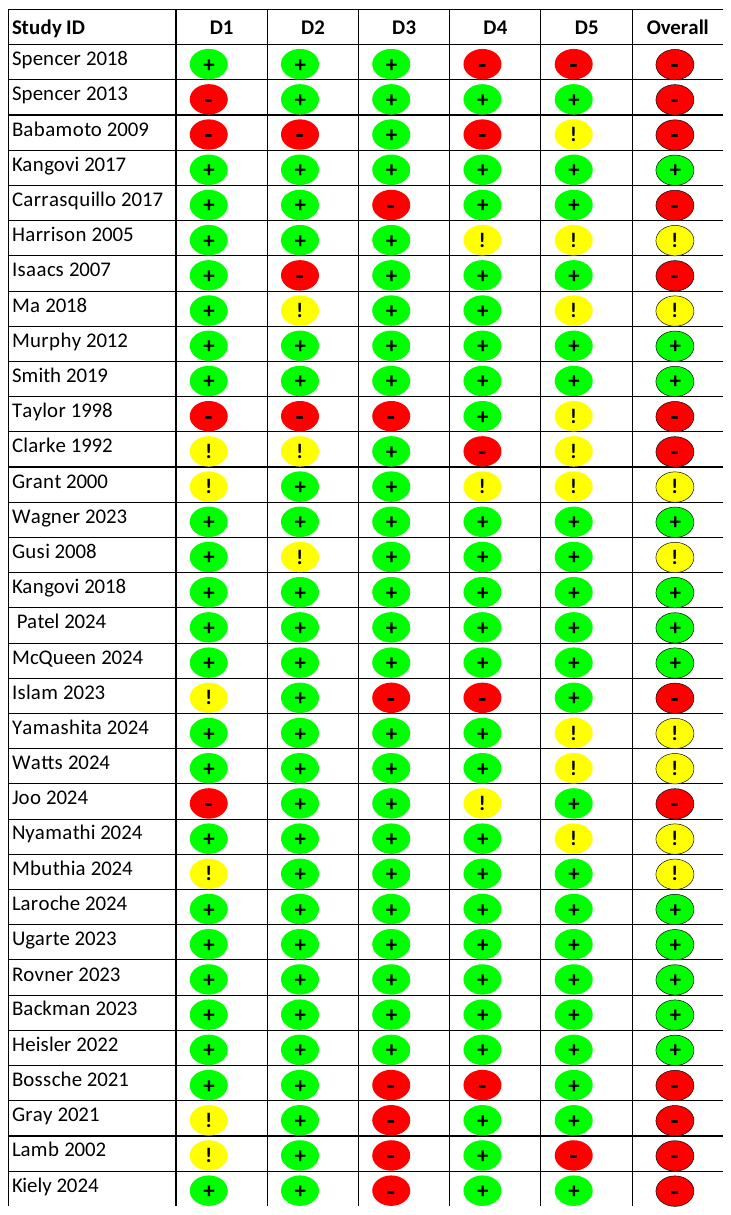

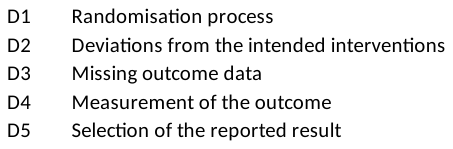
