## supplementary tables for "Effects of Social Prescribing on Mental, Physical, and Social Health Outcomes: A Systematic Review and Meta-Analysis of Randomised Trials"

**Supplementary Table 1: Search strategy for** **Ovid MEDLINE(R) ALL <1946 to November 01, 2024>**

| **#** | **Query** | **Results from 5 Nov 2024** |
| --- | --- | --- |
| 1 | ((social or non-medical or non-clinical or community) adj (prescri* or referral*)).ti. | 548 |
| 2 | ((social determinant* or social risk* or social need*) and (identif* or screen* or connect* or referral* or address* or navigat*)).ti. | 1,081 |
| 3 | social support/ or community support/ or social interaction/ | 84,070 |
| 4 | ((social* or communit*) adj3 (activ* or refer*)).mp. | 38,905 |
| 5 | ((social* adj3 prescri*) or (community adj3 prescri*)).mp. | 2,378 |
| 6 | (referral scheme* or link* scheme* or (communit* adj3 link*)).mp. | 3,590 |
| 7 | (social program* or communit* program*).mp. | 3,097 |
| 8 | (link worker* or linkworker* or communit* navigator* or health advis?r* or medical advis?r* or health trainer* or health navigator* or community health worker* or wellbeing coordinator* or well-being co-ordinator*).mp. | 13,207 |
| 9 | (randomized controlled trial or controlled clinical trial).mp. | 773,142 |
| 10 | (randomi?ed or placebo or randomly).ab. | 1,253,050 |
| 11 | clinical trials as topic.sh. | 203,720 |
| 12 | trial.ti. | 321,416 |
| 13 | or/1-8 | 142,030 |
| 14 | or/9-12 | 1,690,557 |
| 15 | 13 and 14 | 11,432 |
| 16 | exp animals/ not humans.sh. | 5,272,795 |
| 17 | 15 not 16 | 11,373 |
| 18 | limit 17 to english language | 11,128 |
| 19 | limit 18 to yr="2023 -Current" | 1,016 |

**Supplementary Table 2: Search strategy for** **CINAHL: 2023 to 29 October 2024**

| **#** | **Search terms** | **Results** |
| --- | --- | --- |
| S12 | Limit S11 to (English language, Humans, Adults, Date (2023-Oct 2024)) | 557 |
| S11 | S9 AND S10 | 20,915 |
| S10 | S6 OR S7 OR S8 | 444,298 |
| S9 | S1 OR S2 OR S3 OR S4 OR S5 | 555,267 |
| S8 | TI trial | 200,142 |
| S7 | AB (randomi?ed or placebo or randomly) | 640 |
| S6 | PT ((MH "Randomized Controlled Trials+") OR (MH "Clinical Trials+")) | 361,184 |
| S5 | "social prescribing or social prescription or community referral or link working" OR (MH "Social Inclusion") OR (MH "Social Networking+") OR (MH "Social Participation") OR (MH "Social Skills") OR (MH "Social Networks") OR (MH "Social Mobility") OR (MH "Social Alienation") OR (MH "Community Programs") | 44,340 |
| S4 | TX (social prescri*” OR referral OR pathway OR linkage) | 513,128 |
| S3 | TX (referral scheme* or link* scheme* or communit*) | 2 |
| S2 | TX (social program* or communit* program*) | 3,827 |
| S1 | TX (social support or community support or social interaction) | 152 |

**Supplementary Table 3: Search strategy for** **APA PsycINFO: 2023 to 29th October 2024**

| **#** | **Search terms** | **Results** |
| --- | --- | --- |
| 1 | ((social or non-medical or non-clinical or community) adj (prescri* or referral*)).ti. | 170 |
| 2 | ((social determinant* or social risk* or social need*) and (identif* or screen* or connect* or referral* or address* or navigat*)).ti. | 204 |
| 3 | social support/ or community support/ or social interaction/ | 75,660 |
| 4 | ((social* or communit*) adj3 (activ* or refer*)).mp. | 32,493 |
| 5 | ((social* adj3 prescri*) or (community adj3 prescri*)).mp. | 1,509 |
| 6 | (social program* or communit* program*).mp. | 4,723 |
| 7 | (link worker* or linkworker* or communit* navigator* or health advis?r* or medical advis?r* or health trainer* or health navigator* or community health worker* or wellbeing coordinator* or well-being co-ordinator*).mp. | 2,561 |
| 8 | ("social prescri*" or referral or pathway or linkage).mp. | 103,030 |
| 9 | (randomi?ed or placebo or randomly).ab. | 209,027 |
| 10 | trial.ti. | 41,874 |
| 11 | 1 or 2 or 3 or 4 or 5 or 6 or 7 or 8 | 213,926 |
| 12 | 9 or 10 | 219,926 |
| 13 | 11 and 12 | 7,876 |
| 14 | limit 13 to (human and English language and ("300 adulthood " or 320 young adulthood or 340 thirties or 360 middle age or "380 aged " or "390 very old ") and English and yr="2023 -Current") | 407 |

**Supplementary Table 4: Search strategy for** **Web of Science: 2023 to 30^th^ October 2024**

|  | | **Search terms** | **Results** |
| --- | --- | --- | --- |
| 1 | ((social or non-medical or non-clinical or community) adj (prescri* or referral*)).ti. (All Fields) | | 248 |
| 2 | ((social determinant* or social risk* or social need*) and (identif* or screen* or connect* or referral* or address* or navigat*)).ti. (All Fields) | | 1,003 |
| 3 | social support or community support or social interaction (All Fields) | | 1,502,834 |
| 4 | (social program* or communit* program*).mp. (All Fields) | | 12,506 |
| 5 | (link worker* or linkworker* or communit* navigator* or health advis?r* or medical advis?r* or health trainer* or health navigator* or community health worker* or wellbeing coordinator* or well-being co-ordinator*).mp. (All Fields) | | 2,427 |
| 6 | ((“social prescri*” OR referral OR pathway OR linkage)).mp (All Fields) | | 24,152 |
| 7 | (randomized controlled trial or controlled clinical trial).pt. (All Fields) | | 4,245 |
| 8 | (randomi?ed or placebo or randomly).ab. (All Fields) | | 38,441 |
| 9 | trial.ti. (All Fields) | | 2 |
| 10 | #1 OR #2 OR #3 OR #4 OR #5 OR #6 | | 1,535,124 |
| 11 | #9 OR #7 OR #8 | | 42,545 |
| 12 | #11 AND #10 | | 2,567 |
| 13 | #11 AND #10 Timespan: 2023-01-01 to 2024-10-30 | | 363 |

**Supplementary Table 5: Search strategy for** **NHS EED: 2023 to 30^th^ October 2024**

| **#** | **Search terms** | **Results** |
| --- | --- | --- |
| 1 | (social support) OR (community support) OR (social interaction) | 582 |
| 2 | (social prescribing) | 1 |
| 3 | (link worker) OR (linkworker) OR (communit* navigator*) | 2 |
| 4 | (referral scheme) OR (link scheme) | 8 |
| 5 | (social program) OR (community program) | 1 |
| 6 | (social or non-medical or non-clinical or community):TI | 1392 |
| 7 | (social determinant* or social risk* or social need*):TI AND (identif* or screen* or connect* or referral* or address* or navigat*):TI | 1 |
| 8 | MeSH DESCRIPTOR Randomized Controlled Trials as Topic EXPLODE ALL TREES | 12072 |
| 9 | (trial):TI | 2253 |
| 10 | #1 OR #2 OR #3 OR #4 OR #5 OR #6 OR #7 | 1900 |
| 11 | #8 OR #9 | 14111 |
| 12 | #10 AND #11 | 277 |
| 13 | (#12) FROM 2023 TO 2024 | 0 |
| 14 | (#12) WHERE LPD FROM 01/01/2023 TO 30/10/2024 | 0 |

**Supplementary Table 6: Search strategy for** **CEA Registry: 2023 to 30^th^ October 2024**

| **#** | **Search terms** | **Results** |
| --- | --- | --- |
| 1 | social program | 1 |
| 2 | community program | 4 |
| 3 | social | 14 |
| 4 | community | 17 |

**Supplementary Table 7: Search strategy for** **AMED database search strategy:**

| **#** | **Query** | **Results from 5 Nov 2024** |
| --- | --- | --- |
| 1 | ((social or non-medical or non-clinical or community) adj (prescri* or referral*)).tw. | 31 |
| 2 | ((social determinant* or social risk* or social need*) and (identif* or screen* or connect* or referral* or address* or navigat*)).ab. | 54 |
| 3 | social support/ or community support/ or social interaction.tw. | 3,009 |
| 4 | ((social* or communit*) adj3 (activ* or refer*)).tw. | 1,516 |
| 5 | ((social* adj3 prescri*) or (community adj3 prescri*)).tw. | 47 |
| 6 | (referral scheme* or link* scheme* or (communit* adj3 link*)).tw. | 82 |
| 7 | (social program* or communit* program*).tw. | 66 |
| 8 | (link worker* or linkworker* or communit* navigator* or health advis?r* or medical advis?r* or health trainer* or health navigator* or community health worker* or wellbeing coordinator* or well-being co-ordinator*).tw. | 32 |
| 9 | or/1-8 | 4,656 |
| 10 | (randomized controlled trial or controlled clinical trial).pt. | 6,141 |
| 11 | (randomi?ed or placebo or randomly).ab. | 22,082 |
| 12 | clinical trial*.tw. | 7,757 |
| 13 | trial.tw. | 14,846 |
| 14 | or/10-13 | 30,340 |
| 15 | 9 and 14 | 353 |
| 16 | limit 15 to English | 349 |

**Supplementary Table 8: Search strategy for** **Embase Classic+Embase <1947 to 2024 November 01>**
**Search Strategy:**
**1** ((social or non-medical or non-clinical or community) adj (prescri* or referral*)).ti.
**2** ((social determinant* or social risk* or social need*) and (identif* or screen* or connect* or referral* or address* or navigat*)).ti.
**3** social support/ or community support/ or social interaction/
**4** ((social* or communit*) adj3 (activ* or refer*)).mp.
**5** ((social* adj3 prescri*) or (community adj3 prescri*)).mp.
**6** (referral scheme* or link* scheme* or (communit* adj3 link*)).mp.
**7** (social program* or communit* program*).mp.
**8** (link worker* or linkworker* or communit* navigator* or health advis?r* or medical advis?r* or health trainer* or health navigator* or community health worker* or wellbeing coordinator* or well-being co-ordinator*).mp.
**9** (randomized controlled trial or controlled clinical trial).mp.
**10** (randomi?ed or placebo or randomly).ab.
 **11** clinical trials as topic.sh.
**12** trial.ti.
 **13** or/1-8
 **14** or/9-12
**15** 13 and 14
**16** limit 15 to (English language and humans)
 **17** limit 16 to yr="2023 -Current"
**18** limit 17 to (adult or aged)

**Supplementary Table 9: All search results**

| MEDLINE | 1016 |
| --- | --- |
| EMBASE | 2199 |
| CENTRAL | 757 |
| AMED | 349 |
| PsycInfo | 407 |
| Web of Science | 363 |
| CINAHL | 557 |
| NHS EED | 0 |
| CEA | 0 |
| Clinicaltrials.gov | 116 |
| WHO clinical trials registry | 24 |
| Open gray | 0 |

Supplementary Table 10. Eligibility criteria for study selection

| **Criteria** | **Inclusion** | **Exclusion** |
| --- | --- | --- |
| **Participants** | Adults with any non-communicable diseases | Children adolescents, communicable diseases, people with severe mental disorders |
| **Intervention** | Social prescribing intervention | Interventions without link worker involvement |
| **Control** | Active control, standard case, usual care, waitlist control | NA |
| **Outcomes** | Disease specific outcome measures, mental health, quality of life, anthropometrics, and cost | NA |
| **Study design** | Randomised controlled trials | Pre-post study designs, cluster-randomised controlled trials, observational studies |

Supplementary Table 11. Excluded studies

| **Authors** | **Title** | **Reason for exclusion** |
| --- | --- | --- |
| Slavin, M.; Black, C. | Adaptation of a Digital Intervention to Reduce Substance Use and Increase Contraceptive Access Among Women Involved in Criminal Legal Systems | Conference abstract/presentation |
| Loucks, Eric B.; Kronish, Ian M.; Saadeh, Frances B.; Scarpaci, Matthew M.; Proulx, Jeffrey A.; Gutman, Roee; Britton, Willoughby B.; Schuman-Olivier, Zev | Adapted Mindfulness Training for Interoception and Adherence to the DASH Diet: A Phase 2 Randomized Clinical Trial | Wrong intervention |
| Lambert, J.; Taylor, A.; Streeter, A.; Greaves, C.; Ingram, W. M.; Dean, S.; Jolly, K.; Mutrie, N.; Price, L.; Campbell, J. | Adding web-based support to exercise referral schemes improves symptoms of depression in people with elevated depressive symptoms: A secondary analysis of the e-coachER randomised controlled trial | Wrong intervention |
| Tagliaferri, S. D.; Belavy, D. L.; Bowe, S. J.; Clarkson, M. J.; Connell, D.; Craige, E. A.; Gollan, R.; Main, L. C.; Miller, C. T.; Mitchell, U. H.; Mundell, N. L.; Neason, C.; Samanna, C. L.; Scott, D.; Tait, J. L.; Vincent, G. E.; Owen, P. J. | Assessing safety and treatment efficacy of running on intervertebral discs (ASTEROID) in adults with chronic low back pain: Protocol for a randomised controlled trial | Protocol |
| Valenstein M, Pfeiffer PN, Brandfon S, Walters H, Ganoczy D, Kim HM, Cohen JL, Benn-Burton W, Carroll E, Henry J, Garcia E. | Augmenting ongoing depression care with a mutual peer support intervention versus self-help materials alone: A randomized trial | Wrong intervention |
| Pocock, M. J. O.; Hamlin, I.; Christelow, J.; Passmore, H. A.; Richardson, M. | The benefits of citizen science and nature-noticing activities for well-being, nature connectedness and pro-nature conservation behaviours | Wrong patient population |
| Rebelo, P.; Brooks, D.; Cravo, J.; Mendes, M. A.; Oliveira, A. C.; Rijo, A. S.; Moura, M. J.; Marques, A. | Beyond pulmonary rehabilitation: can the PICk UP programme fill the gap? A randomised trial in COPD | Wrong intervention |
| Kate, M. P.; Samuel, C.; Singh, S.; Jain, M.; Kamra, D.; Singh, G. B.; Sharma, M.; Pandian, J. D. | Community health volunteer for blood pressure control in rural people with stroke in India: Pilot randomised trial | Wrong intervention |
| Kitzman, H.; Dodgen, L.; Vargas, C.; Khan, M.; Montgomery, A.; Patel, M.; Ajoku, B.; Allison, P.; Strauss, A. M.; Bowen, M. | Community health worker navigation to improve allostatic load: The Integrated Population Health (IPOP) study | Wrong patient population |
| Carter, Nd; Khan, Km; McKay, Ha; Petit, Ma; Waterman, C; Heinonen, A; Janssen, Pa; Donaldson, Mg; Mallinson, A; Riddell, L; Kruse, K; Prior, Jc; Flicker, L | Community-based exercise program reduces risk factors for falls in 65- to 75-year-old women with osteoporosis: randomized controlled trial | Wrong intervention |
| Ciechanowski, P; Wagner, E; Schmaling, K; Schwartz, S; Williams, B; Diehr, P; Kulzer, J; Gray, S; Collier, C; LoGerfo, J | Community-integrated home-based depression treatment in older adults: a randomized controlled trial | Wrong intervention |
| Peretro, G.; Ballico, A. L.; Avelar, N. C. D.; Haupenthal, D. P. D. S.; Arcencio, L.; Haupenthal, A. | Comparison of aquatic physiotherapy and therapeutic exercise in patients with chronic low back pain | Wrong intervention |
| Nikoletou, D.; Chis Ster, I.; Lech, C. Y.; MacNaughton, I. S.; Chua, F.; Aul, R.; Jones, P. W. | Comparison of high-intensity interval training versus moderate-intensity continuous training in pulmonary rehabilitation for interstitial lung disease: a randomised controlled pilot feasibility trial | Wrong intervention |
| Dingle, G. A.; Sharman, L. S.; Hayes, S.; Haslam, C.; Cruwys, T.; Jetten, J.; Haslam, S. A.; McNamara, N.; Chua, D.; Baker, J. R.; Johnson, T. | A controlled evaluation of social prescribing on loneliness for adults in Queensland: 8-week outcomes | Non-RCT |
| Chen, Brian K.; Dunsiger, Shira I.; Pinto, Bernardine M. | Cost-effectiveness of peer-delivered physical activity promotion and maintenance programs for initially sedentary breast cancer survivors | Wrong intervention |
| Shah, M. K.; Wyatt, L. C.; Gibbs-Tewary, C.; Zanowiak, J. M.; Mammen, S.; Islam, N. | A Culturally Adapted, Telehealth, Community Health Worker Intervention on Blood Pressure Control among South Asian Immigrants with Type II Diabetes: Results from the DREAM Atlanta Intervention | Wrong intervention |
| Ostojic, K.; Karem, I.; Dee-Price, B. J.; Paget, S. P.; Berg, A.; Burnett, H.; Scott, T. R.; Strnadova, I.; Woolfenden, S. R. | Development of a new social prescribing intervention for families of children with cerebral palsy | Wrong patient population |
| Agarwal, Anish K.; Southwick, Lauren; Gonzales, Rachel E.; Bellini, Lisa M.; Asch, David A.; Shea, Judy A.; Mitra, Nandita; Yang, Lin; Josephs, Michael; Kopinksy, Michael; Kishton, Rachel; Balachandran, Mohan; Benjamin Wolk, Courtney; Becker-Haimes, Emily M.; Merchant, Raina M. | Digital Engagement Strategy and Health Care Worker Mental Health: A Randomized Clinical Trial | Wrong patient population |
| Whitehouse, C. R.; Knowles, M.; Long, J. A.; Mitra, N.; Volpp, K. G.; Xu, C.; Sabini, C.; Gerald, N.; Estrada, I.; Jones, D.; Kangovi, S. | Digital Health and Community Health Worker Support for Diabetes Management: a Randomized Controlled Trial | Wrong intervention |
| Yeo, GeckHong; Loo, Gladys; Oon, Matt; Pang, Rachel; Ho, Dean | A digital peer support platform to translate online peer support for emerging adult mental well-being: Randomized controlled trial | Wrong patient population |
| Azadbakht, Mojtaba; Tanjani, Parisa Taheri; Fadayevatan, Reza; Foroughan, Mahshid; Zanjari, Nasibeh | The Effect of a Peer Social Support Network Intervention on Self-management of the Elderly With Type 2 Diabetes | Wrong intervention |
| Quansah, D. Y.; Gilbert, L.; Arhab, A.; Gonzalez-Rodriguez, E.; Hans, D.; Gross, J.; Lanzi, S.; Stuijfzand, B.; Lacroix, A.; Horsch, A.; Puder, J. J. | Effect of a prepartum and postpartum, complex interdisciplinary lifestyle and psychosocial intervention on metabolic and mental health outcomes in women with gestational diabetes mellitus (the MySweetheart trial): randomised, single centred, blinded, controlled trial | Wrong intervention |
| Petruseviciene, D; Surmaitiene, D; Baltaduoniene, D; Lendraitiene, E | Effect of community-based occupational therapy on health-related quality of life and engagement in meaningful activities of women with breast cancer | Wrong intervention |
| Balci, H.; Faydali, S. | The Effect of Education Performed Using Mobile Application on Supportive Care Needs and Quality of Life in Women with Breast Cancer: Randomized Controlled Trial | Wrong intervention |
| Van Uytsel, H.; Ameye, L.; Devlieger, R.; Bijlholt, M.; Van der Gucht, K.; Jacquemyn, Y.; Bogaerts, A. | Effect of the INTER-ACT lifestyle intervention on maternal mental health during the first year after childbirth: A randomized controlled trial | Wrong intervention |
| Torri A, Volpato E, Merati G, Milani M, Toccafondi A, Formenti D, La Rosa F, Agostini S, Agliardi C, Oreni L, Sacco A | EffectiVenEss of a Rehabilitation Treatment With Nordic Walking in obEse or oveRweight Diabetic patiEnts With Cardiovascular Disease. The VENERE Study | Protocol |
| Wong, M. Y. C.; Leung, K. M.; Thogersen-Ntoumani, C.; Ou, K.; Chung, P. K. | Effectiveness of a supervised group-based walking program on physical, psychological and social outcomes among older adults: a randomised controlled trial protocol | Protocol |
| Chan, Aw; Lee, A; Suen, Lk; Tam, Ww | Effectiveness of a Tai chi Qigong program in promoting health-related quality of life and perceived social support in chronic obstructive pulmonary disease clients | Wrong intervention |
| Tavares, V. D. D.; Schuch, F. B.; de Sousa, G. M.; Hallgren, M.; Neto, L. O.; Cabral, D. A. R.; de Almeida, R. N.; Barbosa, D. C.; de Almeida, V. R. N.; Tinoco, H.; Lira, R. A.; Hallak, J. E.; Arcoverde, E.; Cuthbert, C.; Patten, S.; Galvao-Coelho, N. L. | Effectiveness of an affect-adjusted, supervised, multimodal, online and home-based exercise group protocol for major depression: A randomized controlled trial | Wrong intervention |
| Kim, I.; An, H.; Yun, S.; Park, H. Y. | Effectiveness of community-based interventions for older adults living alone: a systematic review and meta-analysis | Review |
| Htun, H. L.; Teshale, A. B.; Cumpston, M. S.; Demos, L.; Ryan, J.; Owen, A.; Freak-Poli, R. | Effectiveness of social prescribing for chronic disease prevention in adults: a systematic review and meta-analysis of randomised controlled trials | Review |
| O'Sullivan, D. J.; Bearne, L. M.; Harrington, J. M.; Cardoso, J. R.; McVeigh, J. G. | The effectiveness of social prescribing in the management of long-term conditions in community-based adults: A systematic review and meta-analysis | Review |
| Jensen, M. T.; Nielsen, S. S.; Jessen-Winge, C.; Madsen, C. M. T.; Thilsing, T.; Larrabee Sonderlund, A.; Christensen, J. R. | The effectiveness of social-support-based weight-loss interventions-a systematic review and meta-analysis | Review |
| Wong MY, Leung KM, Thøgersen-Ntoumani C, Ou K, Chung PK. | The Effectiveness of Supervised Group-based Walking Program on Physical, Psychological and Social Outcomes Among Older Adults: A Randomized Controlled Trial | protocol |
| Wong, A. K. C.; Tso, W. C.; Su, J. J.; Hui, V. C. C.; Chow, K. K. S.; Wong, S. M.; Wong, B. B.; Wong, F. K. Y. | Effectiveness of support from community health workers on the sustained use of a wearable monitoring device among community-dwelling older adults: A randomized trial protocol | protocol |
| Litt, J. S.; Alaimo, K.; Harrall, K. K.; Hamman, R. F.; Hebert, J. R.; Hurley, T. G.; Leiferman, J. A.; Li, K.; Villalobos, A.; Coringrato, E.; Courtney, J. B.; Payton, M.; Glueck, D. H. | Effects of a community gardening intervention on diet, physical activity, and anthropometry outcomes in the USA (CAPS): an observer-blind, randomised controlled trial | Wrong patient population |
| Kawaguchi, K.; Nakagomi, A.; Ide, K.; Kondo, K. | Effects of a Mobile App to Promote Social Participation on Older Adults: Randomized Controlled Trial | Wrong patient population |
| Talevski, J.; Gianoudis, J.; Bailey, C. A.; Ebeling, P. R.; Nowson, C. A.; Hill, K. D.; Sanders, K. M.; Daly, R. M. | Effects of an 18-month community-based, multifaceted, exercise program on patient-reported outcomes in older adults at risk of fracture: secondary analysis of a randomised controlled trial | Wrong intervention |
| Ye, Y.; Wan, M. Y.; Lin, H. Y.; Xia, R.; He, J. Q.; Qiu, P. T.; Zheng, G. H. | Effects of Baduanjin exercise on cognitive frailty, oxidative stress, and chronic inflammation in older adults with cognitive frailty: a randomized controlled trial | Wrong intervention |
| Zhao, X.; Sha, X.; Qi, L. | Effects of comprehensive exercise training on frailtynegative emotions and physical functions of elderly patients with diabetes | Wrong intervention |
| Sanchez-Alcala, M.; Aibar-Almazan, A.; Hita-Contreras, F.; Castellote-Caballero, Y.; Carcelen-Fraile, M. D. C.; Infante-Guedes, A.; Gonzalez-Martin, A. M. | Effects of Dance-Based Aerobic Training on Mental Health and Quality of Life in Older Adults with Mild Cognitive Impairment | Wrong intervention |
| Depenbusch, J.; Hiensch, A. E.; Schmidt, M.; Monninkhof, E. M.; Puente, M. P.; Clauss, D.; Zimmer, P.; Belloso, J.; Trevaskis, M.; Rundqvist, H.; Wiskemann, J.; Muller, J.; Fremd, C.; Altena, R.; Kufel-Grabowska, J.; Bijlsma, R. M.; Van Leeuwen-Snoeks, L.; Bokkel-Huinink, D. T.; Sonke, G.; Mann, B.; Francis, P.; Richardson, G.; Malter, W.; Lopez, I. M. A.; Van Der Wall, E.; Aaronson, N.; Senkus, E.; Urruticoechea, A.; Zopf, E.; Bloch, W.; Stuiver, M. M.; Wengstrom, Y.; May, A.; Steindorf, K. | Effects of exercise on fatigue and quality of life in metastatic breast cancer patients: The randomized controlled PREFERABLE-EFFECT study | Conference abstract/presentation |
| Zheng, S. S.; Zhang, H.; Zhang, M. H.; Li, X.; Chang, K.; Yang, F. C. | The effects of group-based cognitive behavioral therapy in the rehabilitation of patients with chronic schizophrenia with more than two years of community-based mental health group rehabilitation | Wrong intervention |
| Zhang, Jun; Yang, Chao; Pan, Yujie; Wang, Li | Effects of Multicomponent Exercise on Community-Dwelling Older Adults With Mild Cognitive Impairment | Wrong intervention |
| Planta, O.; Cami, M.; Matskiv, J.; Plonka, A.; Gros, A.; Beauchet, O. | Effects of museum-based art activities on older community dwellers' physical activity: the A-health randomized controlled trial results | Wrong intervention |
| Ho, A. H. Y.; Ma, S. H. X.; Tan, M. K. B.; Bajpai, R.; Goh, S. S. N.; Yeo, G.; Teng, A.; Yang, Y.; Galery, K.; Beauchet, O. | Effects of participatory 'A'rt-Based Activity On 'Health' of Older Community-Dwellers: results from a randomized control trial of the Singapore A-Health Intervention | Wrong patient population |
| Cronan, Ta; Hay, M; Groessl, E; Bigatti, S; Gallagher, R; Tomita, M | The effects of social support and education on health care costs after three years | Wrong intervention |
| Jiang, D.; Tang, V. F. Y.; Kahlon, M.; Chow, E. O. W.; Yeung, D. Y. L.; Aubrey, R.; Chou, K. L. | Effects of Wisdom-Enhancement Narrative-Therapy and Empathy-Focused interventions on loneliness over 4 weeks among older adults: A Randomized Controlled Trial | Wrong intervention |
| Rawal, S.; White, J.; Nepal, J.; Subedi, S.; Daneault, J. F.; Shakya, P.; Shrestha, A. | Efficacy of a Mobile App Intervention for Management of Gestational Diabetes: An Exploratory Randomized Controlled Trial | Conference abstract/presentation |
| Stockton, M. B.; Ward, K. D.; McClanahan, B. S.; Vander Weg, M. W.; Coday, M.; Wilson, N.; Relyea, G.; Read, M. C.; Connelly, S.; Johnson, K. C. | The Efficacy of Individualized, Community-Based Physical Activity to Aid Smoking Cessation: A Randomized Controlled Trial | Wrong intervention |
| Fiszdon, J. M.; Dixon, H. D.; Davidson, C. A.; Roberts, D. L.; Penn, D. L.; Bell, M. D. | Efficacy of social cognition and interaction training in outpatients with schizophrenia spectrum disorders: randomized controlled trial | Wrong intervention |
| Xie, Q.; Cheng, L. | The empowerment-driven app-assisted program among patients with poorly controlled type 2 diabetes | Conference abstract/presentation |
| Ostojic, K.; Karem, I.; Paget, S.; Berg, A.; Burnett, H.; Scott, T.; Martin, T.; Dee-Price, B. J.; McIntyre, S.; Smithers-Sheedy, H.; Mimmo, L.; Masi, A.; Scarcella, M.; Azmatullah, S.; Calderan, J.; Mohamed, M.; Olaso, A.; van Hoek, M.; van Hoek, D.; Woodbury, M.; Wilkinson, A.; Chambers, G.; Zwi, K.; Dale, R.; Eapen, V.; Lingam, R.; Strnadova, I.; Woolfenden, S. | EPIC-CP pilot trial study protocol: a multicentre, randomised controlled trial investigating the feasibility and acceptability of social prescribing for Australian children with cerebral palsy | Wrong patient population |
| Silva, H. J.; Miranda, J. P.; Melo, C. S.; Fonseca, L. S.; Mascarenhas, R. O.; Veloso, N. S.; Silva, W. T.; Bastone, A. C.; Oliveira, V. C. | The ESCAPE Trial for Older People With Chronic Low Back Pain: A Feasibility Study of a Clinical Trial of Group-Based Exercise in Primary Health Care | Wrong intervention |
| Mewara, N.; Chhajed, B. K. B. | Evaluating the Impact of a Community Based, Peer-Led Exercise Program on the Prevention of Osteoporotic Fractures among Elderly Women | Wrong intervention |
| Lincoln, Nb; Walker, Mf; Dixon, A; Knights, P | Evaluation of a multiprofessional community stroke team: a randomized controlled trial | Wrong patient population |
| Ward, M | Experiences in physical activity promotion in health care settings for primary prevention in the UK | Letter |
| Hernon, S. M.; Singh, Y.; Ward, N.; Kramer, A. F.; Travison, T. G.; Verghese, J.; Fielding, R. A.; Kowaleski, C.; Reid, K. F. | A feasibility randomized controlled trial of a community-level physical activity strategy for older adults with motoric cognitive risk syndrome | Wrong intervention |
| Kwiringira, A.; Migisha, R.; Bulage, L.; Kwesiga, B.; Kadobera, D.; Upenytho, G.; Mbaka, P.; Harris, J. R.; Hayes, D.; Ario, A. R. | Group-based Education and monitoring program delivered by community health workers to improve control of high blood pressure in island districts of lake victoria, Uganda | Non-RCT |
| Bantum, E. O.; Yamada, P. M.; Makolo, T.; Yu, H.; Pagano, I.; Subia, N.; Walsh, C.; Loo, L. W. M. | Hula as a physical activity and social support intervention for sustained activity in female breast and gynecologic cancer survivors | Wrong intervention |
| Wildman, John; Wildman, Josephine M. | Impact of a link worker social prescribing intervention on non-elective admitted patient care costs: A quasi-experimental study | Non-RCT |
| Carter, J. A.; Isselbacher, E.; Donelan, K.; Thorndike, A. N. | Implementing a Digitally-Enabled Community Health Worker Intervention for Patients With Heart Failure: A Randomized Controlled Trial | Conference abstract/presentation |
| Goncalves, C.; Bravo, J.; Pais, J.; Abreu, A.; Raimundo, A. | Improving Health Outcomes in Coronary Artery Disease Patients with Short-Term Protocols of High-Intensity Interval Training and Moderate-Intensity Continuous Training: A Community-Based Randomized Controlled Trial | Wrong intervention |
| Joseph, J. J.; Nolan, T. S.; Brock, G.; Williams, A.; Zhao, S.; McKoy, A.; Kluwe, B.; Metlock, F.; Campanelli, K.; Odei, J. B.; Khumalo, M. T.; Lavender, D.; Gregory, J.; Gray, D. M. | Improving mental health in black men through a 24-week community-based lifestyle change intervention: the black impact program | Non-RCT |
| Jansson, A. K.; Lubans, D. R.; Duncan, M. J.; Smith, J. J.; Bauman, A.; Attia, J.; Robards, S. L.; Cox, E. R.; Beacroft, S.; Plotnikoff, R. C. | Increasing participation in resistance training using outdoor gyms: A study protocol for the ecofit type III hybrid effectiveness implementation trial | protocol |
| Snethen, Gretchen; McCormick, Bryan P.; Nagata, Shinichi; Salzer, Mark S. | Independence Through Community Access and Navigation: A Supported Leisure Intervention for Individuals With Negative Symptoms | Wrong intervention |
| Goff, D. L.; Perraud, G.; Derriennic, J.; Aujoulat, P.; Guillou, M.; Barais, M.; Reste, J. Y. L. | Innovative population-based strategies for primary prevention of cardiovascular disease: A 2-year randomised control trial (RCT) evaluating behavioural change led by community cham-pions versus brief advice | Conference abstract/presentation |
| Lim, S.; Wyatt, L.; Belli, H.; Mammen, S.; Zanowiak, J.; Hussain, S.; Mohaimin, S.; Islam, N. | An Integrated Electronic Health Record and Community Health Worker Intervention Shows Clinically Significant Weight Loss among South Asian Patients at Risk for Diabetes | Conference abstract/presentation |
| Bostrom, C.; Dahlin, K.; Nilsson-Khelili, R.; Nasman, L.; Sarac, D. C.; Enman, Y.; Metsios, G. S.; Bucher, S. M.; Parodis, I. | Interim Results from a Randomized Controlled Trial Evaluating Aerobic High-Intensity Interval Training Combined with Resistance Training in Systemic Lupus Erythematosus | Conference abstract/presentation |
| Lim, M. H.; Hennessey, A.; Qualter, P.; Smith, B. J.; Thurston, L.; Eres, R.; Holt-Lunstad, J. | The KIND Challenge community intervention to reduce loneliness and social isolation, improve mental health, and neighbourhood relationships: an international randomized controlled trial | Wrong patient population |
| Xiang, X.; Kayser, J.; Turner, S.; Ash, S.; Himle, J. A. | Layperson-Supported, Web-Delivered Cognitive Behavioral Therapy for Depression in Older Adults: Randomized Controlled Trial | Wrong intervention |
| Peck, R. N.; Issarow, B.; Kisigo, G. A.; Kabakama, S.; Okello, E.; Rutachunzibwa, T.; Willkens, M.; Deogratias, D.; Hashim, R.; Grosskurth, H.; Fitzgerald, D. W.; Ayieko, P.; Lee, M. H.; Murphy, S. M.; Metsch, L. R.; Kapiga, S. | Linkage Case Management and Posthospitalization Outcomes in People with HIV: The Daraja Randomized Clinical Trial | Wrong intervention |
| Pinto, B. M.; Dunsiger, S. I.; DeScenza, V. R.; Stein, K. | Mediators of physical activity outcomes in a peer-led intervention for breast cancer survivors | Wrong patient population |
| Deckers, K.; Zwan, M. D.; Soons, L. M.; Waterink, L.; Beers, S.; van Houdt, S.; Stiensma, B.; Kwant, J. Z.; Wimmers, S. C. P. M.; Heutz, R. A. M.; Claassen, J. A. H. R.; Oosterman, J. M.; de Heus, R. A. A.; van de Rest, O.; Vermeiren, Y.; Voshaar, R. C. O.; Smidt, N.; Broersen, L. M.; Sikkes, S. A. M.; Aarts, E.; Kohler, S.; van der Flier, W. M. | A multidomain lifestyle intervention to maintain optimal cognitive functioning in Dutch older adults-study design and baseline characteristics of the FINGER-NL randomized controlled trial | Wrong intervention |
| Underwood, Martin; Achana, Felix; Carnes, Dawn; Eldridge, Sandra; Ellard, David R.; Griffiths, Frances; Haywood, Kirstie; Wan Hee, Siew; Higgins, Helen; Mistry, Dipesh; Mistry, Hema; Newton, Sian; Nichols, Vivien; Norman, Chloe; Padfield, Emma; Patel, Shilpa; Petrou, Stavros; Pincus, Tamar; Potter, Rachel; Sandhu, Harbinder | Non-pharmacological educational and self-management interventions for people with chronic headache: the CHESS research programme including a RCT | Non-RCT |
| Lee, S.; Bae, S.; Harada, K.; Makino, K.; Anan, Y.; Suzuki, T.; Shimada, H. | A non-pharmacological multidomain intervention of dual-task exercise and social activity affects the cognitive function in community-dwelling older adults with mild to moderate cognitive decline: A randomized controlled trial | Wrong intervention |
| Lee, S.; Harada, K.; Bae, S.; Harada, K.; Makino, K.; Anan, Y.; Suzuki, T.; Shimada, H. | A non-pharmacological multidomain intervention of dual-task exercise and social activity affects the cognitive function in community-dwelling older adults with mild to moderate cognitive decline: A randomized controlled trial | Wrong intervention |
| O'Connell, Mj; Sledge, Wh; Staeheli, M; Sells, D; Costa, M; Wieland, M; Davidson, L | Outcomes of a peer mentor intervention for persons with recurrent psychiatric hospitalization | Wrong intervention |
| Liu, S.; Cai, Y.; Yao, S.; Chai, J.; Jia, Y.; Ge, H.; Huang, R.; Li, A.; Cheng, H. | Perceived social support mediates cancer and living meaningfully intervention effects on quality of life after breast cancer surgery | Wrong intervention |
| Hao, X.; Yi, Y.; Lin, X.; Li, J.; Chen, C.; Shen, Y.; Sun, Y.; He, J. | Personalised graded psychological intervention on negative emotion and quality of life in patients with breast cancer | Wrong intervention |
| Burdett Trust for, Nursing; Nottinghamshire Healthcare, N. H. S. Trust; Nottingham City Primary Care, Trust; Nottinghamshire County Teaching Primary Care, Trust | Pragmatic Randomised Controlled Trial of a Preferred Intensity Exercise Programme to Improve Physiological and Associated Psychological, Social, and Wellbeing Outcomes of Women Living With Depression | Wrong intervention |
| University of California, Berkeley; University of California, San Diego; National Institute of, Diabetes; Digestive,; Kidney, Diseases | PRescription Exercise for Older Men With Urinary Disease (PROUD) Pilot Study | protocol |
| Coringrato, E.; Alaimo, K.; Leiferman, J. A.; Villalobos, A.; Buchenau, H.; Decker, E.; Fahnestock, L.; Quist, P.; Litt, J. S. | A process evaluation of a randomized-controlled trial of community gardening to improve health behaviors and reduce stress and anxiety | Wrong intervention |
| Saha, C. K.; Shubrook, J. H.; Guyton Hornsby, W.; Yang, Z.; Pillay, Y.; Mather, K. J.; de Groot, M. | Program ACTIVE II: 6- and 12-month outcomes of a treatment approach for major depressive disorder in adults with type 2 diabetes | Wrong intervention |
| Katzow, M. W.; Messito, M. J.; Mendelsohn, A. L.; Scott, M. A.; Gross, R. S. | Protective Effect of Prenatal Social Support on the Intergenerational Transmission of Obesity in Low-Income Hispanic Families | Wrong patient population |
| Clark, Ms; Rubenach, S; Winsor, A | A randomized controlled trial of an education and counselling intervention for families after stroke | Wrong intervention |
| Bomyea, J.; Sweet, A.; Davey, D. K.; Boland, M.; Paulus, M. P.; Stein, M. B.; Taylor, C. T. | Randomized controlled trial of computerized approach/avoidance training in social anxiety disorder: Neural and symptom outcomes | Wrong intervention |
| Shorey, S; Chan, Sw; Chong, Ys; He, Hg | A randomized controlled trial of the effectiveness of a postnatal psychoeducation programme on self-efficacy, social support and postnatal depression among primiparas | Wrong intervention |
| Houlihan, Bethlyn Vergo; Brody, Miriam; Everhart-Skeels, Sarah; Pernigotti, Diana; Burnett, Sam; Zazula, Judi; Green, Christa; Hasiotis, Stathis; Belliveau, Timothy; Seetharama, Subramani; Rosenblum, David; Jette, Alan | Randomized Trial of a Peer-Led, Telephone-Based Empowerment Intervention for Persons With Chronic Spinal Cord Injury Improves Health Self-Management | Conference abstract/presentation |
| Smith, J; Forster, A; Young, J | A randomized trial to evaluate an education programme for patients and carers after stroke | Wrong intervention |
| Greysen, S. R.; Turner, G.; Oon, A. L.; Harkins, K.; Karlawish, J. H. | An Rct of Gamification with Social Support to Increase Physical Activity in Older Adults at Risk for Alzheimer's: The Step 4 Life Trial | Conference abstract/presentation |
| Barcelona Institute of Global, Health; Helsinki City Older People's, Services; The Finnish Association for the Welfare of Older, People | RECETAS (Re-Imagining Environments for Connection and Engagement: Testing Actions for Social Prescribing in Natural Spaces) in Helsinki | Non-RCT |
| Alegria, M.; Cruz-Gonzalez, M.; Markle, S. L.; Falgas-Bague, I.; Poindexter, C.; Stein, G. L.; Eddington, K.; Martinez Vargas, A. E.; Fuentes, L.; Cheng, M.; Shrout, P. E. | Referrals to Community and State Agencies to Address Social Determinants of Health for Improving Mental Health, Functioning, and Quality of Care Outcomes for Diverse Adults | Non-RCT |
| Carter, Nd; Khan, Km; Petit, Ma; Heinonen, A; Waterman, C; Donaldson, Mg; Janssen, Pa; Mallinson, A; Riddell, L; Kruse, K; Prior, Jc; Flicker, L; McKay, Ha | Results of a 10 week community based strength and balance training programme to reduce fall risk factors: a randomised controlled trial in 65-75 year old women with osteoporosis | Wrong intervention |
| Sadler, M. S.; Wash, K.; DePaul Trumbach, L. M.; Cronan, T. A. | A secondary analysis of three types of social support in relation to self-efficacy, disease impact, and depression in fibromyalgia | Wrong intervention |
| Webster, Vs; Holdsworth, Lk; McFadyen,; Little, H | Self-referral, access and physiotherapy: patients' knowledge and attitudes. Results of a national trial | Wrong intervention |
| Ding, E. L.; Feigl, A. B.; Watson, K. T.; Ng, T. L. J.; Makerechi, L.; Bui, N.; Ireifij, A.; Farraj, R.; Zoughbie, D. E. | Social network enhanced behavioral interventions for diabetes and obesity: A 3 arm randomized trial with 2 years follow-up in Jordan | Wrong intervention |
| Robinson, W. L.; Whipple, C. R.; Jason, L. A.; Cafaro, C.; Lemke, S.; Keenan, K. | Social Support Coping for African American Adolescents: Effect of a Culturally Grounded Randomized Controlled Trial Intervention | Wrong patient population |
| Warner, L. M.; Jiang, D.; Yeung, D. Y.; Choi, N. G.; Ho, R. T. H.; Kwok, J. Y. Y.; Song, Y.; Chou, K. L. | Study protocol of the 'HEAL-HOA' dual randomized controlled trial: Testing the effects of volunteering on loneliness, social, and mental health in older adults | protocol |
| Elsworth, C; Winward, C; Sackley, C; Meek, C; Freebody, J; Esser, P; Hooshang, I; Soundy, A; Barker, K; Hilton-Jones, D | Supported community exercise in people with long-term neurological conditions: A phase II randomized controlled trial | Wrong patient population |
| ￼ | Supporting community-based exercise in long-term neurological conditions: experience from the Long-term Individual Fitness Enablement (LIFE) project | Wrong intervention |
| Geraci, J. C.; Dichiara, A.; Greene, A.; Gromatsky, M.; Finley, E. P.; Kilby, D.; Frankfurt, S.; Edwards, E. R.; Solomon Kurz, A.; Sokol, Y.; Sullivan, S. R.; Mobbs, M.; Seim, R. W.; Goodman, M. | Supporting servicemembers and veterans during their transition to civilian life using certified sponsors: A three-arm randomized controlled trial | Wrong patient population |
| Kolster, A.; Heikkinen, M.; Pajunen, A.; Mickos, A.; Wennman, H.; Partonen, T. | Targeted health promotion with guided nature walks or group exercise: a controlled trial in primary care | Non-RCT |
| Bisby, M. A.; Barrett, V.; Staples, L. G.; Nielssen, O.; Dear, B. F.; Titov, N. | Things You Do: A randomized controlled trial of an unguided ultra-brief intervention to reduce symptoms of depression and anxiety | Wrong intervention |
| Lutenbacher, M.; Elkins, T.; Dietrich, M. S. | Using Community Health Workers to Improve Health Outcomes in a Sample of Hispanic Women and Their Infants: Findings from a Randomized Controlled Trial | Wrong patient population |
| Sweeney, A. M.; Wilson, D. K.; Zarrett, N.; Simmons, T.; Mansfield, M.; Decker, L. | Using formative process evaluation to improve program implementation and accessibility of competitive group-based physical activity in the TEAM-PA trial | Wrong intervention |
| Meins, I. A.; Muijsson-Bouwman, D. C.; Nijman, S. A.; Greaves-Lord, K.; Veling, W.; Pijnenborg, G. H. M.; van der Stouwe, E. C. D. | VR-SOAP, a modular virtual reality treatment for improving social activities and participation of young people with psychosis: a study protocol for a single-blind multi-centre randomized controlled trial | Protocol |
| Greenwood, S.; Young, H.; Briggs, J.; Walklin, C.; Mangahis, E.; Billany, R.; Cooper, N.; Worboys, H.; Castle, E.; Campbell, J.; Bramham, K.; Macdonald, J. | WCN24-1556 The clinical and health economic effect of a 6-month physical activity digital health intervention on health-related quality of life in people with chronic kidney disease | Conference abstract/presentation |
| Hartmann, Steffen; Timm, Christina; Barnow, Sven; Rubel, Julian A.; Lalk, Christopher; Pruessner, Luise | Web-Based Cognitive Behavioral Treatment for Bulimia Nervosa: A Randomized Clinical Trial | Wrong intervention |
| Wu, C. Y.; Yu, K.; Arnold, S. E.; Das, S.; Dodge, H. H. | Who Benefited Most from the Internet-Based Conversational Engagement RCT (I-CONECT)? Application of the Personalized Medicine Approach to a Behavioral Intervention Study | Wrong intervention |
| Mercer SW, Fitzpatrick B, Grant L, et al. | Effectiveness of community-links practitioners in areas of high socioeconomic deprivation. | Non-RCT |
| McDermott RA, Schmidt B, Preece C, et al | Community health workers improve diabetes care in remote Australian indigenous communities: Results of a pragmatic cluster randomized controlled trial. | Non-RCT |
| Gallegos-Carrillo K, García-Peña C, Salmerón J, et al. | Brief Counseling and Exercise Referral Scheme: A Pragmatic Trial in Mexico | Non-RCT |
| Gallegos-Carrillo K, Garcia-Peña C, Salgado-de-Snyder N, et al. | Levels of adherence of an exercise referral scheme in primary health care: effects on clinical and anthropometric variables and depressive symptoms of hypertensive patients | Non-RCT |
| Thomson, L.J., Lockyer, B., Camic, P.M. and Chatterjee, H.J., | Effects of a museum-based social prescription intervention on quantitative measures of psychological wellbeing in older adults. Perspectives in public health, 138(1), pp.28-38. | Non-RCT |
| Elston J, Gradinger F, Asthana S, et al. | Does a social prescribing 'holistic' link-worker for older people with complex, multimorbidity improve well-being and frailty and reduce health and social care use and costs? A 12-month before-and-after evaluation. | Non-RCT |
| Giebel C, Morley N, Komuravelli A | A socially prescribed community service for people living with dementia and family carers and its long‐term effects on well‐being. | Non-RCT |
| Jones C, Hartfiel N, Brocklehurst P, et al. | Social return on investment analysis of the health Precinct community hub for chronic conditions. | Non-RCT |
| Kim JE, Lee YL, Chung MA, et al. | Effects of social prescribing pilot project for the elderly in rural area of South Korea during COVID-19 pandemic | Non-RCT |
| Loftus AM, McCauley F, McCarron MO. | Impact of social prescribing on general practice workload and polypharmacy. | Non-RCT |
| Kellezi B, Wakefield JRH, Stevenson C, et al. | The social cure of social prescribing: a mixed-methods study on the benefits of social connectedness on quality and effectiveness of care provision | Non-RCT |
| Coop British Red Cross. | Tackling-loneliness-and-isolation-connecting-communities. | Non-RCT |
| Waterfall D. Dudley CVS: | integrated plus impact evaluation | Non-RCT |
| Kimberlee R, Ward R, Jones M, et al. | Measuring the economic impact of wellspring healthy living centre’s social prescribing wellbeing programme for low level mental health issues encountered by GP services | Non-RCT |
| Dayson C, Bennett E. | Evaluation of Doncaster Social Prescribing Service: understanding outcomes and impact, 2016. | Non-RCT |
| Dayson C. | Evaluating social innovations and their contribution to social value: the benefits of a ‘blended value’ approach, 2017 | Non-RCT |
| Alden S, Wigfield A, Kispeter E. | CIRCLE (Centre for international research on care, labour and equalities) University of Leeds: 60. | Non-RCT |
| Callaghan, L.; Thompson, T.P.; Creanor, S.; Quinn, C.; Senior, J.; Green, C.; Hawton, A.; Byng, R.; Wallace, G.; Sinclair, J.; et al. | Individual health trainers to support health and well-being for people under community supervision in the criminal justice system: The STRENGTHEN pilot RCT. | Wrong intervention |
| Afuwape, Sarah A., Tom KJ Craig, Tirril Harris, Marva Clarke, Anneline Flood, Dele Olajide, Eleanor Cole, Morven Leese, Paul McCrone, and Graham Thornicroft. | The Cares of Life Project (CoLP): an exploratory randomised controlled trial of a community-based intervention for black people with common mental disorder | Wrong patient population |
| Carnes D, Sohanpal R, Frostick C, et al. | The impact of a social prescribing service on patients in primary care: a mixed methods evaluation | Non-RCT |

Supplementary Table 12. Study characteristics

| **Study ID** | **City, Country** | **Study duration** | **Sample size (randomised)** | **Reasons for lost to follow-up** | **Population age (**mean± standard deviation or range**)** | **%Female; %Male** | **Disease condition** | **Educational background** | **Ethnicity** | **Funding Source** |
| --- | --- | --- | --- | --- | --- | --- | --- | --- | --- | --- |
| **Spencer 2018** | Detroit, USA | 18 months | 222 | Withdrew, deceased, missed data collection | Mean 48.9 ± 10.6 years | Female: 61%; Male: 39% | T2DM | High school graduates: 30.6% | Latino (100%) | Peers for Progress grant from the American Association of Family Physicians Foundation, by the National Institute of Diabetes and Digestive and Kidney Diseases (grant P30-DK092926 to the Michigan Center for Diabetes Translational Research and grant R18-DK-0785501A1), and by the Centers for Disease Control and Prevention (cooperative agreement no. U50/CCU417409). |
| **Kangovi 2018** | Philadelphia, Pennsylvania, USA | 14 months | 592 | Lost to the CHW despite 10 outreach attempts (including 1 home visit) or said they no longer wanted to work with a CHW | Goal Setting Alone 52.1 ± 11.5; Goal Setting Plus CHW 53.1 ± 10.7 years | Female: 62%; Male: 38% | Diagnosis for 2 or more targeted chronic diseases | NR | Goal Setting Alone: African American 274 (95.1%), Hispanic 0 (0%); Goal Setting Plus CHW: African American 284 (93.4%), Hispanic 11 (3.7%) | National Institutes of Health National Heart, Lung, and Blood Institute  Patient-Centered Outcomes Research Institute Award. |
| **Kangovi 2017** | Philadelphia, Pennsylvania, USA | 15 months | 302 | Either lost to the CHW despite 10 attempts to contact the patient (including 1 home visit) or said they no longer wanted to work with a CHW | Goal Setting Alone 56.1 ± 12.6; Goal Setting Plus CHW 56.6 ± 13.6. years | Female: 75%; Male: 25% | Diagnosed with 2 or more chronic diseases (diabetes, obesity, tobacco dependence, hypertension) | Education not specified. Health literacy: Goal Setting Alone: 2.2 ± 3; Goal Setting Plus CHW 2.1 ± 1.3 | Goal Setting Alone: African American 144 (94.7%), Hispanic 4 (2.7%); Goal Setting Plus CHW: African American 142 (94.7%), Hispanic 4 (2.8%) | University of Pennsylvania, School of Medicine Agency for Healthcare Research and Quality Institutional Career Development grant, the National Heart, Lung, and Blood Institute |
| **Spencer 2013** | Detroit, USA | 22 months | 183 | Conflict in schedule, deceased, medical condition, moved out of area, not interested, unable to contact, changed healthcare provider | Intervention: 50 (47 to 52 95% CI); Control: 55 (53 to 57 95% CI) years | Intervention: 75% Female; Control: 67% | T2DM | Highschool graduate: Intervention 60%; Control 59% | African American (Intervention: 53%; Control: 61%);  Latino (Intervention: 47%; Control: 39%) | National Institute of Diabetes and Digestive and Kidney Disease, Centers for Disease Control and Prevention, the Michigan Diabetes Research and Training Center and the Robert Wood Johnson Foundation Clinical Scholars Program. |
| **Gray 2021** | Public Health-Seattle King County, USA | 27 months | 287 | Unwilling to continue, lost to follow-up, withdrawn, moved out of area | Intervention: 51.8 ± 9.4: Control: 53.5 ± 8.9 years | Female: 50%; Male: 50% | T2DM | Less than high school education: 33.1 vs 36.2 | Black: 28.6 vs 23.8; Latinx: 40.6 vs 42.3; White: 21.1 vs 21.5 | National Institutes of Health, National Institute of Diabetes and Digestive and Kidney Diseases and United States Department of Veterans Affairs, Health Services Research & Development Program |
| **Ramirez 2019** | Chicago, Illinois, and San Antonio, Texas, USA | 15 months | 288 | Lost to follow-up | 56.05 ± 10.20 years | Female: 44%; Male: 46% | Breast, Prostate, and Colorectal Cancer Survivors | Less than high school: 42%; High school: 26; Junior college or more: 19% | Latino (100%) | National Cancer Institute; the Mays Cancer Center at University of Texas Health San Antonio; the Robert H. Lurie Comprehensive Cancer Center at Northwestern University |
| **Carrasquillo 2017** | Miami, USA | 4.5 years | 300 | Lost to follow-up | 55.2 ± 7.0 years | Female: 55%; Male: 45% | T2DM | Educational attainment < year 12: 40% | Latino (100%) | University of Miami |
| **Bossche 2021** | Ghent, Belgium | 4 months | 135 | Refusing to fill in the second questionnaire | 60.04 years (range 19-93) | Female: 62.2%; Male: 37.8% | Psychiatric history, or a precarious Social context, or an uncertain residence status, or a chronic illness | No or primary school: 18%; Primary secondary education: 26%; Higher secondary education: 30%; Higher education: 26% | European: 75%; African: 7%; Eastern Mediterranean: 9% and others | None |
| **Patel 2023** | Atlantic City, NJ, and Chicago, IL, USA | 33 months | 160 | Died | Total: Median age: 58 (21-89) Intervention: Median age 58 (21-80); Control: 58 (31-89) | Female: 51%; Male: 49% | Cancer (newly diagnosed with all solid tumour and hematologic malignancies or recurrent disease) | Less than high school: 80.6%; High school: 15.6%; 2 year college or Bachelor degree: 3.8% | White: 51.3%; African American or Black: 27.5%; Asian 19.3%; South-East Asian 7.5%; Vietnamese: 6.2%; Chinese 5.6%; American Indian or Alaska Native: 1.3%; Native Hawaiian 0.6% | National Institute on Minority Health and Health Disparities of the National Institutes of health |
| **McQueen 2024** | Louisiana, USA | 44 months | 473 | Died, lost Louisiana Healthcare Connections (LHCC) membership, did not respond to multiple outreach attempts, another 3 either did not engage until months into the intervention period | Intervention: 51.79 ± 9.35; Control 51.31 ± 9.71 years | Female: 75%; Male: 25% | Type II diabetes | Less than high school: Intervention: 77 (32.9%); Control: 74 (31.0%) High School/GED: Intervention: 74 (31.6%); Control: 77 (32.2%); Some college/degree: Intervention: 81 (34.6%); Control: 88 (36.8%) | Non-Hispanic Black/African American: Intervention: 150 (64.1%); Control: 152 (63.6%); Non-Hispanic White: Intervention: 65 (27.8%); Control: 66 (27.6%) | National Institute for Diabetes and Digestive and Kidney Diseases |
| **Yamashita 2024** | Kumamoto, Japan | 3 months | 59 | Open heart surgery, personal reasons, family care, ankle sprain. | 72.5 ± 5.3 years | Female: 30%; Male: 70% | CVD | NR | NR | Japan Sports Agency |
| **Watts 2024** | Kansas City, USA | 4 years | 219 | Health problems, change in circumstances, COVID-19 concerns, serious adverse event, problem with testing, problem with waitlist group | 71.7 ± 4.7 years | Female: 81.3%; Male: 18.7% | Sedentary/ underactive older adults | Graduate degree 36.1%; College degree 42.0%; Some college credits, no degree 7.8%; High school degree 12.8%; No high school degree 1.4% | White 86.8%; African American 11.9%; Asian 0.9%; Other 0.5% | National Institute on Aging |
| **Joo 2024** | North East, USA | 26 months | 149 | Medical illness, difficulty adhering to research assessments, poor hearing, and limited time | 69.5 (51- 95) years | Female 84%; Male: 16% | Depression | Grade school: 1.5%; Some high school, high school or obtained GED: 38.1%; 1-3 years of college or technical school: 30.6%; At least 4 years of college or postgraduate: 29.9% | American Indian or Alaska Native 0.7%; Asian 0.7%; Black 52.2%; White 41.0%; Multiracial 4.5%; Other 0.7% | National Institute of Mental Health and the National Institute on Aging Mentored Patient-Oriented Research Career Development Award |
| **Nyamathi 2024** | Los Angeles, California, USA | 24 months | 36 | Homeless adults | 53.7 ± 11.8 years | Female 20%; Male: 80% | Homelessness people | NR | Latin: 36%; Black: 36% | University of California |
| **Mbuthia 2024** | Kiambu County, Kenya | 6 months | 80 | NR | Intervention: 47.3 ± 14.4; Comparator: 46.3 ±9.7 years | Female 88%; Male: 12% | Hypertension | NR | Africans | Consortium for Advanced Research Training in Africa |
| **Isaacs 2007** | Barnet, London, UK | 3 years | 949 | Pre-existing vascular disease which was not disclosed prior to randomisation or, in one case, because of withdrawal of consent | 57 ± 9 years | Female 33%; Male: 77% | Not currently physically active and with at least one cardiovascular risk factor | No formal education: Fitness for life: 23%; Walking partners: 22%; Control: 20% | Asian: 14%; White: 76% | Department of Health, UK |
| **Laroche 2024** | Iowa, USA | 12 months | 208 | Discontinued intervention, no longer interested, language barrier | 35.1 ± 0.7 years | Female 95%; Male:5% | Obesity and diabetes | No high school degree: 27%; High school graduate: 50%; College graduate: 23% | Black/African American: 34%; Hispanic: 27% | National Heart, Lung, and Blood Institute of the National Institutes of Health |
| **Wagner 2023** | Connecticut, USA | 4.5 years | 188 | Discontinued, removed for medical reasons | 54 years | Female: 78%; Male: 22% | People with elevated risk for diabetes | NR | Cambodian or Cambodian-American | Penn State from Proactive Life LLC |
| **Ugarte 2023** | California, USA | 6 weeks | 300 | NR | 38-39 Years | Female 80%; Male: 20% | NR | Some College/Certificate:30%; Bachelor’s degree:30%; | Non-Hispanic White: 74%; Non-Hispanic Black: 9%; Hispanic: 11%; Asian:4% | National Institute of Allergy and Infectious Diseases and the National Institute of Mental Health |
| **Islam 2023** | New York City, USA | July 2017 to September 2018 | 303 | Lost to follow-up | 56.8 ± 11.2 years | Female 54%; Male: 56% | SBP≥140 or DBP≥90 mm Hg within the previous 6 months | Less than high school: 45%; High school/some college: 22%; College graduate: 32 | South Asian ethnicity | Centers for Disease Control and Prevention, National  Institutes of Health; National Institute of Diabetes and Digestive and Kidney Diseases, National Institute on Minority Health and Health |
| **Heisler 2022** | Detroit, USA | February 2018–June 2020 | 3159 | Lost to follow-up | 29 years | Female 64%: Male: 36% | NR | NR | NR | Blue Cross, Blue Shield of Michigan Foundation, the Michigan Department of Health and Human Services, the Ralph C. Wilson Foundation, Poverty Solutions at the University of Michigan, and the National Institute of Diabetes and Digestive and Kidney Diseases |
| **Rovner 2023** | Philadelphia, PA, USA | NR | 200 | Loss of interest and died | 64.9 ± 9.5 years | Female 73%; Male: 27% | Type 1 or type 2 diabetes | 12+ | Black | Pennsylvania Department of Health |
| **Gusi 2008** | Cáceres, Spain | 2003-2004 | 127 | Had to care for a relative | Intervention: 71 ± 5; Control: 74 ± 6 | Female 100% | Moderately depressed and over weight | Primary school or higher: ~40% | Spanish | European Social Funds and the Government of Extremadura, Spain |
| **Harrison 2005** | Local authority borough in the north-west of England | March 2000 and December 2001 | 545 | Did not return postal questionnaire | Intervention 40% 18-44 years, 37% 45-59 years, 23% ≥60 years; Control 40% 18-44 years, 36% 45-59 years, 24% ≥60 years | Female: 77%; Male: 33% | NR | NR | White | Health Authority/Primary Care Trust and the Metropolitan Borough Council |
| **Ma 2018** | Guangzhou, China | March 2015 to November 2016 | 158 | Time limitation, no interest, hospital admission, moved other places, default, lost contact and died | Intervention: 70.24 ± 10.5; Control: 69.71 ± 10.8 years | Female: 32%; Male 68% | Hypertension diagnosed 5-10 years ago | Secondary school and below: 50%; High school and above:50% | Chinese | Guangdong Provincial Department of Science and Technology, the Guandong Special Program for Scientific Development |
| **Murphy 2012** | Wales, UK | 2008-2008 | 2160 | NR | 52 ± 14.7 years | Female: 66%; Male: 44% | Coronary heart disease risk (CHD, 1559, 72%), mild to moderate depression, anxiety or stress (79, 4%) or both (522, 24%) | Beyond min school leaving age: 52% | White | Welsh Assembly Government |
| **Smith 2019** | Exeter and Birmingham, UK | November 2014 to June 2015 | 314 | Health problems, family issues, disliked measures, bereavement | Intervention: 61 ± 10; Control: 61 ± 9.9 years | Female: 44%; Male: 66% | High risk for type 2 diabetes due to elevated blood glucose (fasting plasma glucose 6.1-6.9 mmol/l, or HbA1c 42- 47 mmol/mol) and BMI ≥25 kg/m2 | Primary/some secondary: Intervention 2%, Control 6% | White British: 74%; White other: 4%; Asian:15%; Black:7%; Mixed:1% | National Institute for Health Research (NIHR) |
| **Taylor 1998** | Hailsham, East Sussex, UK | 1996-1997 | 142 | NR | Intervention: 54 ± 8; Control: 54 ± 9 years | Female 62%; Male: 38% | Hypertensive (SBP ≥140 mmHg, DBP ≥90 mmHg) or overweight (BMI >25 kg/m2) | NR | NR | The South Thames Regional Health Authority Primary Care Development Fund |
| **Clarke 1992** | Melton Mowbray, Leicestershire, UK | 1985-1988 | 523 | High dependency, terminally ill and died | Over 75 years | NR | NR | NR | NR | Nuffield Provincial Hospitals Trust and Leicestershire Health Authority |
| **Grant 2000** | Avon, Bristol, UK | August 1997 and September 1998 | 161 | NR | Mean 43.2 years | Female 75%; Male: 25% | NR | NR | NR | Avon Health Authority |
| **Babamoto 2009** | Los Angeles, USA | July 2002 and July 2003 | 318 | Moved out of the area, disenrolled from the study, or could not be contacted during the follow-ups | 50 (18-87) years | Female: 64%; Male: 36% | T2DM, diagnosed 6 months ago | Less than a sixth-grade education: 61% | Hispanic/Latino | Pfizer Foundation and Pfizer Health Solutions Inc |
| **Lamb 2002** | Reading, UK | NR | 260 | Lost to follow up | 50.2 (8.1) years | Female: 51%; Male: 49% | Healthy adults | NR | NR | British Heart Foundation and Countryside Agency |
| **Kiely 2024** | Dublin, Ireland | July 2020 to January 2021 | 251 | Lost to follow-up, uncontactable, declined, hospitalised, deceased |  | Female: 63%; Male: 37% | Multimorbid conditions | Primary education or below 30% in intervention and 25% in control group | White | Health Research Board Ireland and Department of Health Sláintecare Integration Fund |

T2DM: Type 2 diabetes mellitus; NA: Not available; NR: Not reported; CHD: Coronary heart disease; CVD: cardiovascular disease; CHW: Community health worker; SD: standard deviation; BMI: Body mass index; CHW: Community healthcare worker; USA: United States of America; UK: United Kingdom

Supplementary Table 13. Intervention characteristics

| **Reference** | **Referral methods** | **Intervention details** | **Control details** |
| --- | --- | --- | --- |
| Spencer, M. S., Kieffer, E. C., Sinco, B., Piatt, G., Palmisano, G., Hawkins, J., ... & Heisler, M. (2018). Outcomes at 18 months from a community health worker and peer leader diabetes self-management program for Latino adults. Diabetes care, 41(7), 1414-1422. | **Referral pathway:** Indirect  **Referral made by:** CHWs  **Link worker background:**  The CLWs were Spanish-fluent Latinas from the southwest Detroit community who had completed high school or obtained a GED. They received comprehensive training, including 160 hours on community link work, 80 hours on diabetes education, and instruction in empowerment-based approaches, motivational interviewing, and goal setting | **Setting**: Community, home and clinic  **Sample size:** 60  **Intervention Name:** “Journey to Health/ El Camino a la Salud”  **Intervention Description**: A culturally tailored DSME curriculum, grounded in the empowerment approach that emphasizes a collaborative approach to facilitate self-directed behaviour change of patients  **Intervention Deliverer(s)**: Community Health Workers & Peer Leaders  **Intervention Duration:** 18 months (6-month program + 12-month program)  **Frequency of Occurrence:** Fortnightly and monthly (6-month program). Weekly (CHW+PL 12-month program)  **Time per Occurrence**: 6-month program: two 60-min home visits each month and 11 x 2 hr group sessions per fortnight.  **Most Effective Components:** Including a peer leader was most effective (i.e., PL + CHW). The CHW+PL group had a significant intervention effect on reducing depressive symptoms at 18 months.  **Least Effective Components**: CHW only (with no peer leader and received only telephone calls to check the progress and meeting goals) | **Type of control:** Active  **Sample size:** 73  **Description of control group:**  The Enhanced Usual Care (EUC) group received a 2-h class conducted by a research assistant covering how to interpret their clinical and anthropometric results. EUC participants were contacted once each month to update contact information |
| Kangovi, S., Mitra, N., Norton, L., Harte, R., Zhao, X., Carter, T., ... & Long, J. A. (2018). Effect of community health worker support on clinical outcomes of low-income patients across primary care facilities: a randomized clinical trial. *JAMA internal medicine*, *178*(12), 1635-1643. | **Referral pathway:** Indirect  **Referral made by:** CHWs  **Link worker background:**  The CHWs are required to have a high school diploma and are screened through behavioural interviews for personality traits, such as empathy. They undergo a month-long training that covers topics such as action planning and motivational interviewing. They are supervised by a manager (typically with a master’s degree in social work) who provides real-time support, ongoing training, and help with clinical integration | **Setting:** Community and primary care  **Sample size:** 304  **Intervention Name:** IMPaCT intervention  **Intervention Description:** Participants set a chronic disease management goal with their primary care physician; those randomized to the CHW intervention received 6 months of tailored support. IMPaCT occurs in 3 stages: goal-setting, tailored support, and connection with long-term support.  **Intervention Deliverer(s)**: CHWs  **Intervention Duration**: 6 months  **Frequency of Occurrence:** The CHWs communicated with patients at least once per week, including monthly face-to-face contact. If a patient was hospitalized during the intervention, CHWs visited the patient and coordinated with the inpatient care team  **Time per Occurrence:** Not specified  **Most Effective Components:** Attributed to the role of the CHW as a "trusted mediator between disadvantaged patients and formal health care organizations". CHWs share a background with disadvantaged patients. "This shared identity allows CHWs to establish trust, offer practical support based on their own experience of common problems, and provide non-judgmental support."  **Least Effective Components**: Not clear | **Type of control:** Passive  **Sample size:** 288  **Description of control group:** Patients assigned to goal-setting only (control arm) went on to receive usual care |
| Kangovi, S., Mitra, N., Grande, D., Huo, H., Smith, R. A., & Long, J. A. (2017). Community health worker support for disadvantaged patients with multiple chronic diseases: a randomized clinical trial. *American journal of public health*, *107*(10), 1660-1667. | **Referral pathway:** Indirect  **Referral made by:** CHWs  **Link worker background:**  Community health workers, with high school diploma. 1 month training in motivational interviewing, action planning and on the job. Based in primary care clinics | **Setting:** Community and primary care  **Sample size:** 150  **Intervention Name:** IMPaCT intervention  **Intervention Description:** CHWs provide tailored coaching, social support, advocacy, and navigation to outpatients with multiple chronic conditions. The intervention was developed and refined through a participatory action research framework. The intervention consists of 3 stages: action planning, tailored support, and connection with long-term support  **Intervention Deliverer(s):** CHW  **Intervention Duration:** 6 months  **Frequency of Occurrence:** Monthly face-to-face meetings and weekly telephone check-ins  **Time per Occurrence:** Not specified  **Most Effective Components:** Attributed to the flexibility of the CHW's approach and tailored, personalized goal setting and action plans focused on health behaviour change and psychosocial needs  **Least Effective Components:** Not clear | **Type of control:** Active  **Sample size:** 152  **Description of control group:**  Goal Setting Alone (no CHW). Patients assigned to goal-setting alone went on to receive usual care in accordance with guidelines at each site (including potential referrals to a social worker or diabetes nutritionist) |
| Spencer, M. S., Hawkins, J., Espitia, N. R., Sinco, B., Jennings, T., Lewis, C., ... & Kieffer, E. (2013). Influence of a community health worker intervention on mental health outcomes among low-income Latino and African American adults with type 2 diabetes. *Race and Social Problems*, *5*, 137-146.  Spencer, M. S., Rosland, A. M., Kieffer, E. C., Sinco, B. R., Valerio, M., Palmisano, G., ... & Heisler, M. (2011). Effectiveness of a community health worker intervention among African American and Latino adults with type 2 diabetes: a randomized controlled trial. *American journal of public health*, *101*(12), 2253-2260. | **Referral pathway:** Indirect  **Referral made by:** CHW/family health advocates  **Link worker background:**  Trained CHWs | **Setting:** Community, home, clinic  **Sample size:** 84  **Intervention Name:** A diabetes intervention conducted since 2000 by the REACH Detroit Partnership, as part of the Centres for Disease Control and Prevention (CDC)-funded Racial and Ethnic Approaches to Community Health (REACH) Initiative  **Intervention Description:** Using an empowerment-based approach, community health workers provided participants with diabetes self-management education and regular home visits and accompanied them to a clinic visit during the 6-month intervention period  **Intervention Deliverer(s)** : CHW/ 'Family Health Advocates'  **Intervention Duration**: 6 months  **Frequency of Occurrence:** Fortnightly (diabetes education classes), monthly (home visits) and 1 clinic visit (during 6 month intervention)  **Time per Occurrence**: 2 hours (group classes); approx. 60 minutes (home visits); clinic visit (not specified)  **Most Effective Components**: Unclear but CHWs who were trained in empowerment approaches and promoted diabetes self-management in their own communities formed the most active part of the intervention  **Least Effective Components:** Not clear | **Type of control:** Passive  **Sample size:** 99  **Description of control group:**  Participants in the control group were contacted once per month to update contact information |
| Gray, K. E., Hoerster, K. D., Taylor, L., Krieger, J., & Nelson, K. M. (2021). Improvements in physical activity and some dietary behaviors in a community health worker-led diabetes self-management intervention for adults with low incomes: results from a randomized controlled trial. *Translational Behavioral Medicine*, *11*(12), 2144-2154.  Nelson K, Taylor L, Silverman J, et al. Randomized Controlled Trial of a Community Health Worker Self-Management Support Intervention Among Low-Income Adults With Diabetes, Seattle, Washington, 2010-2014. *Prev Chronic Dis* 2017; **14**: E15. | **Referral pathway:** Indirect  **Referral made by:** CHWs  **Link worker background:**  High school or equivalent degrees and 5–8 years of experience as CLWs | **Setting:** Home-based, community resource locations  **Sample size:** 130  **Intervention Name:** PeerAID intervention  **Intervention Description:** The PeerAID intervention was delivered by two bilingual (Spanish/English) Community Health Workers (CHWs) employed by PHSKC. The intervention involved an initial intake and five follow-up home visits per participant, guided by protocols specifying education content, client skill development goals, and CHW actions. Participants received low-literacy diabetes education materials in English and Spanish  **Intervention Deliverer(s):** CHW  **Intervention Duration**: 10 months  **Frequency of Occurrence**: Baseline, 0.5, 1.5, 3.5, 7 and 10 months  **Time per Occurrence**: NR  **Most Effective Components**: Social support  **Least Effective Components**: Group-based diabetes education classes | **Type of control:** Active  **Sample size:** 133  **Description of control group:**  Received usual care and were offered one optional diabetes self-management educational visit after completing the 12-month outcome assessment |
| Ramirez, A. G., Choi, B. Y., Munoz, E., Perez, A., Gallion, K. J., Moreno, P. I., & Penedo, F. J. (2020). Assessing the effect of patient navigator assistance for psychosocial support services on health‐related quality of life in a randomized clinical trial in Latino breast, prostate, and colorectal cancer survivors. *Cancer*, *126*(5), 1112-1123. | **Referral pathway: Ind**irect  **Referral made by:** Patient navigator, lay community health workers or promotors  **Link worker background:**  NR | **Setting:** Community  **Sample size:** 144  **Intervention Name:** The Patient Navigator LIVESTRONG Cancer Navigation Services program  **Intervention Description:** For the first three months, PN-LCNS participants received patient navigator (PN) services to facilitate their engagement with the phone-based PN-LCNS program (available throughout the 12-month study). Navigators introduced the program, encouraged its use, addressed barriers, provided community resource information (social work, psychosocial services, care, transportation, financial aid), and assisted with medical appointments  **Intervention Deliverer(s):** Lay community health workers or promotors  **Intervention Duration:** 12 months  **Frequency of Occurrence:** Weekly (first 3 months), then monthly at other follow-ups  **Time per Occurrence:** Not specified.  **Most Effective Components:** Coping with emotional concerns  **Least Effective Components:** Education about risks to fertility, preservation options, and access to discounted fertility services | **Type of control:** Active  **Sample size:** 144  **Description of control group:** Print materials relevant to breast, colorectal, and prostate cancer survivorship from organizations |
| Carrasquillo, O., Lebron, C., Alonzo, Y., Li, H., Chang, A., & Kenya, S. (2017). Effect of a community health worker intervention among Latinos with poorly controlled type 2 diabetes: the Miami healthy heart initiative randomized clinical trial. *JAMA internal medicine*, *177*(7), 948-954. | **Referral pathway:** Indirect  **Referral made by:** Patient  Clinicians  **Link worker background:**  NR | **Setting:** Community  **Sample size:** 150  **Intervention Name:** Community health worker (CHW) intervention  **Intervention Description:** 4 home visits, 12 calls during the 12 months & bimonthly exercise groups in parks Total interventions: home visits received- 5, Telephone calls- 20; Participants (126 (84%) received 12 CLW contacts per year Mode: Individualised, group face to face  **Intervention Deliverer(s):** CHWs  **Intervention Duration:** 12 months  **Frequency of Occurrence:** Weekly  **Time per Occurrence:** 75 hours  **Most Effective Components:** Bimonthly exercise groups in parks  **Least Effective Components:** One on one counselling and coaching, and assistance with resource referrals | **Type of control:** Passive  **Sample size:** 150  **Description of control group:**  Enhanced Usual Care |
| Vanden Bossche, D., Lagaert, S., Willems, S. and Decat, P., 2021. Community health workers as a strategy to tackle psychosocial suffering due to physical distancing: a randomized controlled trial. *International journal of environmental research and public health*, *18*(6), p.3097. | **Referral pathway:** Indirect  **Referral made by:** CHWs  **Link worker background:**  NR | **Setting:** Community  **Sample size:** 67  **Intervention Name:** Community health worker’s psychosocial support  **Intervention Description:** CHWs provided 8 weeks of hands-on, tailored support to patients spanning the domains of social support, coaching, advocacy, and navigation to healthcare if needed  **Intervention Deliverer(s):** CHWs  **Intervention Duration:** 8 weeks  **Frequency of Occurrence:** Weekly  **Time per Occurrence:** 2 hours  **Most Effective Components:** Social support, coaching, advocacy, and navigation  **Least Effective Components:** Offering a sympathetic ear and gave attention to their patients’ worries, stories, and questions | **Type of control:** Passive  **Sample size:** 68  **Description of control group:**  Usual care: care as usual is provided by professional primary care provider |
| Patel MI, Kapphahn K, Wood E, et al. Effect of a Community Health Worker-Led Intervention Among Low-Income and Minoritized Patients With Cancer: A Randomized Clinical Trial. *J Clin Oncol* 2024; 42(5): 518-28. | **Referral pathway:** Indirect  **Referral made by:** CHWs  **Link worker background:**  Bilingual in Spanish, Hindi, Vietnamese, and Chinese. From 2021 pilot: "Health Advocates undergo several weeks of training including an annual 2-day refresher course in motivational interviewing, community health, and complex disease management." | **Setting:** Community, telephone calls (home)  **Sample size:** 80  **Intervention Name:** Lay Health Workers Educate, Engage, and Activate Patients to Share (LEAPS - includes an added component to assist patients with identified HRSNs  **Intervention Description:** A multilevel, multicomponent community health worker (CHW)–led advance care planning, cancer symptom management, and health-related social needs intervention. The CHW assisted participants with advance care planning (ACP), proactively screened identified health-related social needs  **Intervention Deliverer(s):** CHWs  **Intervention Duration:** 12 months  **Frequency of Occurrence:** Weekly, by telephone, for 4 months and monthly thereafter up to 12 months or death, whichever was first  **Time per Occurrence:** 30 minutes  **Most Effective Components:** Unclear but potentially the capacity of bilingual CHW to communicate with patients in their preferred language. The frequency of weekly telephone calls (for 4 months) to screen for symptoms may also have potentially been most effective  **Least Effective Components:** Not clear | **Type of control:** Passive  **Sample size:** 80  **Description of control group:**  Usual care - outpatient care in oncology clinics |
| McQueen, A., von Nordheim, D., Caburnay, C., Li, L., Herrick, C., Grimes, L., ... & Kreuter, M. (2024). A Randomized Controlled Trial Testing the Effects of a Social Needs Navigation Intervention on Health Outcomes and Healthcare Utilization among Medicaid Members with Type 2 Diabetes. *International journal of environmental research and public health*, *21*(7), 936. | **Referral pathway:** Indirect  **Referral made by:**  Navigator (Social Needs Navigator)  **Link worker background:**  NR | **Setting:** Community  **Sample size:** 234  **Intervention Name:** Social Needs Navigation intervention  **Intervention Description:** The focus of the Social Needs Navigation intervention was to proactively help participants not otherwise identified for care management to address unmet social needs. If asked, navigators also offered encouragement, information, and resources related to health needs, but this was not their primary focus for outreach calls  **Intervention Deliverer(s):** Navigator (Social Needs Navigator)  **Intervention Duration:** 6 months  **Frequency of Occurrence:** The number and frequency of calls were determined by participants’ needs, interests, and willingness to interact. Either party could initiate a call  **Time per Occurrence:** No time limit  **Most Effective Components:** Addressing social needs: "Many calls with participants addressed food insecurity (e.g., participants were offered a referral to a food bank) or assistance paying utilities (e.g., participants were offered a referral for the Low-Income Home Energy Assistance Program [LIHEAP])  **Least Effective Components:** Not clear | **Type of control:** Passive  **Sample size:** 239  **Description of control group:** Usual care. Usual care participants could receive care management services if identified and referred by the health plan or providers to address high acuity needs |
| Yamashita, R., Sato, S., Sakai, Y., Tamari, K., Nozuhara, A., Kanazawa, T., ... & Tsujita, K. (2024). Effects of small community walking intervention on physical activity, well-being, and social capital among older patients with cardiovascular disease in the maintenance phase: A randomized controlled trial. *Journal of Physical Therapy Science*, *36*(3), 128-135. | **Referral pathway:** Unclear  **Referral made by:**  A registered cardiac rehabilitation instructor  **Link worker background:**  Healthcare worker | **Setting:** Community  **Sample size:** 29  **Intervention Name:** Small community walking group  **Intervention Description:** The SCW group participants walked in groups of three with their families or close friends as buddies to utilize the power of peer pressure  **Intervention Deliverer(s):** CR-registered instructor/ healthcare worker  **Intervention Duration:** 3 months  **Frequency of Occurrence:** Approx. once per week  **Time per Occurrence:** NR  **Most Effective Components:** Walking with a buddy  **Least Effective Components:** Not clear | **Type of control:** Active  **Sample size:** 30  **Description of control group:** Walking alone group |
| Watts, A., Szabo‐Reed, A., Baker, J., Morris, J. K., Vacek, J., Clutton, J., ... & Burns, J. M. (2024). LEAP! Rx: A randomized trial of a pragmatic approach to lifestyle medicine. *Alzheimer's & Dementia*, *20*(12), 8374-8386. | **Referral pathway:**  Mostly direct (53.9% physician referral; 46.1% self referral)  **Referral made by:**  Physician  **Link worker background:**  Physician/ GP | **Setting:** Community/YMCAs  **Sample size:** 219  **Intervention Name:** The LEAP Rx Program  **Intervention Description:** The LEAP Rx Program involved a 12-week intensive supervised exercise phase at YMCAs, followed by a 40-week lifestyle phase with intermittent supervision from a dedicated coach (52 weeks total). The exercise goal was 150 minutes/week of aerobic activity (3-5 days) and 2 days/week of resistance training. Participants also received monthly Alzheimer's prevention classes and a 1-year YMCA membership with access to group classes  **Intervention Deliverer(s):** LEAP! Rx coach  **Intervention Duration:** 12 months  **Frequency of Occurrence:** 150 min/week of aerobic exercise (over 3 to 5 days/week) and 2 days/week of resistance exercise  **Time per Occurrence:** Coaches provided generalized guidance to participants to increase total aerobic exercise duration by approximately 15 min each week over the initial 6 weeks to a goal of 150 min/week  **Most Effective Components:** Cardiorespiratory fitness  **Least Effective Components:** Not clear | **Type of control:** Active  **Sample size:** 109  **Description of control group:** Standard care |
| Hui Joo, J., Xie, A., Choi, N., Gallo, J. J., Zhong, Y., Ma, M., ... & Solomon, P. (2025). A mixed methods effectiveness study of a peer support intervention for older adults during the COVID-19 pandemic: results of a randomized clinical trial. *The American Journal of Geriatric Psychiatry*, *33*(4), 389-401. | **Referral pathway:**  Indirect  **Referral made by:**  Peer mentor  **Link worker background:**  Social workers | **Setting:** Telephone (due to COVID restrictions)  **Sample size:** 75  **Intervention Name:** Peer Enhanced Depression Care (PEERS)  **Intervention Description:** The PEERS intervention tested the effect of one-to-one peer support, focused on psychosocial support, self-care, and coping, against social interaction with nonpeers for improving access to and engagement in depression care among at-risk older adults  **Intervention Deliverer(s):** Peer mentor  **Intervention Duration:** 8 weeks  **Frequency of Occurrence:** Weekly  **Time per Occurrence:** Not specified for intervention group. For control group, the phone calls are 45 min duration.  **Most Effective Components:** The discussion of coping skills and self-care behaviour with a peer mentor  **Least Effective Components:** The delivery of self-care skills by peer mentors may not have been structured or intensive enough, suggesting the need for modifications in training and intervention content | **Type of control:** Active  **Sample size:** 74  **Description of control group:**  Social interaction control group |
| Nyamathi, A. M., Salem, B. E., Gelberg, L., Garfin, D. R., Wolitsky‐Taylor, K., Shin, S. S., ... & Lee, D. (2024). Pilot randomized controlled trial of biofeedback on reducing psychological and physiological stress among persons experiencing homelessness. *Stress and Health*, *40*(4), e3366. | **Referral pathway:** Indirect  **Referral made by:** CHWs  **Link worker background:**  Lay health leaders | **Setting:** Community-based clinic settings  **Sample size:** 17  **Intervention Name:** HRV-BF intervention  **Intervention Description:** Over 8 weeks, our RN/CHW team delivered weekly 30-minute individual in-person HRV-BF sessions using videos and scripted material. Each session included a 10-minute video teaching diaphragmatic breathing, breath-slowing techniques, and slow-paced breathing. Videos also provided basic education on stress and its effects, and guidance on daily application of HRV-BF techniques  **Intervention Deliverer(s):** CHWs  **Intervention Duration:** 8 weeks  **Frequency of Occurrence:** Weekly  **Time per Occurrence:** 30 minutes  **Most Effective Components:** Video content also included simple educational content on stress and the effects of stress on the body, as well as content designed to guide participants regarding day‐ to‐day applications of the HRV‐BF techniques  **Least Effective Components:** Not clear | **Type of control:** Active  **Sample size:** 19  **Description of control group:**  Health promotion (HP) intervention: The HP active control group was delivered over 30 min, once weekly for 8 weeks, by our RN/CHW team, trained to deliver scripted materialised content created |
| Mbuthia, G. W., Mwangi, J., Magutah, K., Oguta, J. O., Ngure, K., & McGarvey, S. T. (2024). Preliminary efficacy of a community health worker homebased intervention for the control and management of hypertension in Kiambu County, Kenya-a randomized control trial. *Plos one*, *19*(8), e0293791. | **Referral pathway:** Indirect  **Referral made by:** CHWs  **Link worker background:**  CHWs | **Setting:** Home  **Sample size:** 40  **Intervention Name:** CHW-led homebased intervention  **Intervention Description:** Home based community health worker–led intervention (health coaching, home BP monitoring, BP audit and feedback) implemented over a period of 6 months  **Intervention Deliverer(s):** CHWs  **Intervention Duration:** 6 months  **Frequency of Occurrence:** Two in the first month then monthly up to 6 months  **Time per Occurrence:** 90 minutes  **Most Effective Components:** Not clear  **Least Effective Components:** Not clear | **Type of control:** Active  **Sample size:** 40  **Description of control group:** Attending the prescribed clinic visits for hypertension at the primary health care facility |
| Isaacs, A. J., Critchley, J. A., Tai, S. S., Buckingham, K., Westley, D., Harridge, S. D. R., ... & Gottlieb, J. M. (2007). Exercise Evaluation Randomised Trial (EXERT): a randomised trial comparing GP referral for leisure centre-based exercise, community-based walking and advice only. *HEALTH TECHNOLOGY ASSESSMENT-SOUTHAMPTON-*, *11*(10). | **Referral pathway:** Direct  **Referral made by:** GP  **Link worker background:**  Qualified instructors | **Setting:** Leisure centres  **Sample size:** 317  **Intervention Name:** Fitness for Life”, a leisure centre-based programme  **Intervention Description:** Available classes included aerobics, body conditioning (strengthening), aqua-aerobics (aerobic with strengthening), gymnasium (stationary bikes, rowers, treadmills, steppers, fixed weights), and optional swimming.  **Intervention Deliverer(s):** Qualified instructors  **Intervention Duration:** 2.5 months  **Frequency of Occurrence:** NR  **Time per Occurrence:** NR  **Most Effective Components:** Aerobics, body conditioning  **Least Effective Components:** Optional swimming class  **Setting:** Community  **Sample size:** 311  **Intervention Name:** Walking Partners”, a community-based walking programme  **Intervention Description:** Across 12 borough park locations, 20 walking classes were offered 7 days a week. Each class included a 10-minute warm-up, 30-40 minutes of graded walking (levels 1-5), 10 minutes of strengthening exercises, and a 5-10 minute cool-down, with participants free to choose any class  **Intervention Deliverer(s):** Qualified instructors  **Intervention Duration:** 2.5 months  **Frequency of Occurrence:** Daily  **Time per Occurrence:** 60-70 minutes  **Most Effective Components:** Walking  **Least Effective Components:** Strengthening exercises | **Type of control:** Active  **Sample size:** 315  **Description of control group:** Information on physical activity including local exercise facilities |
| Laroche, H. H., Andino, J., O'Shea, A. M., Engebretsen, B., Rice, S., DeJear Jr, M., ... & Snetselaar, L. (2024). Family-Based Motivational Interviewing and Resource Mobilization to Prevent Obesity: Living Well Together Trial. *Journal of nutrition education and behavior*, *56*(9), 631-642. | **Referral pathway:** Direct  **Referral made by:** Health coaches  **Link worker background:**  No background related to health | **Setting:** Community, home community, home  **Sample size:** 103  **Intervention Name:** Health Coaching and Community Screening  **Intervention Description:** **Behavioural: health coaching:** Motivational interviewing-based health coaching focused on family diet and exercise changes, and linking families to relevant community resources based on their goals.  **Behavioural: community screening:** Screening families for eligibility for community resources addressing basic needs such as shelter, food, and health insurance  **Intervention Deliverer(s):** Health coaches  **Intervention Duration:** 10.5 months  **Frequency of Occurrence:** Every 6-7 weeks  **Time per Occurrence:** NR  **Most Effective Components:** Motivational interviewing focus on family diet and exercise change and connection with community resources specific to goals set.  **Least Effective Components:** Handouts and videos | **Type of control:** Passive  **Sample size:** 105  **Description of control group:** Screening for community resources that families may be eligible for to receive help with basic needs including shelter, food, health insurance etc. |
| Wagner, J. A., Bermúdez-Millán, A., Buckley, T. E., Buxton, O. M., Feinn, R. S., Kong, S., ... & Scully, M. F. (2023). Community-based diabetes prevention randomized controlled trial in refugees with depression: effects on metabolic outcomes and depression. *Scientific reports*, *13*(1), 8718. | **Referral pathway:** Indirect  **Referral made by:** CHWs  **Link worker background:**  Lay health workers | **Setting:** Community  **Sample size:** 70  **Intervention Name:** Eat, Walk, Sleep  **Intervention Description:** Eat, Walk, Sleep (EWS) is a trauma-informed, cardiometabolic education curriculum, rooted in Buddhist health concepts, designed for Community Health Educators (CHEs) to deliver to learners with low literacy and numeracy. EWS targets eating no more than one small bowl of (brown) rice per meal, walking at least 30 minutes daily (6 days/week), and achieving 7-9 hours of restful sleep nightly, with these goals being progressively implemented through session-to-session goal setting.  **Intervention Deliverer(s):** CHEs and CHWs  **Intervention Duration:** 12 months  **Frequency of Occurrence:** Weekly  **Time per Occurrence:** NR  **Most Effective Components**: 1 small bowl of (brown) rice per meal, walking at least 30 min per day 7-9 hours’ sleep  **Least Effective Components:** Not clear | **Type of control:** Passive  **Sample size:** 53  **Description of control group:**  Participants who were assigned to social services were assessed for any social service needs such as food or housing assistance, referral to a healthcare provider, tax preparation, or citizenship applications. |
| Ugarte, D. A., Lin, J., Qian, T., & Young, S. D. (2022). An online community peer support intervention to promote COVID-19 vaccine information among essential workers: a randomized trial. *Annals of medicine*, *54*(1), 3078-3083. | **Referral pathway:** Indirect  **Referral made by:**  Mental health providers  **Link worker background:**  Peer leaders | **Setting:** Online  **Sample size:** 150  **Intervention Name:** Facebook group with peer leaders  **Intervention Description:** Over six weeks (April 5 to May 17, 2020), intervention participants were assigned to peer leader-supported online groups (two peer leaders per participant), while control participants were in similar groups without peer leaders. Participants were instructed to use the group as desired and continue normal Facebook use, unaware of the peer leader identities within their group. All online community features (wall posts, direct messaging, chat) were available. Peer leaders were trained and guided on weekly topics, aiming to communicate with participants each week and freely post/comment  **Intervention Deliverer(s):** NA  **Intervention Duration:** 6 weeks  **Frequency of Occurrence:** Weekly  **Time per Occurrence:** 30 minutes to 3 hours  **Most Effective Components**: Communicate with participants each week  **Least Effective Components:** Not clear | **Type of control:** Passive  **Sample size:** 150  **Description of control group:** Facebook group without peer leaders |
| Islam, N. S., Wyatt, L. C., Ali, S. H., Zanowiak, J. M., Mohaimin, S., Goldfeld, K., ... & Trinh-Shevrin, C. (2023). Integrating community health workers into community-based primary care practice settings to improve blood pressure control among South Asian immigrants in New York City: results from a randomized control trial. *Circulation: Cardiovascular Quality and Outcomes*, *16*(3), e009321. | **Referral pathway:** Indirect  **Referral made by:**  CHWs  **Link worker background:**  NR | **Setting:** Primary care practice offices and community spaces  **Sample size:** 159  **Intervention Name:** Community-Based Primary Care Practice Settings to Improve Blood Pressure Control  **Intervention Description:**  Treatment participants received 4 additional group education sessions and individualized health coaching over a 6-month period.  **Intervention Deliverer(s):** CHWs  **Intervention Duration:** 5 months  **Frequency of Occurrence:** Bi-weekly  **Time per Occurrence:** 60 to 90 minutes  **Most Effective Components**: Group education sessions  **Least Effective Components:** Nutrition | **Type of control:** Passive  **Sample size:** 144  **Description of control group:**  Control group participants were instructed to seek care as usual after the first session |
| Heisler, M., Lapidos, A., Kieffer, E., Henderson, J., Guzman, R., Cunmulaj, J., ... & Ayanian, J. Z. (2022). Impact on health care utilization and costs of a Medicaid community health worker program in Detroit, 2018–2020: a randomized program evaluation. *American Journal of Public Health*, *112*(5), 766-775. | **Referral pathway:** Indirect  **Referral made by:** CHWs  **Link worker background:**  CHWs underwent training by a trainer from the Detroit Health Department (R. G.), under a contract with the Michigan Community Health Worker Alliance | **Setting:** Community  **Sample size:** 1782  **Intervention Name:** Community health worker (CHW) program  **Intervention Description:** (1) conducting an initial comprehensive health, behavioural, and social needs assessment; (2) developing an individualized action plan; and (3) linking members to necessary services  **Intervention Deliverer(s):** CHWs  **Intervention Duration:** 12 months  **Frequency of Occurrence:** Monthly  **Time per Occurrence:** NR  **Most Effective Components**: Comprehensive health, behavioural, and social needs assessment  **Least Effective Components:** Linking members to necessary services | **Type of control:** Passive  **Sample size:** 1377  **Description of control group:**  Usual care |
| Rovner, B. W., Casten, R., Chang, A. M., Hollander, J. E., Leiby, B. E., Nightingale, G., ... & Rising, K. (2023). Interprofessional intervention to reduce emergency department visits in Black individuals with diabetes. *Population Health Management*, *26*(1), 46-52. | **Referral pathway:** Indirect  **Referral made by:** CHWs  **Link worker background:**  NR | **Setting:** Community and home  **Sample size:** 98  **Intervention Name:**  DM I-TEAM (Diabetes Interprofessional Team to Enhance Adherence to Medical Care)  **Intervention Description:** Home-based multidisciplinary behavioural intervention that integrates care from a community health worker (CHW), the participant's primary care physician (PCP), a DM nurse educator, and a clinical pharmacist  **Intervention Deliverer(s):** CHWs  **Intervention Duration:** 12 months  **Frequency of Occurrence:** Weekly  **Time per Occurrence:** 90 min  **Most Effective Components**: Diabetes self-care education and behavioural activation  **Least Effective Components:** Medication | **Type of control:** Passive  **Sample size:** 102  **Description of control group:**  usual non-protocol-driven primary care practice |
| Gusi, N., Reyes, M. C., Gonzalez-Guerrero, J. L., Herrera, E., & Garcia, J. M. (2008). Cost-utility of a walking programme for moderately depressed, obese, or overweight elderly women in primary care: a randomised controlled trial. *BMC public health*, *8*, 1-10. | **Referral pathway:** Unclear  **Referral made by:** GP  **Link worker background:**  Qualified exercise leaders who had expertise in fitness testing and supervision of group physical activities | **Setting:** Both primary care & community  **Sample size:** 64  **Intervention Name:** Exercise programme  **Intervention Description:** Each session lasted for 50 min, 3 times/week. It comprised walking alternating with specific following exercises: 5 min for joint mobility, 15 min for brisk-walking, 5 min for strengthening and stretching, 20 min of brisk-walking including 20 foot-steps and 50 hand-claps  **Intervention Deliverer(s):** CHWs  **Intervention Duration:** 6 months  **Frequency of Occurrence:** 3 time/week  **Time per Occurrence:** 50 min each session  **Most Effective Components**: Walking and exercise  **Least Effective Components:** Simple nutrition advice and Socialising within the group was encouraged | **Type of control:** Active  **Sample size:** 63  **Description of control group:**  Routine care and a recommendation of physical activity. |
| Harrison, R. A., Roberts, C., & Elton, P. J. (2005). Does primary care referral to an exercise programme increase physical activity one year later? A randomized controlled trial. *Journal of Public Health*, *27*(1), 25-32. | **Referral pathway:** Unclear  **Referral made by:** GP  **Link worker background:**  Exercise officers | **Setting:** Community  **Sample size:** 157  **Intervention Name:** Exercise programme  **Intervention Description:** Clients received a one-hour consultation with an exercise officer for personalized physical activity advice and information, tailored to their preferences and abilities. All clients were offered a subsidized 12-week leisure pass for council-run facilities and encouraged to attend at least two sessions weekly Information on other community-based activities was also provided.  **Intervention Deliverer(s):** Exercise officers  **Intervention Duration:** 3 months  **Frequency of Occurrence:** 2 time/week  **Time per Occurrence:** 90 min each session  **Most Effective Components**: Physical activity  **Least Effective Components:** Non-leisure centre-based activities | **Type of control:** Active  **Sample size:** 270  **Description of control group:**  Leaflets on the importance of physical activity to health and wellbeing, information on local council-run physical activity facilities, and the telephone number of the Exercise |
| Ma, C., Zhou, W., Tang, Q., & Huang, S. (2018). The impact of group-based Tai chi on health-status outcomes among community-dwelling older adults with hypertension. *Heart & Lung*, *47*(4), 337-344. | **Referral pathway:** Direct  **Referral made by:**  Healthcare professionals  **Link worker background:**  Tai chi-certified physical education teachers | **Setting:** Recreation centre for older people in the community  **Sample size:** 79  **Intervention Name:** Group-based Tai chi training  **Intervention Description:** At a community recreation centre for older people: (i) The simplified 24-form Tai Chi was taught in two 90-minute weekly classes for five weeks (10-minute warm-up, 70-minute instruction/practice, 10-minute cool-down), with participants receiving a free Tai Chi video after the first class. (ii) A participant leader organized exercise sessions at a local park, mentored by trainers twice monthly, with group practice encouraged 3-5 times per week  **Intervention Deliverer(s):** Tai chi-certified physical education teachers  **Intervention Duration:** 5 weeks  **Frequency of Occurrence:** Weekly  **Time per Occurrence:** 90 min each session  **Most Effective Components**: Tai Chi and exercise  **Least Effective Components:** Motions, postures, and movement speed | **Type of control:** Active  **Sample size:** 79  **Description of control group:** Visited doctors once a month and given brochure about education about hypertension, medication use, dietary, exercise, lifestyle changes, and stress reduction |
| Murphy, S. M., Edwards, R. T., Williams, N., Raisanen, L., Moore, G., Linck, P., ... & Moore, L. (2012). An evaluation of the effectiveness and cost effectiveness of the National Exercise Referral Scheme in Wales, UK: a randomised controlled trial of a public health policy initiative. *J Epidemiol Community Health*, *66*(8), 745-753. | **Referral pathway:** Unclear  **Referral made by:** GP  **Link worker background:**  Exercise professionals | **Setting:** Leisure centres  **Sample size:** 1080  **Intervention Name:** The Wales National Exercise Referral Scheme (NERS)  **Intervention Description:** Consultations were based on motivational interview16 principles which facilitated patient-centred achievable goals and included relapse-prevention strategies at 4 and 16 weeks to review goals and encourage attendance. The primary goal was for participants to achieve 30 min of moderate physical activity on at least 5 days per week  **Intervention Deliverer(s):** Exercise professionals  **Intervention Duration:** 12 months  **Frequency of Occurrence:** 5 days/Week  **Time per Occurrence:** 30 min each session  **Most Effective Components**: One-to-one exercise instruction and/or group exercise classes  **Least Effective Components:** Motivational interviews | **Type of control:** Active  **Sample size:** 1080  **Description of control group: L**eaflet highlighting the benefits of exercise, and were given the addresses of local facilities |
| Smith, J. R., Greaves, C. J., Thompson, J. L., Taylor, R. S., Jones, M., Armstrong, R., ... & Abraham, C. (2019). The community-based prevention of diabetes (ComPoD) study: a randomised, waiting list controlled trial of a voluntary sector-led diabetes prevention programme. *International Journal of Behavioral Nutrition and Physical Activity*, *16*, 1-14. | **Referral pathway:** Unclear  **Referral made by:** GP  **Link worker background:**  Voluntary sector providers | **Setting:** Local community venues  **Sample size:** 315  **Intervention Name:** LWTC programme  **Intervention Description:** It comprised four, weekly, 2- hour sessions in groups of up to 12 participants (plus accompanying partners/supporters where desired). There were planned support contacts at 2, 3, 6, 9, and 12 months. The programme delivered four education sessions (covering pre-diabetes & healthy lifestyle, healthy eating, physical activity, and positive mental health & wellbeing). The programme also offered 5 hours of additional classes of choice (e.g. exercise sessions, cooking classes, walk in local communities, visits to the provider’s gym)  **Intervention Deliverer(s):** Voluntary sector providers  **Intervention Duration:** 12 months  **Frequency of Occurrence:** 2 days/Week  **Time per Occurrence:** 120 min each session  **Most Effective Components**: Education sessions (covering pre-diabetes & healthy lifestyle, healthy eating, physical activity, and positive mental health & wellbeing)  **Least Effective Components:** Exercise sessions, cooking classes, walk in local communities, visits to the provider’s gym | **Type of control:** Passive  **Sample size:** 157  **Description of control group:**  Participants went onto a six-month waiting list for the programme whilst routine care was continued from their GP involving minimal or no follow-up related to their diabetes risk |
| Taylor, A. H., Doust, J., & Webborn, N. (1998). Randomised controlled trial to examine the effects of a GP exercise referral programme in Hailsham, East Sussex, on modifiable coronary heart disease risk factors. *Journal of Epidemiology & Community Health*, *52*(9), 595-601. | **Referral pathway:** Direct  **Referral made by:** GP  **Link worker background:**  Leisure centre staff | **Setting:** Community health centres and a leisure centre  **Sample size:** 97  **Intervention Name:** Exercise programme  **Intervention Description:** Patients received a prescription card outlining referral details, recommended exercise intensity (three levels), and prohibited activities. They were instructed to present this at Hailsham Lagoon Leisure Centre (East Sussex) to book an introductory session for a 10-week program (up to 20 half-price sessions at £1.30 each). The initial session involved a brief lifestyle assessment, exercise goal discussion, health measurements (blood pressure, weight, height), and guidance on gym equipment and record cards. Patients were encouraged to gradually increase exercise duration and intensity, with supervision available on request during informal weekday sessions (9 am-5 pm, typically up to one hour). Formal components were limited to mid- and end-of-program individual assessments, while leisure centre staff tracked attendance  **Intervention Deliverer(s):** Leisure centre staff  **Intervention Duration:** 2.5 months  **Frequency of Occurrence:** Weekly  **Time per Occurrence:** 60 min each session  **Most Effective Components**: Exercise perceptions and goals, and advice on use of the cycle ergometers, rowing machines, treadmills, and stair climbing machines  **Least Effective Components:** BP, height and weight measurement | **Type of control:** Active  **Sample size:** 45  **Description of control group:**  Leaflets included education on preventing coronary heart disease but no specific advice to change the lifestyle was given |
| Clarke, M., Clarke, S. J., & Jagger, C. (1992). Social intervention and the elderly: a randomized controlled trial. *American Journal of Epidemiology*, *136*(12), 1517-1523. | **Referral pathway:** Indirect  **Referral made by:** CHWs  **Link worker background:**  Lay community-based health worker, training and experience not specified | **Setting:** Community  **Sample size:** 261  **Intervention Name:** Social intervention  **Intervention Description:** Social and social services (arranging visits to another elderly person or outings with voluntary organizations; Meals on Wheels; home help); financial (liaising with local administrative offices for rates, benefits, or collecting pensions); housing (installing safety chains and spy holes onto doors, arranging for volunteers to do gardening or decorating); nursing (referral for assessment for a bath nurse or requesting advice from the continence nurse); and medical (assistance in making an appointment to see the family doctor or informal liaison/ discussion with the general practitioner)  **Intervention Deliverer(s):** Lay community-based health worker  **Intervention Duration:** 24 months  **Frequency of Occurrence:** NR  **Time per Occurrence:** NR  **Most Effective Components**: Individual packages of support that aimed at enhanced social contacts  **Least Effective Components:** Making an appointment to see the family doctor | **Type of control:** Passive  **Sample size:** 262  **Description of control group:** Usual care |
| Grant, C., Goodenough, T., Harvey, I., & Hine, C. (2000). A randomised controlled trial and economic evaluation of a referrals facilitator between primary care and the voluntary sector. *Bmj*, *320*(7232), 419-423. | **Referral pathway:** Direct  **Referral made by: GP**  **Link worker background:**  Lay ‘referral facilitator’ trained and employed by a community organisation | **Setting:** Primary care practice  **Sample size:** 90  **Intervention Name:** Amalthea group  **Intervention Description:** The 10-week ADA education sessions were tailored to the participants’ needs, such as knowledge, identified problems, goals, and level of progress.  **Intervention Deliverer(s):** Lay ‘referral facilitator’ trained and employed by a community organisation.  **Intervention Duration:** 1 month  **Frequency of Occurrence:** NR  **Time per Occurrence:** NR  **Most Effective Components**: Not clear  **Least Effective Components:** Not clear | **Type of control:** Passive  **Sample size:** 71  **Description of control group:** Usual care |
| Babamoto, K. S., Sey, K. A., Camilleri, A. J., Karlan, V. J., Catalasan, J., & Morisky, D. E. (2009). Improving diabetes care and health measures among hispanics using community health workers: results from a randomized controlled trial. *Health Education & Behavior*, *36*(1), 113-126. | **Referral pathway:** Indirect  **Referral made by:**  GP/nurse practitioners  **Link worker background:**  Three with a high school degree and full-time bilingual Hispanic CHWs who had diabetes or had experienced it through a family member or friend | **Setting:** Home and clinic  **Sample size:** 75  **Intervention Name:** CHW intervention  **Intervention Description:** The 10-week ADA education sessions were tailored to the participants’ needs, such as knowledge, identified problems, goals, and level of progress.  **Intervention Deliverer(s):** CHWs  **Intervention Duration:** 6 months  **Frequency of Occurrence:** Multiple sessions  **Time per Occurrence:** NR  **Most Effective Components**: Diabetes education and monitoring services and telephonic calls for monitoring  **Least Effective Components:** Not clear  **Case Management**  **Setting:** Home and clinic  **Sample size:** 54  **Intervention Name:** Case Management  **Intervention Description:** Received diabetes care and education in the clinic setting from two linguistically competent and culturally sensitive registered nurses, as an adjunct to standard provider care  **Intervention Deliverer(s):** Case managers  **Intervention Duration:** 6 months  **Frequency of Occurrence:** Monthly  **Time per Occurrence:** NR  **Most Effective Components**: Patient assessment, development of a case management treatment plan incorporating provider treatment, coordination and referral of community resources  **Least Effective Components:** Not clear | **Type of control:** Active  **Sample size:** 60  **Description of control group:**  Standard Provider Care. Patients assigned to the standard provider care group received only standardized clinical care by physicians and nurse practitioners, without case management or CHW services |
| Lamb, S. E., Bartlett, H. P., Ashley, A., & Bird, W. (2002). Can lay-led walking programmes increase physical activity in middle aged adults? A randomised controlled trial. *Journal of Epidemiology & Community Health*, *56*(4), 246-252. | **Referral pathway:** Direct  **Referral made by:**  GP/nurse practitioners  **Link worker background:**  Lay volunteers | **Setting:** Community  **Sample size:** 131  **Intervention Name:** Health walks  **Intervention Description:** Participants received verbal and written information about a local health walks program (accompanied walks led by volunteers at various times, and self-guided walks with provided packs). Up to three phone calls encouraged participation, and a walk pack with promotional flyers was mailed to each person. Walks were free. The key message was to achieve at least 120 minutes of moderate-intensity activity weekly through enjoyable and convenient options like walking, swimming, racquet sports, or aerobics  **Intervention Deliverer(s):** Lay volunteers  **Intervention Duration:** 1 year  **Frequency of Occurrence:** Weekly  **Time per Occurrence:** ~120 min  **Most Effective Components:** Health walks  **Least Effective Components:** Not clear | **Type of control:** Active  **Sample size:** 129  **Description of control group:** Advice group: Participants attended a 30-minute, physiotherapist-led group advice session (10-20 people) in primary care. The session covered exercise benefits, recommended activity levels (≥120 minutes/week moderate intensity), tips for starting and maintaining enjoyable physical activity (e.g., swimming, racquet sports, aerobics, walking), and the indicator of moderate intensity (slight sweat/breathlessness). Participants were encouraged to ask questions and share experiences. General written guidance supplemented the seminar, but the control group was not referred to the health walks scheme |
| Kiely, B., Hobbins, A., Boland, F., Clyne, B., Galvin, E., Byers, V., ... & Smith, S. M. (2024). An exploratory randomised trial investigating feasibility, potential impact and cost effectiveness of link workers for people living with multimorbidity attending general practices in deprived urban communities. *BMC Primary Care*, *25*(1), 233. | **Referral pathway:** Indirect  **Referral made by:**  Link worker  **Link worker background:**  Non-health or social care professional | **Setting:** GP practice and remote  **Sample size:** 123  **Intervention Name:** LinkMM intervention  **Intervention Description:** The **LinkMM** is a complex intervention with several key components: training for link workers and GPs, mapping of local community resources, participant meetings with follow-up, and financial support for practices  **Intervention Deliverer(s):** Link worker  **Intervention Duration:** 1 month  **Frequency of Occurrence:** Not clear  **Time per Occurrence:** over a month  **Most Effective Components:** Not clear  **Least Effective Components:** Not clear | **Type of control:** Passive  **Sample size:** 117  **Description of control group:**  Usual care: Usual care received from their GP.  Participants were invited to a one-off meeting with the link worker to identify their needs and received a list of suggested resources and activities. |

**Supplementary table 14: Ongoing trials**

| **Study title** | **Clinical trial ID** | **Estimated Study completion date** |
| --- | --- | --- |
| Randomised controlled trial of the Community Navigator programme to reduce loneliness and depression for adults with treatment-resistant depression in secondary community mental health services: trial protocol | ISRCTN13205972 | 31/12/2025 |
| Comparing cognitive behavioral therapy and social prescribing in patients with loneliness on long-term opioid therapy to reduce opioid misuse: protocol for a randomized controlled trial | NCT06285032 | 01/05/2026 |
| Social Prescribing to Improve Adherence and Outcomes in Women With Heart Failure | NCT06628973 | 31/12/2028 |
| Testing the Effectiveness of a Nature-based Social Intervention on Loneliness Among Lonely Adults in Barcelona - a Randomized Controlled Trial | NCT05507684 | 28/02/2028 |
| Social Prescribing in Sweden (SPiS)- An Interventional Research Project Evaluating a Swedish Model | NCT04336553 | 31/12/2025 |
| Social Prescribing and Relationship Cognitive Strategies (SPARCS) | NCT06656975 | 01/07/2026 |
| OUTdoor Swimming as a nature-based Intervention for DEpression (OUTSIDE): study protocol for a feasibility randomised control trial comparing an outdoor swimming intervention to usual care for adults experiencing mild to moderate symptoms of depression | ISRCTN90851983 | Completed* |
| Rationale and design of the Linking education, produce provision, and community referrals to improve diabetes care (LINK) study | NCT05472441 | 01/10/2026 |
| An Effectiveness-implementation Hybrid Study of Social Prescribing in a Singapore Community Hospital Setting | NCT04840420 | Unknown |
| The Role of Intermediaries in Connecting Individuals to Local Physical Activity - Study Protocol | NCT06260995 | 30/03/2025* |
| Testing a Nature-based Social Intervention on Loneliness: the RECETAS-PRG Trial | NCT05522140 | 31/10/2025 |
| Protocol for the Houston Hospital-based violence intervention program | NCT06263647 | 31/05/2028 |
| Social prescribing to improve health and well-being of patients presenting with non-medical health related social needs in primary care: Study protocol of a multi-center randomized controlled pragmatic feasibility trial | DRKS00034654 | Ongoing |
| A community health worker-delivered intervention (STEPS) to support chronic pain self-management among older adults in an underserved urban community: protocol for a randomized trial | NCT05278234 | 01/04/2026 |
| Effectiveness Evaluation of Social Prescribing | NCT07029334 | 01/03/2027 |
| CAMHS and Social Prescribing Applications (CASPA) | NCT07143383 | 31/05/2027 |
| Arts-based Social Prescribing for Mental Health (AoP-II) | NCT07137572 | 01/2026 |
| EPIC-ND: a multisite, randomised controlled trial evaluating the effectiveness of social prescribing to address the unmet social needs of children with a neurodisability and their parent/carers | ACTRN12625000324415 | 01/05/2028 |
| Decreasing Loneliness to Optimize Pain Care (DLoop) | NCT06285032 | 01/05/2026 |

***:** Not updated/results not posted

**Supplementary table 15: Outcomes not contributing to meta-analyses**

| **Study ID** | **Outcome** | **Primary/secondary/uncategorised** | **Outcome measurement** | **Assessment time** | **Outcome assessed by** | **Outcome type (continuous/binary)** | **Narrative finding(s)** |
| --- | --- | --- | --- | --- | --- | --- | --- |
| Harrison 2005 | Moderate/vigorous physical activity | Uncategorised | 7-Day Physical Activity Recall (7dPAR) questionnaire | 3 months | Self-reported via post | Binary | NR |
| Harrison 2005 | Physical activity | Secondary | 7-Day Physical Activity Recall (7dPAR) questionnaire | 6 months | Self-reported via post | Binary | Intervention group found to be more physically active than control group |
| Harrison 2005 | Physical activity | Uncategorised | 7-Day Physical Activity Recall (7dPAR) questionnaire | 9 months | Self-reported via post | Binary | No significant difference |
| Harrison 2005 | Physical activity (MVPA) | Primary | 7-Day Physical Activity Recall (7dPAR) questionnaire | 12 months | Self-reported via post | Binary | No significant difference |
| Harrison 2005 | Satisfaction | Uncategorised | NA | 3 months | Self-reported via post | Binary | The intervention increased satisfaction with information |
| Ma 2018 | Social support | Uncategorised | NA | 24 weeks | NR | Continuous | Greater improvements showed in intervention |
| Smith 2019 | N (%) > 3% weight loss | Secondary | Tanita scales | 6 months | Trained researcher | Binary | Greater reduction in intervention group |
| Smith 2019 | N (%) > 5% weight loss | Secondary | Tanita scales | 6 months | Trained researcher | Binary | Greater reduction in intervention group |
| Smith 2019 | Health rating | Secondary | EuroQol EQ-5D | 6 months | Self reported | Continuous | Greater improvement in intervention group |
| Smith 2019 | Dietary behaviour fat subscale scores | Secondary | 27-item Fat and Fibre questionnaire | 6 months | Self reported | Continuous | Greater improvement in intervention group |
| Smith 2019 | Dietary behaviour fibre subscale score | Secondary | 27-item Fat and Fibre questionnaire | 6 months | Self reported | Continuous | Greater improvement in intervention group |
| Smith 2019 | Mean (SD) life satisfaction rating (0–10, higher better) | Secondary | Life satisfaction rating scale | 6 months | Self reported | Continuous | No significant difference |
| Smith 2019 | Mean (SD) mental well-being scores (0–35, higher better) | Secondary | the Short Warwick-Edinburgh Mental Well-being Scale | 6 months | Self reported | Continuous | No significant difference |
| Smith 2019 | Adverse events | Uncategorised | NA | 6 months | Self reported | Binary | Four nonserious adverse events reported by intervention group participants potentially related to the intervention, all short-term injuries stemming from increased exercise (pelvic pain, lower back pain, aggravation of existing sciatica shoulder injury) that resolved on their own or with treatment. There were none reported in the control group |
| Taylor 1998 | BMI | Uncategorised | NA | 16 weeks | Trained staff | Continuous | Significantly reduced in intervention group |
| Taylor 1998 | BMI | Uncategorised | NA | 26 weeks | Trained staff | Continuous | No significant difference |
| Taylor 1998 | SBP | Uncategorised | Sphygmomanometer | 16 weeks | Trained staff | Continuous | No significant difference |
| Taylor 1998 | DBP | Uncategorised | Sphygmomanometer | 16 weeks | Trained staff | Continuous | No significant difference |
| Taylor 1998 | DBP | Uncategorised | Sphygmomanometer | 26 weeks | Trained staff | Continuous | No significant difference |
| Taylor 1998 | Moderate physical activity | Uncategorised | Blair’s seven day recall method | 8 weeks | Trained staff | Continuous | No significant difference |
| Taylor 1998 | Moderate physical activity | Uncategorised | Blair’s seven day recall method | 16 weeks | Trained staff | Continuous | No significant difference |
| Taylor 1998 | Moderate physical activity | Uncategorised | Blair’s seven day recall method | 26 weeks | Trained staff | Continuous | No significant difference |
| Taylor 1998 | Vigorous physical activity | Uncategorised | Blair’s seven day recall method | 8 weeks | Trained staff | Continuous | No significant difference |
| Taylor 1998 | Vigorous physical activity | Uncategorised | Blair’s seven day recall method | 16 weeks | Trained staff | Continuous | No significant difference |
| Taylor 1998 | Vigorous physical activity | Uncategorised | Blair’s seven day recall method | 26 weeks | Trained staff | Continuous | No significant difference |
| Taylor 1998 | Vigorous physical activity | Uncategorised | Blair’s seven day recall method | 37 weeks | Trained staff | Continuous | No significant difference |
| Taylor 1998 | Energy expended | Uncategorised | NA | 8 weeks | Trained staff | Continuous | Expended more energy by people in intervention group |
| Taylor 1998 | Energy expended | Uncategorised | NA | 16 weeks | Trained staff | Continuous | Expended more energy by people in intervention group |
| Taylor 1998 | Energy expended | Uncategorised | NA | 26 weeks | Trained staff | Continuous | No significant difference |
| Taylor 1998 | Energy expended | Uncategorised | NA | 37 weeks | Trained staff | Continuous | No significant difference |
| Clarke 1992 | Perceived health status | Uncategorised | Activities of Daily Living index | 3 years | Field worker | Binary | No significant difference |
| Clarke 1992 | Social contact score | Uncategorised | Tunstall, based on the number of personal contacts during the week and month prior to interview, including both nonfamily and family contacts | 3 years | Field worker | Binary | No significant difference |
| Clarke 1992 | Physical health status | Uncategorised | Activities of Daily Living index | 3 years | Field worker | Binary | No significant difference |
| Clarke 1992 | Deaths | Uncategorised | NA | 3 years | Self reported | Binary | No significant difference |
| Clarke 1992 | Perceived loneliness | Uncategorised | NA | 3 years | Self reported | Binary | No significant difference |
| Grant 2000 | Anxiety | Primary | HADS | 1 month | Self reported | Continuous | The Amalthea group showed significantly greater improvements in anxiety |
| Grant 2000 | Depression | Primary | HADS | 1 month | Self reported | Continuous | No significant difference |
| Grant 2000 | Pain | Secondary | The Dartmouth COOP/WONCA functional health assessment charts | 1 month | Self reported | Continuous | Significantly reduced in intervention group |
| Grant 2000 | Pain | Secondary | The Dartmouth COOP/WONCA functional health assessment charts and the delighted terrible faces scale | 4th month | Self reported | Continuous | Significantly reduced in intervention group |
| Grant 2000 | Cost of contacts with the primary healthcare team and Amalthea Project | Secondary | The Dartmouth COOP/WONCA functional health assessment charts and the delighted terrible faces scale | 4 months | NR | $ | The mean cost was significantly greater in the Amalthea arm than the general practitioner care arm (£153 v £133, P = 0.025). |
| Grant 2000 | Total cost of primary healthcare team contacts, prescribing, and referrals (£) | Secondary | The Dartmouth COOP/WONCA functional health assessment charts and the delighted terrible faces scale | 4 months | NR | $ | NA |
| Grant 2000 | Daily activities | Secondary | The Dartmouth COOP/WONCA functional health assessment charts and the delighted terrible faces scale | 1 and 4th month | Self reported | Continuous | Intervention group showed greater improvement |
| Grant 2000 | Social activities | Secondary | The Dartmouth COOP/WONCA functional health assessment charts and the delighted terrible faces scale | 1 and 4th month | Self reported | Continuous | Intervention group showed greater improvement |
| Grant 2000 | Change in health | Secondary | The Dartmouth COOP/WONCA functional health assessment charts and the delighted terrible faces scale | 1 and 4th month | Self reported | Continuous | Intervention group showed greater improvement |
| Grant 2000 | Feeling | Secondary | The Dartmouth COOP/WONCA functional health assessment charts and the delighted terrible faces scale | 1 and 4th month | Self reported | Continuous | Intervention group showed greater improvement |
| Grant 2000 | Skipping meals a few times a month or less | Secondary | Skipping meals a few times a month or less | 12 months | Self reported | Binary | Intervention group showed greater improvement |
| Grant 2000 | Preparing meals mostly at home | Uncategorised | Preparing meals mostly at home | 12 months | Self reported | Binary | Intervention group showed greater improvement |
| Babamoto 2009 | Physical activity | Uncategorised | Exercising at least 3 days a week | 6 months | CHWs | Binary | Physical activity improved significantly across all groups except the case management group |
| Babamoto 2009 | BMI | Uncategorised | NA | 6 months | NR | Continuous | Greater improvement in intervention group |
| Babamoto 2009 | Two or more servings of fresh fruit per day | Uncategorised | NA | 6 months | NR | Binary | Greater improvement in intervention group |
| Babamoto 2009 | Two or more servings of fresh vegetables per day | Uncategorised | NA | 6 months | NR | Binary | Greater improvement in intervention group |
| Babamoto 2009 | Two or more servings of fatty foods per day | Uncategorised | NA | 6 months | NR | Binary | Greater improvement in intervention group |
| Babamoto 2009 | Diabetes Knowledge | Uncategorised | Diabetes Knowledge Questionnaire | 6 months | NR | Continuous | Greater improvement in intervention group |
| Kangovi 2017 | Mental health | Secondary | Self rated | 6 months | Self reported | Continuous | CHW support showed greater improvements in mental health (2.3 vs –0.2; P = .008) |
| Kangovi 2017 | Supportive of disease self-management | Secondary | Self rated | 6 months | Self reported | Binary | Greater improvement in intervention group |
| Kangovi 2017 | SBP | primary | NR | 6 months |  | Continuous | Greater improvement in intervention group |
| Kangovi 2017 | HbA1C | primary | NR | 6 months |  | Continuous | Greater improvement in intervention group |
| Kangovi 2017 | BMI | Primary | NR | 6 months |  | Continuous | Greater improvement in intervention group |
| Kangovi 2017 | All cause hospitalisation | Secondary | NR | 6 months |  | Binary | No significant difference |
| Kangovi 2017 | Patient activation | Secondary | NR | 6 months |  | Continuous | No significant difference |
| Kangovi 2017 | Self-rated physical health | Secondary | SF-12 | 6 months | Self reported | Continuous | No significant difference |
| Kangovi 2017 | Achievement (yes or no) of chronic disease management goals | Primary | NA | 6 months | Self reported | Binary | CHW support showed greater improvements |
| Kangovi 2017 | Cigarettes per day | Primary | NA | 6 months | Self reported | Binary | Greater improvement in intervention group |
| Kangovi 2017 | Hospital days | Secondary | NA | 12 months |  | Binary | No significant difference |
| Kangovi 2018 | HbA1C | Secondary | Blood | 6 months | RA | Continuous | No significant difference |
| Kangovi 2018 | HbA1C | Secondary | Blood | 9 months | RA | Continuous | No significant difference |
| Kangovi 2018 | BMI | Secondary | NA | 6 months | RA | Continuous | No significant difference |
| Kangovi 2018 | Change in self-rated mental health | Primary | SF-12v2 Health Survey Mental Component Summary instrument | 6 months | RA | Continuous | No significant difference |
| Kangovi 2018 | Change in self-rated mental health | Primary | SF-12v2 Health Survey Mental Component Summary instrument | 9 months | RA | Continuous | No significant difference |
| Kangovi 2018 | Change in patient activation | Secondary | NA | 6 months | RA | Continuous | Greater improvement in patient activation in intervention group |
| Kangovi 2018 | Change in patient activation | Secondary | NA | 9 months | RA | Continuous | Greater improvement in patient activation in intervention group |
| Kangovi 2018 | Cigarettes per day | Secondary | NA | 6 months | RA | Continuous | No significant difference |
| Kangovi 2018 | Cigarettes per day | Secondary | NA | 9 months | RA | Continuous | No significant difference |
| Kangovi 2018 | All-cause hospitalization | Secondary | NA | 6 months | RA | Count | Lower mean number of hospitalisations in intervention group |
| Kangovi 2018 | Quality of care | Secondary | NA | 6 months | RA | Binary | Highest quality of care reported in intervention group |
| Kangovi 2018 | Length of stay | Secondary | NA | 9 months | RA | Binary | Spent fewer total days in the hospital at 6 months in intervention group |
| Kangovi 2018 | 30-day readmissions | Secondary | NA | 9 months | NR | Binary | Lower odds of repeat hospitalizations and 30-day readmissions in intervention group |
| Spencer 2013 | Diabetes-related emotional distress | Primary | The Problem Areas in Diabetes Scale | 6 & 12 months | NR | Continuous | Greater improvement in intervention group |
| Spencer 2013 | Depressive severity | Primary | Patient Health Questionnaire (PHQ) scales | 6 & 12 months | NR | Continuous | Greater improvement in intervention group |
| Spencer 2013 | HbA1C | Primary | Blood | 6 months | NR | Continuous | Significantly reduced in intervention group |
| Spencer 2013 | Self-management score | Uncategorised | CDC’s Behavioural Risk Factor Surveillance System | 6 months | NR | Continuous | Greater improvement in intervention group |
| Spencer 2013 | Meeting physical activity guidelines | Uncategorised | NA | 6 months | NR | Continuous | No significant difference |
| Spencer 2013 | Daily fruit and vegetable servings | Uncategorised | NA | 6 months | NR | Continuous | No significant difference |
| Spencer 2018 | HbA1C | Primary | Blood | 6, 12 and 18 months | NR | Continuous | CHW+PL participants maintained HbA1c improvements at 12 and 18 months, and CHW-only participants maintained improvements in diabetes distress at 12 and 18 months |
| Spencer 2018 | Diabetes distress | Secondary | Patient Health Questionnaire-9 | 6, 12 and 18 months | NR | Continuous | Greater improvement in intervention group |
| Spencer 2018 | LDL | Secondary | Cholestech LDX | 6, 12 and 18 months | NR | Continuous | No significant difference |
| Spencer 2018 | HDL | Secondary | Cholestech LDX | 6, 12 and 18 months | NR | Continuous | No significant difference |
| Spencer 2018 | TC | Secondary | Cholestech LDX | 6, 12 and 18 months | NR | Continuous | No significant difference |
| Spencer 2018 | BMI | Uncategorised | kg/m^2^ | 6, 12 and 18 months | NR | Continuous | No significant difference |
| Spencer 2018 | SBP | Secondary | Welch Allyn Speidel & Keller sphygmomanometer | 6, 12 and 18 months | NR | Continuous | No significant difference |
| Spencer 2018 | DBP | Secondary | Welch Allyn Speidel & Keller sphygmomanometer | 6, 12 and 18 months | NR | Continuous | No significant difference |
| Spencer 2018 | Depressive symptoms | Secondary | Patient Health Questionnaire-9 | 6, 12 and 18 months | NR | Continuous | Greater improvement in intervention group |
| Gray 2021 | HbA1C | Primary | Blood | 12 months | CHWs | Continuous | No difference between intervention and control group participants |
| Gray 2021 | General diet recommendations more days per week | Secondary | NA | 12 months | CHWs | Continuous | Greater improvement in intervention group |
| Gray 2021 | Total cholesterol to HDL ratio (SD) | Secondary | Cholesterol Test Panel | 12 months | CHWs | Continuous | No significant difference |
| Gray 2021 | ED visits past year | Secondary | NR | 12 months | CHWs | Continuous | No significant difference |
| Gray 2021 | Hospitalized in past year | Secondary | NR | 12 months | CHWs | Continuous | No significant difference |
| Gray 2021 | ED visit in past year, n (%) | Secondary | NR | 12 months | CHWs | Binary | No significant difference |
| Gray 2021 | Outpatient clinic visits in past year, n | Secondary | NR | 12 months | CHWs | Continuous | Less number of visits in intervention group compared to control group |
| Gray 2021 | Social burden subscale of Diabetes-39 score | Secondary | Diabetes-39 scales | 12 months | CHWs | Continuous | Decrease in reported social burden in the intervention group relative to the control group |
| Gray 2021 | Diabetes-specific quality-of-life scales | Secondary | Diabetes-specific HRQOL | 12 months | CHWs | Continuous | No significant difference |
| Ramirez 2020 | HRQOL | Primary | 27-item Functional Assessment of Cancer Therapy General (FACT-G) scale | 3 months | Assessor | Continuous | PN-LCNS demonstrated a significant improvement in HRQOL in comparison with PN only for colorectal cancer survivors |
| Ramirez 2020 | HRQOL | Primary | 27-item Functional Assessment of Cancer Therapy General (FACT-G) scale | 6 months | Assessor | Continuous | PN-LCNS demonstrated a significant improvement in HRQOL in comparison with PN only for colorectal cancer survivors |
| Ramirez 2020 | HRQOL | Primary | 27-item Functional Assessment of Cancer Therapy General (FACT-G) scale | 15 months | Assessor | Continuous | PN-LCNS demonstrated a significant improvement in HRQOL in comparison with PN only for colorectal cancer survivors |
| Carrasquillo 2017 | SBP | Primary | NR | 12 months | NR | Continuous | No significant difference |
| Carrasquillo 2017 | HbA1C | Primary | Blood | 12 months | NR | Continuous | Greater improvement in intervention group |
| Carrasquillo 2017 | LDL | Primary | Blood | 12 months | NR | Continuous | No significant difference |
| Bossche 2021 | Emotional support | Primary | Patient-Reported Outcomes Measurement Information System (PROMIS™) | 2 months | Self rated | Continuous | No significant difference |
| Bossche 2021 | Social participation | Primary | Patient-Reported Outcomes Measurement Information System (PROMIS™) | 2 months | Self rated | Continuous | No significant difference |
| Bossche 2021 | Anxiety | Primary | Patient-Reported Outcomes Measurement Information System (PROMIS™) | 2 months | Self rated | Continuous | No significant difference |
| Bossche 2021 | Fear of COVID-19 | Secondary | Patient-Reported Outcomes Measurement Information System (PROMIS™) | 2 months | Self rated | Continuous | No significant difference |
| Patel 2024 | HRQOL | Primary | 27-item Functional Assessment of Cancer Therapy-General | 4 months | RA | Continuous | HRQoL improved in intervention group more than control group |
| Patel 2024 | Patient activation | Secondary | 13-item Patient Activation Measure | 4 months | RA | Continuous | The intervention group had a greater increase in mean scores |
| Patel 2024 | Patient activation | Secondary | 13-item Patient Activation Measure | 12 months | RA | Continuous | The intervention group had a greater increase in mean scores |
| Patel 2024 | Satisfaction With Decision | Secondary | Six-item Satisfaction with Decision scale | 4 months | RA | Binary | Intervention group was more likely to strongly agree with satisfaction with decision |
| Patel 2024 | Satisfaction With Decision | Secondary | Six-item Satisfaction with Decision scale | 12 months | RA | Binary | Intervention group was more likely to strongly agree with satisfaction with decision |
| Patel 2024 | Total Costs of Care | Secondary | NA | 12 months | NA | $ | Intervention group had lower median total costs of care ($67,655 [USD]) than the control group |
| Patel 2024 | Health care use | Secondary | ED visit | 4 months | NA | Binary | Intervention group had lower ED visits than control group |
| Patel 2024 | Health Care use | Secondary | ED visit | 12 months | NA | Binary | Intervention group had lower ED visits than control group |
| Patel 2024 | Health care use | Secondary | Hospital use | 4 months | NA | Binary | Intervention group had lower hospitalisation than control group |
| Patel 2024 | Health care use | Secondary | Hospital use | 12 months | NA | Binary | Intervention group had lower hospitalisation than control group |
| Patel 2024 | Overall survival | Secondary | Deaths | 12 months | NA | Binary | No significant difference |
| Patel 2024 | Advance Care Planning Documentation | Secondary | NA | 4 months | NA | Binary | Greater odds of AD documentation in intervention group |
| Patel 2024 | Advance Care Planning Documentation | Secondary | NA | 12 months | NA | Binary | Greater odds of AD documentation in intervention group |
| Patel 2024 | GOC documentation | Secondary | NA | 4 months | NA | Binary | Greater odds of AD documentation in intervention group |
| Patel 2024 | GOC documentation | Secondary | NA | 12 months | NA | Binary | Greater odds of AD documentation in intervention group |
| Patel 2024 | POLST documentation | Secondary | NA | 4 months | NA | Binary | Greater odds of AD documentation in intervention group |
| Patel 2024 | POLST documentation | Secondary | NA | 12 months | NA | Binary | Greater odds of AD documentation in intervention group |
| McQueen 2024 | HbA1C | Primary | Blood | 12 months | From medical claims data | Continuous | No significant difference |
| McQueen 2024 | Healthcare utilization | Secondary | ED utilization (number of unique ED service claim dates) | 12 months | From medical claims data | Binary | No significant difference |
| McQueen 2024 | Healthcare utilization | Secondary | Hospitalization (number of unique visits based on a managed inpatient calendar linked to inpatient authorizations and claims data ) | 12 months | From medical claims data | Binary | No significant difference |
| McQueen 2024 | Healthcare utilization | Secondary | Wellness visit (any) | 12 months | From medical claims data | Binary | No significant difference |
| McQueen 2024 | Healthcare utilization | Secondary | High ED utilization (>4/year) | 12 months | From medical claims data | Binary | No significant difference |
| McQueen 2024 | Healthcare utilization | Secondary | 90-day rehospitalization | 12 months | From medical claims data | Binary | No significant difference |
| McQueen 2024 | Uncontrolled diabetes outpatient | Secondary | NR | 12 months | RA | Binary | No significant difference |
| McQueen 2024 | Stress | Secondary | Perceived Stress Scale | 12 months | RA | Continuous | No significant difference |
| McQueen 2024 | Diabetes Distress | Secondary | Diabetes Distress Scale (Range: 2–12) | 12 months | RA | Continuous | No significant difference |
| McQueen 2024 | Diabetes Self-Efficacy | Secondary | Diabetes Self-Efficacy (Range: 0–10) | 12 months | RA | Continuous | No significant difference |
| McQueen 2024 | Social Support | Secondary | NR | 12 months | RA | Continuous | No significant difference |
| McQueen 2024 | Diabetes self-management | Secondary | Diabetes self-management (Range 0–70) | 12 months | RA | Continuous | No significant difference |
| McQueen 2024 | Sleep quality in past month | Secondary | Hours of sleep (Range: 0–10.5) | 12 months | RA | Continuous | No significant difference |
| McQueen 2024 | Sleep quality in past month | Secondary | Quality of sleep (Range: 1–4) | 12 months | RA | Continuous | No significant difference |
| McQueen 2024 | Sleep quality in past month | Secondary | Trouble sleeping (Range: 1–4) | 12 months | RA | Continuous | No significant difference |
| McQueen 2024 | HbA1c value change BL to 12 *≥* 0.5% | Secondary | Blood | 12 months | RA | Binary | No significant difference |
| McQueen 2024 | Social needs | Secondary | Food | 12 months | RA | Binary | No significant difference |
| McQueen 2024 | Social needs | Secondary | Transportation | 12 months | RA | Binary | No significant difference |
| McQueen 2024 | Social needs | Secondary | Place to stay | 12 months | RA | Binary | No significant difference |
| McQueen 2024 | Social needs | Secondary | Living space | 12 months | RA | Binary | No significant difference |
| McQueen 2024 | Social needs | Secondary | Neighbourhood safety | 12 months | RA | Binary | No significant difference |
| McQueen 2024 | Social needs | Secondary | Utilities | 12 months | RA | Binary | Required more in intervention group |
| McQueen 2024 | Social needs | Secondary | Unexpected expenses | 12 months | RA | Binary | No significant difference |
| McQueen 2024 | Social needs | Secondary | Personal harm | 12 months | RA | Binary | No significant difference |
| McQueen 2024 | Social needs | Secondary | Childcare | 12 months | RA | Binary | No significant difference |
| Yamashita 2024 | Physical activity | Uncategorised | Steps/day | 3 months | Fitbit Inspire2 | Continuous | No significant difference |
| Yamashita 2024 | Subjective happiness | Uncategorised | Subjective happiness scale | 3 months | RA | Continuous | Greater improvement in intervention group |
| Yamashita 2024 | Civic participation | Uncategorised | Five items: (1) volunteer groups, (2) sports groups, (3) hobby activity, (4) study or cultural group, and (5) skills teaching | 3 months | RA | Continuous | No significant difference |
| Yamashita 2024 | Social cohesion | Uncategorised | Saito’s report | 3 months | RA | Continuous | No significant difference |
| Yamashita 2024 | Reciprocity | Uncategorised | Saito’s report | 3 months | RA | Continuous | No significant difference |
| Watts 2024 | Total lean mass | Secondary | NR | 12 months | NR | Continuous | No significant difference |
| Watts 2024 | Total fat mass | Secondary | NR | 12 months | NR | Continuous | No significant difference |
| Watts 2024 | Total cholesterol | Secondary | Blood | 12 months | NR | Continuous | No significant difference |
| Watts 2024 | HDL cholesterol | Secondary | Blood | 12 months | NR | Continuous | No significant difference |
| Watts 2024 | LDL cholesterol | Secondary | Blood | 12 months | NR | Continuous | No significant difference |
| Watts 2024 | Triglycerides | Secondary | Blood | 12 months | NR | Continuous | No significant difference |
| Watts 2024 | HbA1c | Secondary | Blood | 12 months | NR | Continuous | No significant difference |
| Watts 2024 | Peak oxygen consumption | Primary | VO_2_ peak | 3 months | NR | Continuous | Greater improvement in intervention group |
| Watts 2024 | Peak oxygen consumption | Primary | VO_2_ peak | 12 months | NR | Continuous | Greater improvement in intervention group |
| Watts 2024 | Fluid cognitive composite | Secondary | National Institutes of Health (NIH) Toolbox | 3 months | NR | Continuous | No significant difference |
| Watts 2024 | Fluid cognitive composite | Secondary | National Institutes of Health (NIH) Toolbox | 12 months | NR | Continuous | No significant difference |
| Watts 2024 | Crystallized cognitive composite | Secondary | National Institutes of Health (NIH) Toolbox | 3 months | NR | Continuous | Greater improvement in intervention group |
| Watts 2024 | Crystallized cognitive composite | Secondary | National Institutes of Health (NIH) Toolbox | 12 months | NR | Continuous | Greater improvement in intervention group |
| Joo 2024 | Coping | Secondary | Brief Cope | 12 months | Staff | Continuous | Both groups also experienced decreased active coping |
| Joo 2024 | Self efficacy | Secondary | 10-item General Self-Efficacy Scale | 12 months | Staff | Continuous | Both groups also experienced increased self-efficacy |
| Joo 2024 | Health service use | Secondary | Cornell Service Index | 12 months | Staff | Continuous | No significant difference |
| Joo 2024 | Physical Functioning | Secondary | NA | 12 months | NR | Continuous | No significant difference |
| Joo 2024 | Emotional Functioning | Secondary | NA | 12 months | NR | Continuous | No significant difference |
| Joo 2024 | Social Functioning | Secondary | NA | 12 months | NR | Continuous | No significant difference |
| Nyamathi 2024 | Anxiety | Uncategorised | Generalised Anxiety Disorder −7 | 2 months | NR | Continuous | No significant difference |
| Nyamathi 2024 | Depression | Uncategorised | Patient Health Questionnaire‐9 | 2 months | NR | Continuous | No significant difference |
| Nyamathi 2024 | PTSD symptoms | Uncategorised | PCL‐C | 2 months | NR | Continuous | No significant difference |
| Nyamathi 2024 | Heart Rate Variability | Uncategorised | Resting 5‐min period at week 1 and week 8 sessions by the RN/CHW teams using commercially available, medical grade photodetector (photo plethysmography or PPG) sensor | 2 months | CHWs | Continuous | NR |
| Nyamathi 2024 | Mental Health Status | Uncategorised | Mental Health Inventory 5 | 2 months | NR | Continuous | No significant difference |
| Nyamathi 2024 | Drug and Alcohol Use | Uncategorised | TCU Drug History Form | 2 months | NR | Continuous | No significant difference |
| Nyamathi 2024 | Chronic Physical Diseases | Uncategorised | Self‐reported comorbidity index (SCQ) | 2 months | NR | Binary | No significant difference |
| Nyamathi 2024 | Shelter Stability | Uncategorised | Length of time residing in a shelter versus elsewhere at baseline and 2‐month follow‐up | 2 months | NR | Continuous | NR |
| Nyamathi 2024 | RMSSD | Primary | Root mean square of successive differences between normal heart beats | 2 months | NR | Continuous | No significant difference |
| Nyamathi 2024 | Social support | Uncategorised | Medical Outcome Study (MOS) Social Support Survey | 2 months | NR | Continuous | NR |
| Mbuthia 2024 | DBP | Primary | Omron M1 | 3 months | NR | Continuous | Significantly reduced in intervention group |
| Mbuthia 2024 | BMI | Secondary | CAMRY Mechanical scale, BR9012, Shanghai, China | 3 months | NR | Continuous | No significant difference |
| Mbuthia 2024 | Waist height ratio | Secondary | % | 6 months | NR | Continuous | No significant difference |
| Mbuthia 2024 | BP control | Secondary | *<*140/90 mm Hg | 6 months | NR | Binary | Significantly improved in intervention group |
| Laroche 2024 | BMI | Primary | ELISA kit | 12 months | NR | Continuous | No significant difference |
| Laroche 2024 | Sedentary activity | Primary | NA | 12 months | NR | Continuous | No significant difference |
| Laroche 2024 | Family Nutrition and Physical Activity | Primary | NA | 12 months | NR | Continuous | No significant difference |
| Ugarte 2023 | Online engagement | Secondary | NA | 6 weeks | Self rated | Binary | No significant difference |
| Ugarte 2023 | Requesting e-resources | Secondary | NA | 6 weeks | Self rated | Binary | No significant difference |
| Wagner 2023 | Depression | Primary | Khmer language Hopkins Symptom Checklist | 12 months | RA | Continuous | Significantly reduced in intervention group |
| Wagner 2023 | HbA1c | Primary | Quest laboratory using direct enzymatic assay | 12 months | NA | Continuous | Significantly reduced in intervention group |
| Wagner 2023 | HbA1c | Primary | Quest laboratory using direct enzymatic assay | 15 months | NA | Continuous | Significantly reduced in intervention group |
| Wagner 2023 | Insulin resistance | Primary | Fasting glucose and insulin values | 12 months | NA | Continuous | No significant difference |
| Wagner 2023 | Insulin resistance | Primary | Fasting glucose and insulin values | 15 months | NA | Continuous | No significant difference |
| Wagner 2023 | Hair cortisol | Secondary | ELISA kit | 12 months | NA | Continuous | Decreased in intervention group |
| Wagner 2023 | Hair cortisol | Secondary | ELISA kit | 15 months | NA | Continuous | Decreased in intervention group |
| Wagner 2023 | hsCRP | Secondary | Immunoturbidimetric assay | 12 months | NA | Continuous | No significant difference |
| Wagner 2023 | hsCRP | Secondary | immunoturbidimetric assay | 15 months | NA | Continuous | No significant difference |
| Wagner 2023 | Waist circumference | Secondary | Measured at the umbilicus with an inelastic tape | 12 months | NA | Continuous | No significant difference |
| Wagner 2023 | SBP | Secondary | Omron, Hoffman Estates | 12 months | NA | Continuous | No significant difference |
| Wagner 2023 | DBP | Secondary | Omron, Hoffman Estates | 12 months | NA | Continuous | No significant difference |
| Wagner 2023 | LDL | Secondary | Spectrophotometry assay | 12 months | NA | Continuous | No significant difference |
| Wagner 2023 | HDL | Secondary | Spectrophotometry assay | 12 months | NA | Continuous | No significant difference |
| Wagner 2023 | TC | Secondary | Spectrophotometry assay | 12 months | NA | Continuous | No significant difference |
| Wagner 2023 | TG | Secondary | Spectrophotometry assay | 12 months | NA | Continuous | No significant difference |
| Wagner 2023 | TG | Secondary | Spectrophotometry assay | 15 months | NA | Continuous | No significant difference |
| Isaacs 2007 | SBP | Primary | OMRON-HEM | 10 weeks | NR | Continuous | Significantly reduced in intervention group |
| Isaacs 2007 | Physical activity (Minutes of moderate and/or vigorous activity) | Primary | NR | 10 weeks | NR | Continuous | No significant difference |
| Isaacs 2007 | Physical activity (Total minutes of activity) | Primary | NR | 10 weeks | NR | Continuous | No significant difference |
| Isaacs 2007 | Physical activity (Total minutes of activity) | Primary | NR | 6 months | NR | Continuous | No significant difference |
| Isaacs 2007 | Physical activity (Energy expenditure) | Primary | NR | 10 weeks | NR | Continuous | No significant difference |
| Isaacs 2007 | Physical activity (Energy expenditure) | Primary | NR | 6 months | NR | Continuous | No significant difference |
| Isaacs 2007 | Percentage body fat | Secondary | Bioimpedance | 10 weeks | NR | Continuous | No significant difference |
| Isaacs 2007 | Percentage body fat | Secondary | Bioimpedance | 6 months | NR | Continuous | No significant difference |
| Isaacs 2007 | Cardiorespiratory fitness | Secondary | PEF | 6 months | NR | Continuous | The advice group had a slight but significant increase in FEV1/FVC ratio at 10 weeks and both the advice and leisure centre groups at 6 months, owing to a greater decline in FVC than in FEV1, but there was no significant change in PEF |
| Isaacs 2007 | Health service usage | Secondary | Visits to general physician | 6 months | NR | Binary | More GP visits in intervention group than advise group |
| Isaacs 2007 | Cost | Secondary | ICER | 6 months | NR | $ | The estimated incremental cost-effectiveness ratio (ICER) for the leisure centre group compared with the control group, at 6 months, was £312/0.016, equivalent to a cost of approximately £19,500 per unit increase in SF-36 score |
| Isaacs 2007 | Adverse events | Secondary | Visits for chest pain | 6 months | NR | Binary | No significant difference |
| Isaacs 2007 | Adverse events | Secondary | Visits for aches and pains | 6 months | NR | Binary | No significant difference |
| Isaacs 2007 | Adverse events | Secondary | Visits for sprains | 6 months | NR | Binary | No significant difference |
| Isaacs 2007 | Adverse events | Secondary | Visits for falls | 6 months | NR | Binary | No significant difference |
| Isaacs 2007 | Adverse events | Secondary | Visits for fractures | 6 months | NR | Binary | No significant difference |
| Heisler 2022 | ED visits | Primary | NA | 12 months | NR | Binary | CHW program on average had fewer ED visits than control patients |
| Heisler 2022 | Ambulatory care visits | Primary | NA | 12 months | NR | Binary | No significant differences in average ambulatory care visits |
| Heisler 2022 | Hospital admissions | Primary | NA | 12 months | NR | Binary | No significant differences in overall hospitalizations |
| Heisler 2022 | ED Cost | Secondary | NA | 12 months | NR | $ | Lower ED costs in intervention group |
| Heisler 2022 | Outpatient costs | Secondary | NA | 12 months | NR | $ | No significant difference |
| Heisler 2022 | Inpatient costs | Secondary | NA | 12 months | NR | $ | No significant difference |
| Heisler 2022 | Total costs | Secondary | NA | 12 months | NR | $ | No significant difference |
| Rovner 2020 | ED visits | Primary | NA | 12 months | NR | $ | No significant difference |
| Rovner 2020 | Hospitalizations | Primary | NA | 12 months | NR | Binary | There were no treatment group differences in rates of hospitalisation |
| Rovner 2020 | PIMs | Secondary | Medications with high risk/safety ratios | 12 months | NR | Binary | No significant difference |
| Rovner 2020 | HbA1c | Secondary | Blood | 12 months | NR | Binary | No significant difference |
| Rovner 2020 | Diabetes self-management | Secondary | The Diabetes Self-Care Inventory Revised | 12 months | NR | Continuous | Greater improvement in intervention group |
| Rovner 2020 | Diabetes self-efficacy | Secondary | The 8-item Self-Efficacy for Diabetes | 12 months | NR | Continuous | Greater improvement in intervention group |
| Rovner 2020 | Depressive symptoms | Secondary | The Patient Health Questionnaire- 9 | 12 months | NR | Continuous | No significant difference |
| Islam 2023 | Moderate PA (min) | Uncategorised | Weekly | 6 months | RA | Continuous | No difference in recommended weekly PA between the group |
| Islam 2023 | Vigorous PA | Uncategorised | Weekly | 6 months | RA | Continuous | No difference in recommended weekly PA between the group |
| Islam 2023 | BP control | Primary | <140/90 mm Hg | 6 months | RA | Binary | Achieved significantly in intervention group compared to control group |
| Islam 2023 | General health | Uncategorised | Patient-Reported Outcomes Measurement Information System | 6 months | RA | Continuous | No significant difference |
| Islam 2023 | Carry out usual activities | Uncategorised | Patient-Reported Outcomes Measurement Information System | 6 months | RA | Continuous | No significant difference |
| Islam 2023 | Weight reduction | Uncategorised | NA | 6 months | RA | Continuous | Greater reduction in intervention group than control |
| Lamb 2002 | Physical activities | Primary | Postal questionnaire (Total time – moderate intensity activity in minutes) | 6 months | Self rated | Continuous | No significant difference |
| Lamb 2002 | Physical activities | Primary | Postal questionnaire (Total time – moderate intensity activity in minutes) | 12 months | Self rated | Continuous | No significant difference |
| Lamb 2002 | BMI | Secondary | Kg/m^2^ | 6 months | RA | Continuous | No significant difference |
| Lamb 2002 | BMI | Secondary | Kg/m^2^ | 12 months | RA | Continuous | No significant difference |
| Lamb 2002 | Cholesterol | Secondary | Blood | 6 months | RA | Continuous | No significant difference |
| Lamb 2002 | Cholesterol | Secondary | Blood | 12 months | RA | Continuous | No significant difference |
| Lamb 2002 | SBP | Secondary | Digital monitor (Model UA-702, A&D Ltd, Tokyo, Japan) | 6 months | RA | Continuous | No significant difference |
| Lamb 2002 | DBP | Secondary | Digital monitor (Model UA-702, A&D Ltd, Tokyo, Japan) | 6 months | RA | Continuous | No significant difference |
| Lamb 2002 | Aerobic capacity | Secondary | NA | 6 months | RA | Continuous | No significant difference |
| Lamb 2002 | Aerobic capacity | Secondary | NA | 12 months | RA | Continuous | No significant difference |
| Kiely 2024 | Healthcare utilization | Secondary | GP visits | 1 month | Research team | Continuous | Control group had a significantly greater decrease in GP visits |
| Kiely 2024 | Healthcare utilization | Secondary | GP nurse visits | 1 month | Research team | Continuous | Control group had a significantly greater decrease in GP nurse visits |
| Kiely 2024 | Healthcare utilization | Secondary | Out of Hours | 1 month | Research team | Continuous | No significant difference |
| Kiely 2024 | Healthcare utilization | Secondary | Accident and Emergency | 1 month | Research team | Continuous | No significant difference |
| Kiely 2024 | Healthcare utilization | Secondary | Length of Hospital Stay | 1 month | Research team | Continuous | No significant difference |
| Kiely 2024 | Healthcare utilization | Secondary | Out patient department | 1 month | Research team | Continuous | No significant difference |
| Kiely 2024 | Health status | Secondary | EQ-VAS | 1 month | Research team | Continuous | No significant difference |
| Kiely 2024 | Well-being index | Secondary | ICECAP-A | 1 month | Research team | Continuous | No significant difference |
| Kiely 2024 | Patient activation measures | Secondary | PAM | 1 month | Research team | Continuous | No significant difference |
| Kiely 2024 | Multimorbidity Treatment burden | Secondary | NA | 1 month | Research team | Continuous | No significant difference |
| Kiely 2024 | Activity status | Secondary | Frenchay Activity Index | 1 month | Research team | Continuous | No significant difference |
| Kiely 2024 | Healthcare costs Euro € | Secondary | NA | 1 month | Research team | € | No significant difference |
| Kiely 2024 | Capability and wellbeing | Secondary | QALY (ICECAP-A) | 1 month | Research team | € | The intervention was associated with an increase in mean QALYs of 0.015 (-0.005, 0.035) [p = 0.852] per patient based on the ICECAP-A utility score |
| Kiely 2024 | Mean Total Cost | Secondary | NA | 1 month | Research team | € | €1,191.48 for the intervention group and €225.20 for the control group |

**Abbreviations**: NA: not applicable/not available; NR: not reported; BMI: body mass index; RA: research assistant; SBP: systolic blood pressure; DBP: diastolic blood pressure; BP: blood pressure; CHW: community healthcare worker; PA: physical activity; ED: emergency department; LDL: Low density lipoprotein; HDL: High density lipoprotein; TG: triglycerides; TC: total cholesterol; hsCRP: High-Sensitivity C-Reactive Protein ; HbA1c: glycated haemoglobin; PTSD: Post-Traumatic Stress Disorder; HRQOL: health related quality of life; RMSSD: Root Mean Square of Successive Differences; POLST: Physician Orders for Life Sustaining Treatment; GP: general physician; PEF: Peak expiratory flow; FEV: Forced Expiratory Volume; FCV: Forced Vital Capacity; ICER: incremental cost-effectiveness ratio; GOC: goal of care;

**Supplemental Table 16. GRADE assessment for the outcomes**

| **Certainty assessment** | | | | | | | **No. of patients** | | **Effect** | | **Certainty** |
| --- | --- | --- | --- | --- | --- | --- | --- | --- | --- | --- | --- |
| **No of studies** | **Study design** | **Risk of bias** | **Inconsistency** | **Indirectness** | **Imprecision** | **Other considerations^f^** | **Experimental** | **Control** | **Relative (95% CI)** | **Absolute (95% CI)** |  |
| **Systolic blood pressure** | | | | | | | | | | | |
| 11 | randomised trials | not serious | not serious | not serious | not serious | none | 1586 | 1231 | - | MD 2.69 lower (-5.36 to -0.02) | ⨁⨁⨁⨁ High |
| **Blood pressure control** | | | | | | | | | | | |
| 2 | randomised trials | serious^a^ | not serious | not serious | serious^b^ | none | 127/191 (66.5%) | 64/168 (38.1%) | OR 3.25 (2.11 to 5.03) | 286 more per 1000 (from 184 more to 375 more) | ⨁⨁◯◯  Low |
| **Body mass index** | | | | | | | | | | | |
| 14 | randomised trials | not serious | not serious | not serious | not serious | none | 1907 | 1551 | - | MD 0.19 lower (-0.53 to 0.15) | ⨁⨁⨁⨁ High |
| **Anxiety** | | | | | | | | | | | |
| 6 | randomised trials | not serious | not serious | not serious | serious^c^ | none | 929 | 937 | - | SMD 0.21 SD lower (-0.48 to 0.06) | ⨁⨁⨁◯  Moderate |
| **Quality of life** | | | | | | | | | | | |
| 10 | randomised trials | not serious | not serious | not serious | not serious | none | 1596 | 1538 | - | SMD 0.15 higher (0.01 to 0.29) | ⨁⨁⨁⨁ High |
| **Physical activity** | | | | | | | | | | | |
| 7 | randomised trials | not serious | not serious | not serious | not serious | none | 1848 | 1561 | - | SMD 0.16 higher (0.06 to 0.25) | ⨁⨁⨁⨁ High |
| **Depression** | | | | | | | | | | | |
| 7 | randomised trials | not serious | not serious | not serious | not serious | none | 567 | 520 | - | SMD 0.23 lower (-0.38 to 0.08) | ⨁⨁⨁⨁ High |
| **HbA1C** | | | | | | | | | | | |
| 5 | randomised trials | not serious | serious^d^ | not serious | not serious | none | 681 | 657 | - | MD 0.27 lower (-0.60 to 0.07) | ⨁⨁⨁◯  Moderate |
| High-density lipoprotein | | | | | | | | | | | |
| 2 | randomised trials | not serious | not serious | not serious | not serious | none | 584 | 325 | - | MD 0.49 lower (-1.45 to 0.48) | ⨁⨁⨁⨁ High |
| Low-density lipoprotein | | | | | | | | | | | |
| 3 | randomised trials | not serious | serious^e^ | not serious | not serious | none | 625 | 382 | - | MD 3.15 lower (-10.60 to 4.30) | ⨁⨁⨁◯  Moderate |

CI: confidence interval; MD: mean difference; OR: odds ratio; SMD: standardised mean difference

a. Data were originated from two studies, which were rated as some concerns and high-risk studies
b. Overall sample size was too small
c. The 95% confidence interval includes the possibility of an effect too small to be a meaningful effect
d. Serious inconsistency (substantial heterogeneity I^2^ = 97%), point estimates and confidence intervals vary considerably
e. Serious inconsistency (substantial heterogeneity I^2^ = 93%), point estimates and confidence intervals vary considerably
f. Other considerations include publication bias, large effect, plausible confounding and dose response gradient

**Supplemental Table 17. Narrative outcome assessment**

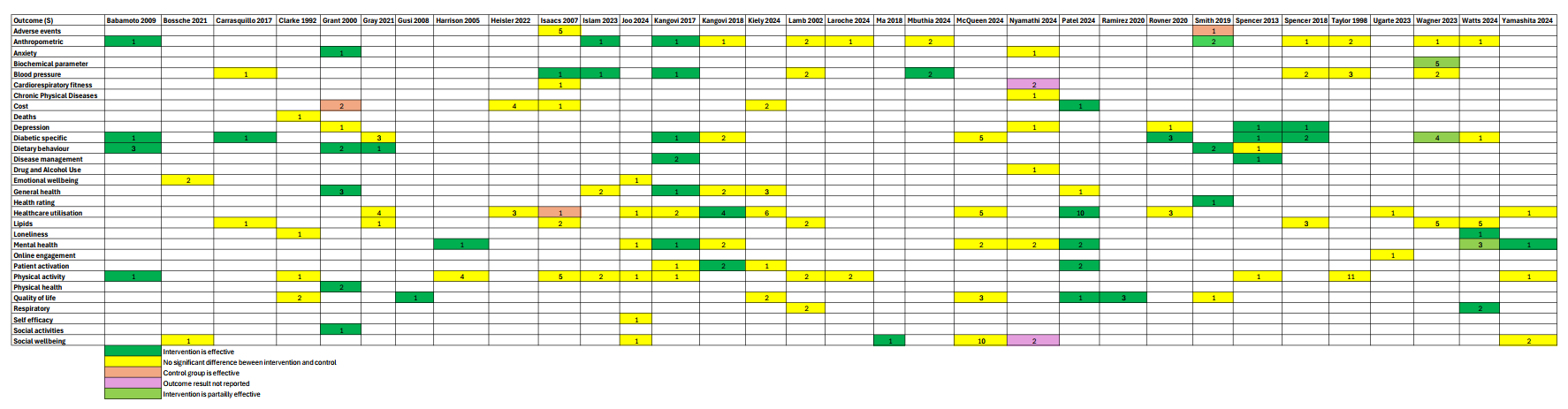
